# Temporal and Geographic Variability of International Urinary Phthalates in Humans: A Systematic Review and Meta-Analysis of Biomonitoring Data

**DOI:** 10.1101/2024.11.11.24317001

**Authors:** Jonathan M. Acevedo, Linda G. Kahn, Kristyn A. Pierce, Vittorio Albergamo, Anna Carrasco, Robbie S.J. Manuel, Marissa Singer Rosenberg, Leonardo Trasande

## Abstract

Phthalates are endocrine-disrupting chemicals (EDCs) that alter hormone functions throughout the lifespan. Growing awareness of the adverse health effects of phthalate exposure has led to regulating certain phthalates in the United States, Canada, and Europe. However, international comparisons of urinary phthalate metabolite concentrations as biomarkers of exposure are sparse, and few studies have controlled for cohort-specific variables like pregnancy.

We aimed to examine trends in urinary phthalate monoester metabolite concentrations in non-occupationally exposed populations globally, excluding locations where representative data are already available at the country level.

We systematically reviewed studies published between 2000 and 2023 that reported urinary phthalate monoester concentrations. We examined changes in metabolite concentrations across time, controlling for region, age, and pregnancy status, using mixed-effects meta-regression models with and without a quadratic term for time. We identified heterogeneity using Cochran’s Q-statistic and I^2^ index, adjusting for it with the trim-and-fill method.

The final analytic sample consisted of 216 studies. Significant differences in phthalate metabolite concentrations were observed across regions, age groups, and between pregnant and non-pregnant cohorts. Our meta-regression identified a significant non-linear trend with time for Mono-n-butyl phthalate and Mono-isononyl phthalate concentration internationally and in Eastern and Pacific Asia (EPA). We also observed significant non-linear associations between time and Mono(2-ethyl-5-hydroxyhexyl) phthalate, Mono(2-carboxymethylhexyl) phthalate, and Mono(3-carboxypropyl) phthalate concentration internationally and/or in EPA, along with Mono(2-ethylhexyl) phthalate, Mono-carboxy-isononyl phthalate, and Mono-ethyl phthalate. Additionally, Mono-ethyl phthalate concentration showed a significant negative linear association with time in Latin America and Africa. Heterogeneity was high, indicating potential bias in our results.

Our findings indicate the need for increased awareness of phthalate exposure. Further analysis of the attributable disease burden and cost at regional and international levels, especially in low- and middle-income countries, is essential to understanding these and other EDCs impact on population health and the economy.

**Highlights:**

- Some phthalate levels significantly differed by region, age, and pregnancy status.
- Many phthalates had non-linear associations internationally from 2000 to 2023.
- MnBP and MiNP levels increased, driven by Eastern and Pacific Asia results.
- Most phthalate metabolites’ levels declined overall and region-specific over time.
- There was insufficient data on phthalate metabolite levels for many regions.

## Introduction

Phthalates were introduced in the 1920s and used as a replacement for volatile and odorous camphor in the manufacturing of plastics. In the 1950s, the development of di(2-ethylhexyl) phthalate (DEHP) quickly led to its adoption in commercial use and the production of polyvinyl chloride (PVC) for construction (Lyche, 2011). From 1970 to 2006, phthalate production more than doubled, increasing from 1.8 to 4.3 million tons globally, and expanded into other markets such as food packaging, cosmetics, medical supplies, paint, and pesticides (Habert et al., 2009).

Phthalate exposure occurs through inhalation, skin contact, and oral consumption. Pharmacokinetic analysis has suggested that phthalates have a relatively short half-life of 4 to 24 hours, varying based on chemical type and body composition. Prolonged exposure to phthalates has been associated with obesity, Type II diabetes, impaired ovarian function, lower sperm count and motility, respiratory complications, neurodevelopment delays/impairment, cancer, and cardiovascular mortality (Trasande et al., 2022; Wang and Qian, 2021).

Despite the increasing recognition of the adverse health effects associated with phthalate exposure, commercial use has remained robust, especially by companies that produce or distribute plastics in low- and middle-income countries (LMICs) or regions lacking strict regulation. In China, plastic production and consumption by sector has grown throughout the years, with primary plastics production increasing from 13.8 million metric tons in 2000 to 144.4 million metric tons in 2022, a trend that is expected to continue in the coming years. With this exponential increase in plastic production and the public’s increased consumption of plastic-containing goods and plastic-contaminated food and drink, it is expected that China’s general population will experience relatively high levels of phthalate exposure(Luan et al., 2021). Like China, various countries in Latin America have experienced heightened exposure to phthalates through the increased consumption of ultra-processed food and beverages containing traces of phthalates(Matos et al., 2021). Previous meetings of the Conferences of the Parties to the Basel and the Stockholm Conventions have also expressed concerns over the impact of plastic waste, exposure, and consumption, especially in Africa. Between 2009 and 2015, per capita plastic consumption has increased in African nations between 21 and 66 percent. The consistent production and use of phthalates involuntarily increases the general population’s risk of exposure and associated health risks (Babayemi et al., 2019).

In response to the ever-growing scientific evidence of the harmful impact of phthalates on human health, several high-income countries have adopted policies aiming to reduce or regulate the general population’s exposure. The United States (US) has banned any children’s toys or childcare articles, such as baby bottles and pacifiers, that contain more than 0.1% (1000 ppm) of di-(2-ethylhexyl) phthalate (DEHP), benzyl butyl phthalate (BBP), diisononyl phthalate (DiNP), diisobutyl phthalate (DiBP), di-n-pentyl phthalate (DnPP), di-n-hexyl phthalate (DnHP), and dicyclohexyl phthalate (DCHP) (U.S. Environmental Protection Agency, 2017; United States Consumer Product Safety Commission, 2019). The European Union has implemented the same regulation as the US and required authorization to use specific phthalates in any medical devices or packaging, food contact material, or any mixture containing greater than 0.1% phthalate by weight. Furthermore, Europe has implemented bans on cosmetics, textiles, clothing, and footwear containing specific phthalates and phthalate mixtures with a concentration greater than 1.0 mg/g (European Environment Agency, 2022; Francaise, 2023). These regulatory actions and increased awareness of the hazardous effects of phthalates have contributed to a decline in exposure throughout the years in both regions (European Environment Agency, 2022; U.S. Environmental Protection Agency, 2017).

While national biomonitoring surveys and observational research measuring phthalate exposure and trends are occurring in the US, Europe, and Canada, such information is not readily available in other regions. Of the few studies that have analyzed trends in other regions, the majority focused on high-income countries rather than LMICs. However, one study that compared temporal changes in phthalate concentrations in Eastern and Pacific Asia (EPA) with the US and Canada documented increases in several phthalate monoester metabolite concentrations in the EPA region between 2009 and 2019 (Domínguez-Romero et al., 2023). An analysis conducted within the Programming Research in Obesity, Growth, Environment, and Social Stressors (PROGRESS) Study in Mexico reported significantly different phthalate monoester metabolite concentrations compared with contemporary US and Sweden cohorts (Shin et al., 2020). Building upon these studies, we performed a systematic review of the literature to extract measures of phthalate monoester metabolite concentrations in non-occupationally exposed populations globally, excluding locations where representative data are already available at the country level, and analyzed these data for trends over time.

## Methods

### 2.1 General Description

This systematic review and meta-analysis follows the Preferred Reporting Items for Systematic Reviews and Meta-Analysis (PRISMA) guidelines. Our protocol was registered in the International Prospective Register of Systematic Reviews (PROSPERO) on February 27^th^, 2024 (registration code: CRD42023481172).

### 2.2. Search strategy

We searched PubMed, Ovid, and Web of Science for Boolean combinations of the following MeSH terms: “Phthalates”, “Mono-2-Ethylhexyl Phthalate”, “MEHP”, “Mono-2-ethyl-5-hydroxyhexy Phthalate”, “MEHHP”, “Mono-2-ethyl-5-oxohexyl Phthalate”, “MEOHP”, “Mono-2-ethyl-5-oxohexyl Phthalate”, “MEOHP”, “Mono-n-butyl Phthalate”, “MnBP”, “Mono(2-ethyl-5-carboxypentyl) Phthalate”, “MECPP”, “Mono(2-carboxymethylhexyl) Phthalate”, “MCMHP”, “Mono-carboxy-isooctyl Phthalate”, “MCOP”, “Mono-isononyl Phthalate”, “MiNP”, “Mono-oxo-isononyl Phthalate”, “MOiNP”, “Mono-hydroxy-isononyl Phthalate”, “OH-MiNP”, “Mono-carboxy-isononyl Phthalate”, “MCiNP”, “Mono(3-carboxypropyl) Phthalate”, “MCPP”, “Mono-octyl Phthalate”, “MOP”, “Monobenzyl Phthalate”, “MBzP”, “Mono-isodecyl Phthalate”, “MiDP”, “Mono-ethyl Phthalate”, “MEP”, “Mono-isobutyl Phthalate”, “MiBP”, “Di(2-ethylhexyl) Phthalate”, “DEHP”, “Diethyl Phthalate”, “DEP”, “Benzyl butyl phthalate”, “BBP”, “Diisononyl Phthalate”, “DiNP”, “Diisodecyl Phthalate”, “DiDP”, “Di-n-octyl phthalate”, “DnOP”, “Dibutyl Phthalate”, “DBP”, “Endocrine Disruptors”, “Endocrine Disruption”, “Endocrine Disrupting Chemical”, “EDs”, and “Xenoestrogens”. These terms were combined with relevant biological sampling vocabulary such as “Urine,” “Plasma,” “Serum,” “Human Tissue,” “Body Fluids,” “Saliva,” “Sweat,” and “Blood.” Additional combinations included “Concentrations,” “Levels,” “Biomonitoring,” “Exposure,” and “Human Exposures.“

### 2.3. Eligibility criteria

We selected studies that reported urinary monoester phthalate metabolite concentrations within their population (arithmetic mean [AM] and standard deviation [SD], geometric mean [GM] and 95% confidence interval [CI], and median and Interquartile Range [IQR]), written in English, and published between January 1, 2000, and December 1, 2023. We also collected secondary citations in articles identified for full-text screenings. We excluded articles with non-human species and populations from locations where representative data are already available at the country level (i.e., the US, Canada, and Europe). Additionally, we excluded articles and data from cohorts that aimed to control or alter the participants’ phthalate exposure and cohorts that focused on participants with chronic or severe health conditions such as diabetes, infertility, asthma, endometriosis, and cancer (thyroid, breast, and prostate). We also excluded cohorts highly exposed based on residency or occupation, as we were interested in tracking routine exposure among the general population.

Furthermore, if two or more analyses were based on the same cohort, the one with the highest number of participants was used. If the analyses had the same population size, we selected the article that measured the greatest number of phthalates. In addition, we excluded articles that provided concentrations of parent diester phthalates rather than the monoester metabolites. Lastly, we excluded papers that lacked sufficient information to perform a regression analysis.

### 2.4. Quality Control and Data Extraction

After removing duplicates, two reviewers (J.M.A. and M.S.R.) used Covidence (Melbourne, Australia) to manage and review each article. They assessed each article’s title and abstract for relevancy and selected articles for a full-text screening. If an article’s eligibility was in question, a third reviewer (A.C.) was consulted to make the final decision.

Upon completing the full-text screening, the reviewers extracted publication details such as author(s), publication year, sample size, and cohort characteristics (e.g., age range, geographical region, pregnant verse non-pregnant populations) from the eligible articles. The geographical areas were determined based on where the study’s sampling occurred (Latin America, Africa, Asia, or Australia; Table 1) (The World Bank, 2024). Due to Asia’s high quantity of articles, we used the World Bank definitions to divide the region into sub-regions: Eastern and Pacific Asia (EPA) and Middle East and South Asia (MESA; Table 1) (World Bank Group, 2024a; World Bank Group, 2024b; World Bank Group, 2024c). We categorized study populations into the following age groups: “youth” (age <18 years), “adults” (18 year and older), and “Mixed” (age range includes both youth and adults). Additionally, we documented whether pregnant individuals were included in the sample. Articles that provided phthalate measures for more than one subset of interest were allowed to contribute multiple data points to this analysis. For instance, articles providing measures for pregnant women and their children were recorded as two distinct observations (“youth” and “adults”).

**Table 1:** Countries and Regions of Interest.

| International Region | Countries |
| --- | --- |
| Latin America | Antigua and Barbuda, Argentina, Aruba*, The Bahamas, Barbados, Belize, Bolivia, Brazil, British Virgin Islands*, Cayman Islands*, Chile, Colombia, Costa Rica, Cuba, Curacao*, Dominica, Dominican Republic, El Ecuador, El Salvador, Grenada, Guatemala, Guyana, Haiti, Honduras, Jamaica, Mexico, Nicaragua, Panama, Paraguay, Peru, Puerto Rico*, Sint Maarten (Dutch part)*, St. Kitts and Nevis, St. Lucia, St. Martin (French part)*, St. Vincent and the Grenadines, Suriname, Trinidad and Tobago, Turks and Caicos Islands*, Uruguay, Venezuela, RB Virgin Islands (U.S.) |
| Africa | Algeria, Angola, Benin, Botswana, Burkina Faso, Burundi, Cabo Verde, Cameroon, Central African Republic, Chad, Comoros, Congo Dem. Rep., Congo Rep., Cote d'Ivoire, Djibouti, Egypt, Equatorial Guinea, Eritrea, Eswatini, Ethiopia, Gabon, The Gambia, Ghana, Guinea, Guinea-Bissau, Kenya, Lesotho, Liberia, Libya, Madagascar, Malawi, Mali, Mauritania, Mauritius, Mozambique, Namibia, Niger, Nigeria, Rwanda, Sao Tome and Principe, Senegal, Seychelles, Sierra Leone, Somalia, South Africa, South Sudan, Sudan, Tanzania, Togo, Tunisia, Uganda, Zambia, Zimbabwe |
| Asia** | **Eastern and Pacific Asia (EPA):** American Samoa*, Brunei Darussalam, Cambodia, China, Fiji, French Polynesia*, Guam*, Hong Kong SAR, China, Indonesia, Japan, Kiribati, Korea Dem. People's Rep., Korea Rep., Lao PDR, China Macao SAR, Malaysia, Marshall Islands, Fed. Sts. Micronesia, Mongolia, Myanmar, Nauru, New Caledonia*, New Zealand, Northern Mariana Islands*, Palau, Papua New Guinea, Philippines, Samoa, Singapore, Solomon Islands, Thailand, Timor-Leste, Tonga, Tuvalu, Vanuatu, Viet Nam<br>**Middle East and South Asia (MESA):** Afghanistan, Bahrain, Bangladesh, Bhutan, India, Iran, Islamic Rep., Iraq, Israel, Jordan, Kuwait, Lebanon, Maldives, Malta*, Morocco, Nepal, Oman, Pakistan, Qatar, Saudi Arabia, Sri Lanka, Syrian Arab Republic, United Arab Emirates, West Bank and Gaza, Yemen, Rep. |
| Australia | Australia*** |

\* These countries were a part of the region but were not analyzed since they are part of the US and European via territories.
\*\* Asia was subdivided into geographical subsets based on World Bank categories due to the abundance of literature within the region.
\*\*\* Australia is categorized as part of Eastern and Pacific Asia but for consistency identified it as its own region.

Along with publication and population details, outcome information, including the phthalate monoester type, chemical concentration, analytical method’s limit of detection (LOD)/quantification (LOQ), percentiles, and sampling year, was recorded. If a phthalate metabolite concentration was below the LOD, we imputed the value as LOD/√2. Based on our search, we selected the following phthalates monoesters based on the how often they were within the literature and their use in industry practices: mono(2-carboxymethylhexyl) phthalate (MnBP); mono(2-ethyl-5-carboxypentyl) phthalate (MEHP); mono(2-ethyl-5-hydroxyhexyl) phthalate (MEHHP); mono(2-ethyl-5-oxohexyl) phthalate (MEOHP); mono(2-ethyl-5-carboxypentyl) phthalate (MECPP); mono(2-carboxymethylhexyl) phthalate (MCMHP); mono-carboxy-isooctyl phthalate (MCOP); mono-isononyl phthalate (MiNP); mono-oxo-isononyl phthalate (MOiNP); Mono-hydroxy-isononyl phthalate (OH-MiNP); mono-carboxy-isononyl phthalate (MCiNP); mono(3-carboxypropyl) phthalate (MCPP); mono-octyl phthalate (MOP); monobenzyl phthalate (MBzP); mono-isodecyl phthalate (MiDP); mono-ethyl phthalate (MEP); and mono-isobutyl phthalate (MiBP). These monoesters are metabolites broken down from their diester parent compounds, as shown in Supplement Table 1. Phthalate metabolite concentrations were documented as AM (± SD), geometric mean (95% CI), and/or median (IQR or minimum/ maximum). The sampling year was defined as the year urine samples were collected or the midpoint if the study was conducted over more than one year.

While the majority of the studies reported phthalate monoester concentrations in their unadjusted state (ng/mL), some articles provided phthalate concentrations adjusted for creatinine (ng/mg). To harmonize these creatinine-adjusted concentrations with the creatinine-unadjusted concentrations, we used the following equation based on Park’s study (Park et al., 2016):

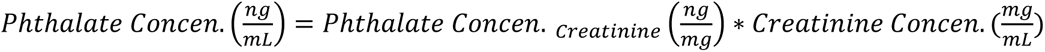

*Equation 1: Adjustment for creatinine levels within urinary biomonitoring samples*.

We used the age-specific creatinine concentration previously quantified by a prior study to adjust the phthalate monoester concentration (Park et al., 2016). If a study provided a monoester concentration for a population that included two or more predefined age groups given by Park et al. (2016), the average creatinine level for the general population, 0.91 mg/mL, was used to adjusted the concentration it approximate unadjusted value (Park et al., 2016). Further details regarding the creatinine concentration within each age group defined by Park et al. (2016) can be found in Supplement Table 2. Below is a worked example of converting creatinine-adjusted phthalates concentration into its unadjusted form based on the average creatinine concentration:

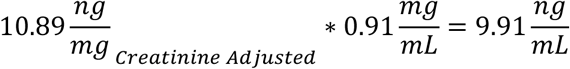

*Example 1: Converting phthalate concentration adjusted for creatinine to phthalate concentration unadjusted for creatinine*.

This study utilized phthalate AM concentrations in our study since we want to acknowledge the variability across different populations, regions, and age groups within the general population’s exposure to phthalates. When the phthalate concentration’s AM was absent, we used either the study’s GM or median value as a substitute. We also estimated missing SD values for several studies by dividing the IQR by the empirical constant of 1.35 (SD = IQR / 1.35) (Higgins, 2008) or by using the median, maximum, and minimum values, following methods published previously (Wan et al., 2014).

Study-specific standard errors (SE) were required for our weighted regression models. If missing, we used the formula SE = SD/√n, with n being the total study population. When the median was substituted for the AM, we applied the formula SE = (1.253*SD)/√n to adjust for variability caused by the distribution of the sample median (Hojo and Pearson, 1931).

### 2.5. Statistical Analysis

Mean phthalate monoester concentrations were compared across international regions, age groups, and pregnant vs. non-pregnant cohorts using either t-tests or analysis of variance (ANOVA) to identify significant differences in exposure levels.

We used mixed-effects regression models to examine the change in urinary monoester phthalate concentrations across the years in which the studies sampled their populations, adjusted for age group, pregnancy status, and region. We also performed a covariate-adjusted meta-regression model weighted for study-specific SE to enhance the precision of the estimated associations between time and each mean phthalate concentrations (DerSimonian and Laird, 1986). In addition, we stratified our meta-regression models by region to examine region-specific trends in phthalate exposure over time.

We then repeated both unstratified and stratified covariate-adjusted meta-regression analyses including a quadratic term for time (year^2^). By adding the quadratic term, we were able to evaluate whether there was a non-linear relationship between time and mean phthalate concentration and, if so, the shape of the exponential curve. To avoid multicollinearity when adding the quadratic term to the meta-regressions, we centered the sampling year variable by subtracting the study’s sampling year from the calculated average sampling year.

We calculated the pooled mean concentration and 95% CI for each phthalate globally and by region. In addition to calculating the pooled means, we collected Cochran’s Q and I^2^ to determine the presence of heterogeneity within our dataset. If Cochran’s Q and/or an I^2^ value were high, we interpreted the data as having significant levels of heterogeneity. We implemented the trim-and-fill method to counteract the effects associated with high heterogeneity (Wang et al., 2012).

Additionally, we calculated the combined and region-specific predicted mean and SD exposure trend for each monoester phthalate across five periods (2003, 2008, 2013, 2018, and 2023) using a meta-regression model adjusted for all covariates except region. When we incorporated the geographical region into our adjustment, we used each region’s respective beta coefficient value to estimate the predicted statistics at each time point. With the predicted mean and SD value, we obtained the estimated concentration at the 10^th^, 25^th^, 50^th^, 75^th^, and 95^th^ percentiles using the inverse of the normal cumulative distribution at each time point.

Lastly, we performed two sensitivity meta-analysis, one excluding studies with GM and the other excluding AM values. We applied a predetermined percentage of 15% to assess whether the use of GM or AM meaningfully affected the results of our analysis.

All hypothesis tests were two-sided, and we used a Type 1 error rate of 0.05 to determine statistical significance. Analyses were conducted using Stata statistical software version 16.0 and R version 4.3.1.

## Results

### 3.1 Characteristics of the included studies

Our search yielded 27,759 articles (Figure 1). After removing duplicates and conducting the initial title and abstract screening, we selected 1,930 articles for a full-text screening. We excluded 1,714 articles during the process due to the articles failing to meet the eligibility criteria previously stated in section 2.3 and seen in Figure 1. Additionally, we identified two secondary citations obtained from the literature that met the criteria for the study. In total, we selected 216 articles to include in our final analysis (Abdo et al., 2023; Aimuzi et al., 2022; Ait Bamai et al., 2015; Al-Saleh et al., 2021; Amin et al., 2018; Asimakopoulos et al., 2016; Bai et al., 2022; Bao et al., 2022; Bao et al., 2015; Barwon Infant Study Investigator et al., 2020; Berman et al., 2009; Binder et al., 2018; Bustamante-Montes et al., 2021; Cao et al., 2020; Chang et al., 2022; Chang et al., 2017; Chang et al., 2021; Chen et al., 2017a; Chen et al., 2015; Chen et al., 2021a; Chen et al., 2023a; Chen et al., 2019a; Chen et al., 2021b; Chen et al., 2017b; Chen et al., 2017c; Chen et al., 2023b; Chen et al., 2019b; Chen et al., 2022; Cheng et al., 2021; Cho et al., 2010; Choi et al., 2014; Choi et al., 2019; Chu et al., 2021; Cohen-Eliraz et al., 2023; Colacino et al., 2011; Cui et al., 2022; Darvishmotevalli et al., 2019; Deng et al., 2022; Ding et al., 2019a; Ding et al., 2019b; Dong et al., 2020; Dong et al., 2018; Dong et al., 2019; Dong et al., 2017; Dong et al., 2022; Duan et al., 2021; Duh et al., 2023; Fu et al., 2023; Gao et al., 2016; Gao et al., 2022; Gao et al., 2017; Guo et al., 2011; Han et al., 2019; He et al., 2021; He et al., 2019; Hong et al., 2017; Hong et al., 2009; Hou et al., 2015; Hsia et al., 2022; Hsu et al., 2012; Hu et al., 2017; Hu et al., 2023; Huang et al., 2017; Huang et al., 2018a; Huang et al., 2018b; Huang et al., 2014; Huang et al., 2015; Huang et al., 2022a; Huang et al., 2022b; Hwang et al., 2022; Hyun Kim et al., 2018; Irvin et al., 2010; Itoh et al., 2007; Jia et al., 2022; Jiang et al., 2018; Jin et al., 2023; Jo et al., 2016; Jung et al., 2019; Jung et al., 2022; Kang et al., 2019; Kim et al., 2020; Kim et al., 2017; Kim et al., 2018a; Kim et al., 2016a; Kim et al., 2013; Kim et al., 2021; Kim et al., 2018b; Kim et al., 2016b; Kim et al., 2014a; Kim et al., 2014b; Kim et al., 2018c; Kuo et al., 2015; Lee et al., 2020a; Lee et al., 2020b; Lee et al., 2020c; Lee et al., 2019a; Lee et al., 2019b; Lee et al., 2021; Lee et al., 2023; Lee et al., 2017; Lee et al., 2018; Lee et al., 2019c; Lee et al., 2020d; Li et al., 2019a; Li et al., 2023a; Li et al., 2024; Li et al., 2019b; Li et al., 2021a; Li et al., 2023b; Li et al., 2023c; Li et al., 2019c; Li et al., 2018; Li et al., 2021b; Li et al., 2020; Liao et al., 2018a; Liao et al., 2022a; Liao et al., 2021; Liao et al., 2018b; Liao et al., 2022b; Lien et al., 2018; Lim et al., 2020; Lin et al., 2020; Lin et al., 2011a; Lin et al., 2011b; Liu et al., 2020a; Liu et al., 2023; Liu et al., 2020b; Liu et al., 2022; López-Carrillo et al., 2010; Lu et al., 2020; Luo et al., 2022; Lyu et al., 2022a; Ma et al., 2022; Medellin-Garibay et al., 2023; Meeker et al., 2009; Menanzambi et al., 2021; Merida-Ortega et al., 2016; Miao et al., 2020; Mohanto et al., 2023; Mok et al., 2021; Mok et al., 2022; Nguyen et al., 2022; On et al., 2021; Park et al., 2019; Park et al., 2015; Park et al., 2021; Rocha et al., 2017; Rodriguez-Baez et al., 2022; Rodriguez Arreola and Peregrina-Lucano, 2021; Shen et al., 2015; Shi et al., 2015; Shih et al., 2022; Sobolewski et al., 2017; Song et al., 2019; Su et al., 2019; Sun et al., 2016; Suzuki et al., 2010; Suzuki et al., 2009; Suzuki et al., 2012; t Mannetje et al., 2021; Tang et al., 2020a; Tian et al., 2023; Toshima et al., 2012; Tsai et al., 2021; Tsai et al., 2018; Velarde et al., 2022; Wang et al., 2015; Wang et al., 2013; Wang and Karmaus, 2015; Wang et al., 2014; Wang et al., 2023; Wang et al., 2021a; Wang et al., 2020; Wang et al., 2021b; Wang et al., 2018a; Wang et al., 2018b; Watkins et al., 2017; Wen et al., 2020; Wu et al., 2020a; Wu et al., 2018; Wu et al., 2020b; Wu et al., 2022; Wu et al., 2017a; Wu et al., 2017b; Xia et al., 2018; Xie et al., 2015; Xu et al., 2020; Yang et al., 2021; Yang et al., 2023a; Yang et al., 2023b; Yang et al., 2017; Yang et al., 2018; Yang et al., 2022; Yao et al., 2019; Yoon et al., 2023; Yoshida et al., 2020; Yu et al., 2022; Yu et al., 2021; Yue et al., 2023; Zamora et al., 2023; Zang et al., 2023; Zhang et al., 2019; Zhang et al., 2022; Zhang et al., 2018a; Zhang et al., 2020; Zhang et al., 2024; Zhang et al., 2018b; Zhang et al., 2021; Zhao et al., 2022a; Zhao et al., 2016; Zhao et al., 2015; Zhao et al., 2022b; Zhong et al., 2022; Zhu et al., 2020; Zhu et al., 2016; Zhu et al., 2018).

**Figure 1:**
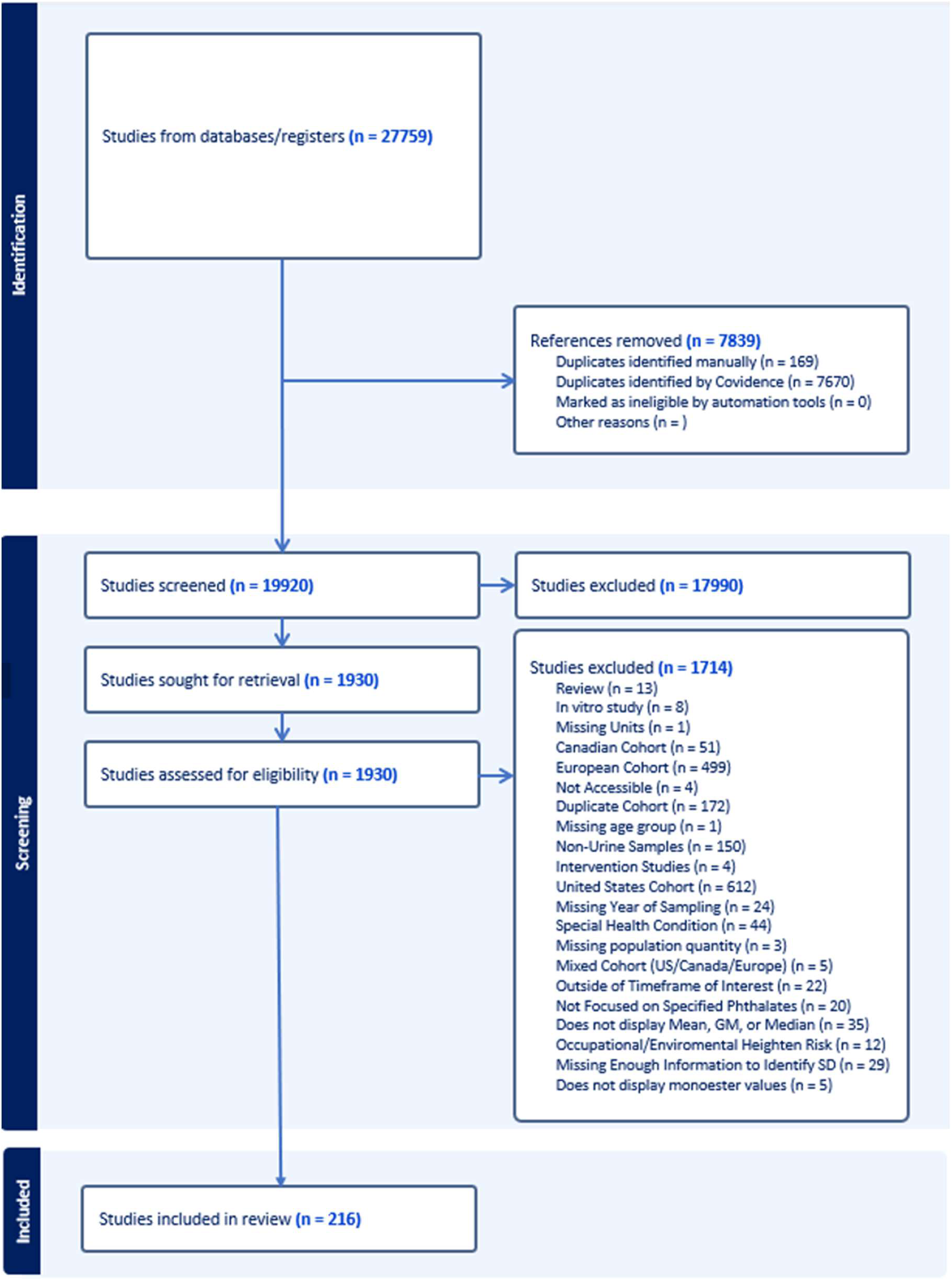
PRISMA flow chart of the study selection process.

Of the 216 articles in our final analysis, over 60% reported urinary concentrations (detectable or below the LOD) for at least one of the phthalate metabolites of di(2-ethylhexyl) phthalate (DEHP) and Dibutyl phthalate (DBP), while fewer than 5% reported a Diisodecyl Phthalate (DiDP) metabolite concentration. MEHHP was the most frequently reported monoester phthalate, with detectable urinary concentrations reported in 190 articles, while only five articles reported detectable urinary concentrations for MOiNP. Data from the EPA region comprised 60% or more of each phthalate monoester dataset. Additional information on the studies included is available in Supplement Table 3.

### 3.2 ANOVA, T-Tests, and Regressions

Table 2 displays statistical summaries and groupwise comparisons of phthalate monoester concentrations by region, age group, and pregnancy status. Several monoesters had statistically significant different mean concentrations across international regions. The MESA region had the highest concentrations of several phthalate monoesters (MnBP [160.48 ng/ml ± 120.93], MEHP [57.95 ng/ml ± 64.02], MEHHP [134.46 ng/mL ± 252.49], MCiNP [13.04 ng/ml ± 11.25], and MBzP [81.06 ng/ml ± 132.12]). Latin America and Australia had the lowest mean concentrations of these same phthalates (Latin America: MCiNP [0.82 ng/ml ± 0.34] and MBzP [3.11 ng/ml ± 1.68]; Australia: MnBP [28.35 ng/ml ± 9.40], MEHP [4.00 ng/ml ± 0.14], and MEHHP [13.85 ng/mL ± 5.87]).

**Table 2:**
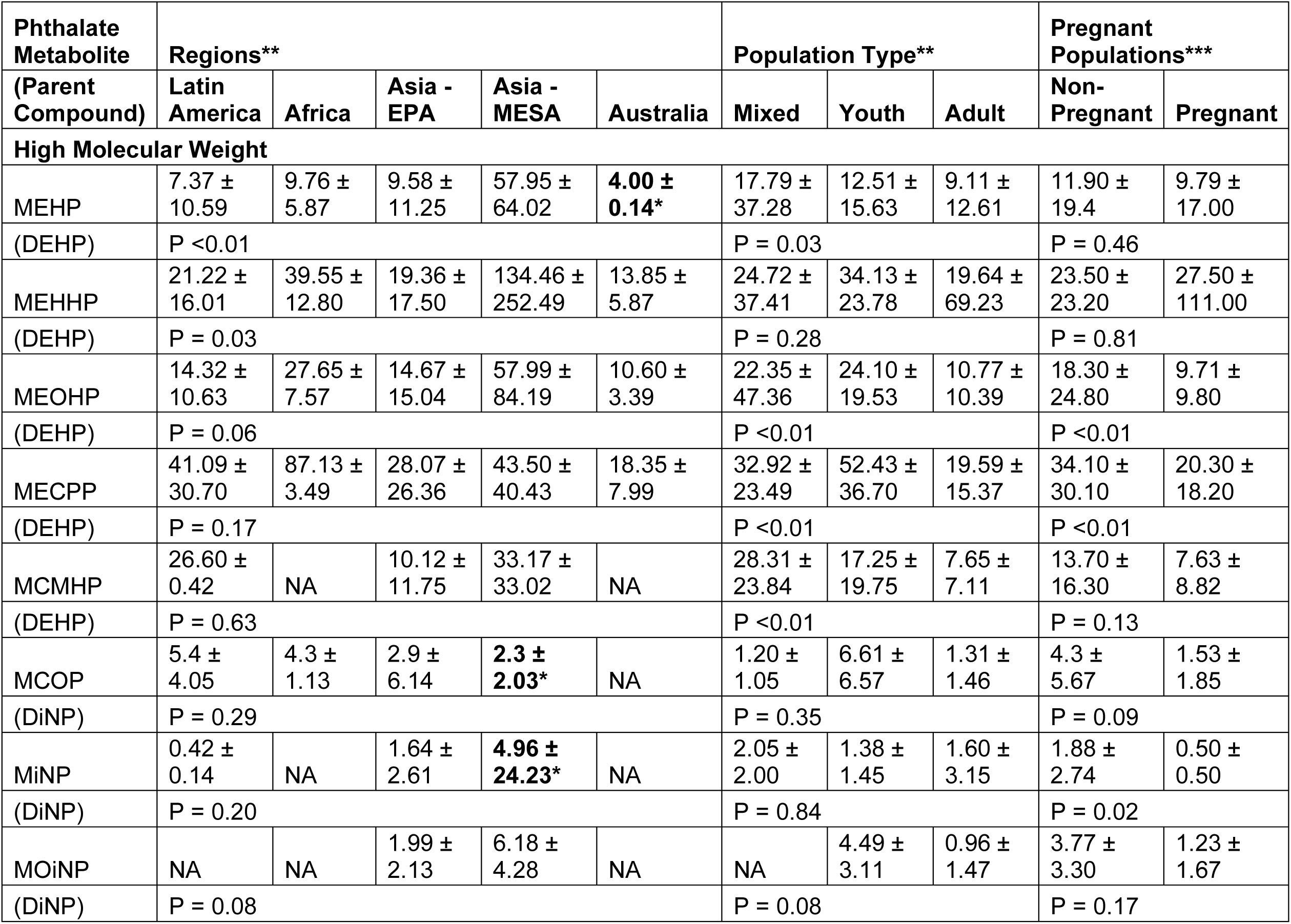
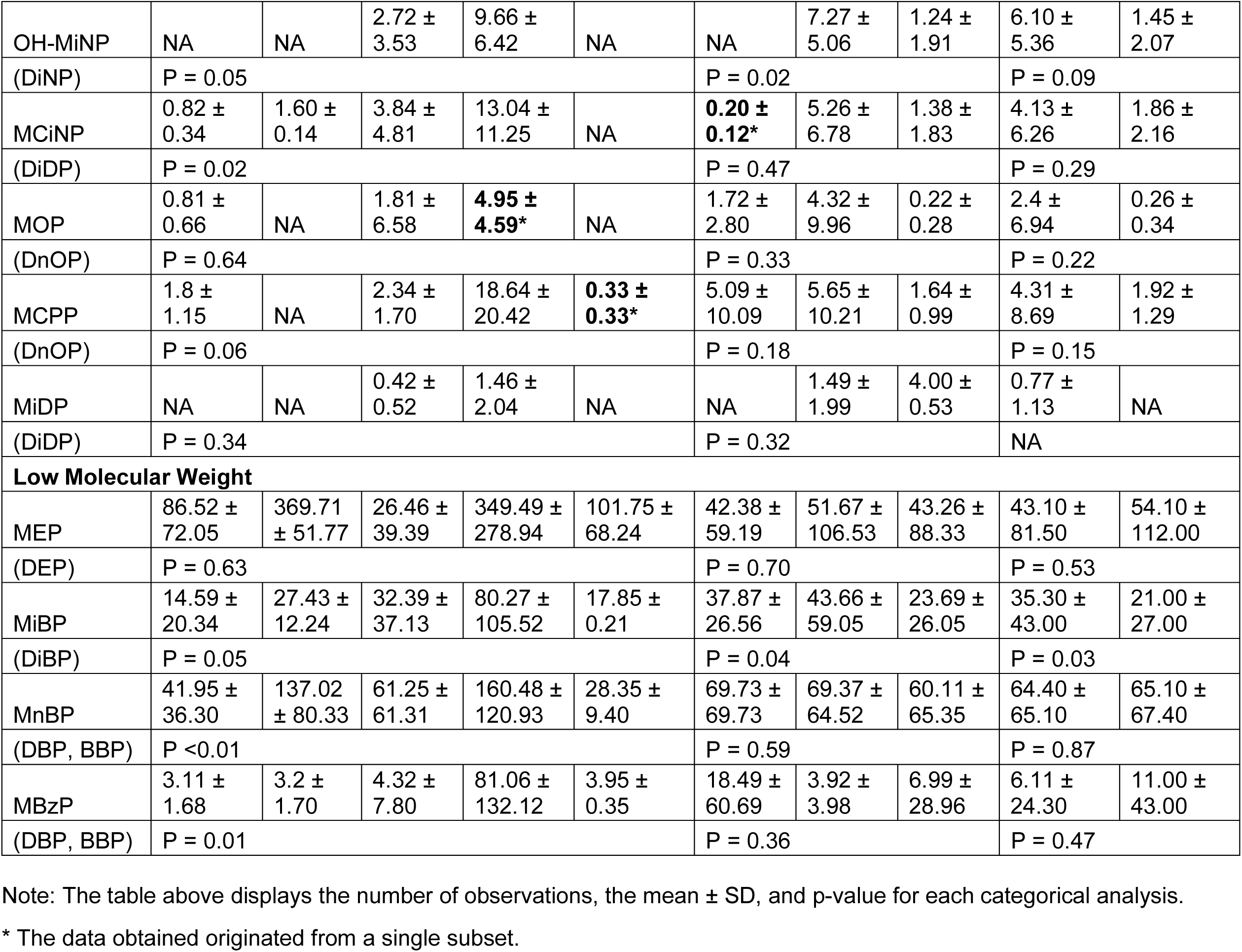
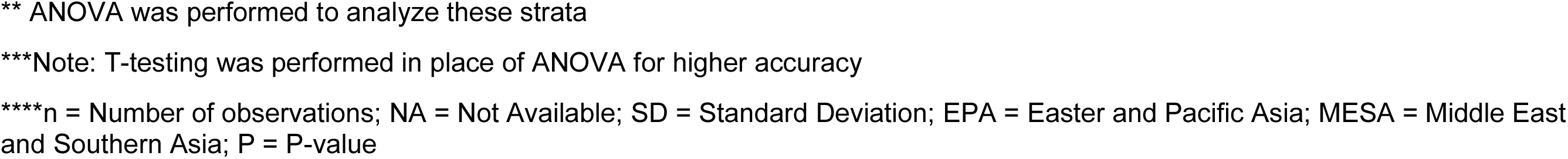
Mean urinary phthalate monoester concentrations (ng/mL) by region, age, and pregnancy status.

| Phthalate Metabolite | Regions** |  |  |  |  | Population Type** |  |  | Pregnant Populations*** |  |
| --- | --- | --- | --- | --- | --- | --- | --- | --- | --- | --- |
| (Parent Compound) | Latin America | Africa | Asia - EPA | Asia - MESA | Australia | Mixed | Youth | Adult | Non-Pregnant | Pregnant |
| **High Molecular Weight** |  |  |  |  |  |  |  |  |  |  |
| MEHP | 7.37 ± 10.59 | 9.76 ± 5.87 | 9.58 ± 11.25 | 57.95 ± 64.02 | **4.00 ± 0.14\*** | 17.79 ± 37.28 | 12.51 ± 15.63 | 9.11 ± 12.61 | 11.90 ± 19.4 | 9.79 ± 17.00 |
| (DEHP) | P < 0.01 |  |  |  |  | P = 0.03 |  |  | P = 0.46 |  |
| MEHHP | 21.22 ± 16.01 | 39.55 ± 12.80 | 19.36 ± 17.50 | 134.46 ± 252.49 | 13.85 ± 5.87 | 24.72 ± 37.41 | 34.13 ± 23.78 | 19.64 ± 69.23 | 23.50 ± 23.20 | 27.50 ± 111.00 |
| (DEHP) | P = 0.03 |  |  |  |  | P = 0.28 |  |  | P = 0.81 |  |
| MEOHP | 14.32 ± 10.63 | 27.65 ± 7.57 | 14.67 ± 15.04 | 57.99 ± 84.19 | 10.60 ± 3.39 | 22.35 ± 47.36 | 24.10 ± 19.53 | 10.77 ± 10.39 | 18.30 ± 24.80 | 9.71 ± 9.80 |
| (DEHP) | P = 0.06 |  |  |  |  | P < 0.01 |  |  | P < 0.01 |  |
| MECPP | 41.09 ± 30.70 | 87.13 ± 3.49 | 28.07 ± 26.36 | 43.50 ± 40.43 | 18.35 ± 7.99 | 32.92 ± 23.49 | 52.43 ± 36.70 | 19.59 ± 15.37 | 34.10 ± 30.10 | 20.30 ± 18.20 |
| (DEHP) | P = 0.17 |  |  |  |  | P < 0.01 |  |  | P < 0.01 |  |
| MCMHP | 26.60 ± 0.42 | NA | 10.12 ± 11.75 | 33.17 ± 33.02 | NA | 28.31 ± 23.84 | 17.25 ± 19.75 | 7.65 ± 7.11 | 13.70 ± 16.30 | 7.63 ± 8.82 |
| (DEHP) | P = 0.63 |  |  |  |  | P < 0.01 |  |  | P = 0.13 |  |
| MCOP | 5.4 ± 4.05 | 4.3 ± 1.13 | 2.9 ± 6.14 | **2.3 ± 2.03\*** | NA | 1.20 ± 1.05 | 6.61 ± 6.57 | 1.31 ± 1.46 | 4.3 ± 5.67 | 1.53 ± 1.85 |
| (DiNP) | P = 0.29 |  |  |  |  | P = 0.35 |  |  | P = 0.09 |  |
| MiNP | 0.42 ± 0.14 | NA | 1.64 ± 2.61 | **4.96 ± 24.23\*** | NA | 2.05 ± 2.00 | 1.38 ± 1.45 | 1.60 ± 3.15 | 1.88 ± 2.74 | 0.50 ± 0.50 |
| (DiNP) | P = 0.20 |  |  |  |  | P = 0.84 |  |  | P = 0.02 |  |
| MOiNP | NA | NA | 1.99 ± 2.13 | 6.18 ± 4.28 | NA | NA | 4.49 ± 3.11 | 0.96 ± 1.47 | 3.77 ± 3.30 | 1.23 ± 1.67 |
| (DiNP) | P = 0.08 |  |  |  |  | P = 0.08 |  |  | P = 0.17 |  |
| OH-MiNP | NA | NA | 2.72 ±<br>3.53 | 9.66 ±<br>6.42 | NA | NA | 7.27 ±<br>5.06 | 1.24 ±<br>1.91 | 6.10 ±<br>5.36 | 1.45 ±<br>2.07 |
| (DiNP) | P = 0.05 |  |  |  |  | P = 0.02 |  |  | P = 0.09 |  |
| MCiNP | 0.82 ±<br>0.34 | 1.60 ±<br>0.14 | 3.84 ±<br>4.81 | 13.04 ±<br>11.25 | NA | **0.20 ±**<br>**0.12\*** | 5.26 ±<br>6.78 | 1.38 ±<br>1.83 | 4.13 ±<br>6.26 | 1.86 ±<br>2.16 |
| (DiDP) | P = 0.02 |  |  |  |  | P = 0.47 |  |  | P = 0.29 |  |
| MOP | 0.81 ±<br>0.66 | NA | 1.81 ±<br>6.58 | **4.95 ±**<br>**4.59\*** | NA | 1.72 ±<br>2.80 | 4.32 ±<br>9.96 | 0.22 ±<br>0.28 | 2.4 ±<br>6.94 | 0.26 ±<br>0.34 |
| (DnOP) | P = 0.64 |  |  |  |  | P = 0.33 |  |  | P = 0.22 |  |
| MCPP | 1.8 ±<br>1.15 | NA | 2.34 ±<br>1.70 | 18.64 ±<br>20.42 | **0.33 ±**<br>**0.33\*** | 5.09 ±<br>10.09 | 5.65 ±<br>10.21 | 1.64 ±<br>0.99 | 4.31 ±<br>8.69 | 1.92 ±<br>1.29 |
| (DnOP) | P = 0.06 |  |  |  |  | P = 0.18 |  |  | P = 0.15 |  |
| MiDP | NA | NA | 0.42 ±<br>0.52 | 1.46 ±<br>2.04 | NA | NA | 1.49 ±<br>1.99 | 4.00 ±<br>0.53 | 0.77 ±<br>1.13 | NA |
| (DiDP) | P = 0.34 |  |  |  |  | P = 0.32 |  |  | NA |  |
| Low Molecular Weight |  |  |  |  |  |  |  |  |  |  |
| MEP | 86.52 ±<br>72.05 | 369.71<br>± 51.77 | 26.46 ±<br>39.39 | 349.49 ±<br>278.94 | 101.75 ±<br>68.24 | 42.38 ±<br>59.19 | 51.67 ±<br>106.53 | 43.26 ±<br>88.33 | 43.10 ±<br>81.50 | 54.10 ±<br>112.00 |
| (DEP) | P = 0.63 |  |  |  |  | P = 0.70 |  |  | P = 0.53 |  |
| MiBP | 14.59 ±<br>20.34 | 27.43 ±<br>12.24 | 32.39 ±<br>37.13 | 80.27 ±<br>105.52 | 17.85 ±<br>0.21 | 37.87 ±<br>26.56 | 43.66 ±<br>59.05 | 23.69 ±<br>26.05 | 35.30 ±<br>43.00 | 21.00 ±<br>27.00 |
| (DiBP) | P = 0.05 |  |  |  |  | P = 0.04 |  |  | P = 0.03 |  |
| MnBP | 41.95 ±<br>36.30 | 137.02<br>± 80.33 | 61.25 ±<br>61.31 | 160.48 ±<br>120.93 | 28.35 ±<br>9.40 | 69.73 ±<br>69.73 | 69.37 ±<br>64.52 | 60.11 ±<br>65.35 | 64.40 ±<br>65.10 | 65.10 ±<br>67.40 |
| (DBP, BBP) | P <0.01 |  |  |  |  | P = 0.59 |  |  | P = 0.87 |  |
| MBzP | 3.11 ±<br>1.68 | 3.2 ±<br>1.70 | 4.32 ±<br>7.80 | 81.06 ±<br>132.12 | 3.95 ±<br>0.35 | 18.49 ±<br>60.69 | 3.92 ±<br>3.98 | 6.99 ±<br>28.96 | 6.11 ±<br>24.30 | 11.00 ±<br>43.00 |
| (DBP, BBP) | P = 0.01 |  |  |  |  | P = 0.36 |  |  | P = 0.47 |  |

Note: The table above displays the number of observations, the mean ± SD, and p-value for each categorical analysis.
\* The data obtained originated from a single subset.
\*\* ANOVA was performed to analyze these strata
\*\*\*Note: T-testing was performed in place of ANOVA for higher accuracy
\*\*\*\*n = Number of observations; NA = Not Available; SD = Standard Deviation; EPA = Easter and Pacific Asia; MESA = Middle East and Southern Asia; P = P-value

When a phthalate monoester’s concentrations were reported for both adult and child age groups, concentrations were consistently higher among children (MEHP: 12.51 ng/mL vs. 9.11 ng/mL; MEOHP: 34.13 ng/mL vs. 19.64 ng/mL; MECPP: 52.43 ng/mL vs. 19.59 ng/mL; MCMHP: 17.25 ng/mL vs. 7.65 ng/mL; OH-MiNP: 7.27 ng/mL vs. 1.24 ng/mL; MiBP: 43.66 ng/mL vs. 23.69 ng/mL). Among the four phthalate monoesters that had significantly different mean concentrations between pregnant and non-pregnant groups, mean concentrations were lower among cohorts of pregnant individuals compared to the non-pregnant individuals (MEOHP: 18.30 ng/mL vs. 9.71 ng/mL; MECPP: 34.10 ng/mL vs. 20.30 ng/mL; MiNP: 1.88 ng/mL vs. 0.50 ng/mL; and MiBP: 35.30 ng/mL vs. 21.00 ng/mL).

Results of regression analyses including studies from all countries are presented in Table 3. In the covariate-adjusted linear regression analysis, we observed a significant increase of 4.29 ng/mL/year (95% CI: 2.44, 6.14) in MnBP concentration and a decrease of −0.89 ng/mL/year (95% CI: −1.57, −0.22) in MEOHP concentration over time. When we performed the weighted covariate-adjusted meta-regression analysis, there were significant non-linear associations between time and MnBP (beta: 0.59 ng/mL/year^2^, 95% CI: 0.31, 0.87), MEHHP (beta: 0.10 ng/mL/year^2^, 95% CI: 0.01, 0.20), MCMHP (beta: 0.33 ng/mL/year^2^, 95% CI: 0.10, 0.56), MiNP (beta: 0.03 ng/mL/year^2^, 95% CI: 0.01, 0.05), and MCPP (beta: −0.02 ng/mL/year^2^, 95% CI: −0.04, −0.002).We also observed significant negative non-linear associations between time and Additionally, we observed significant linear declines in MEOHP (beta: −0.60 ng/mL/year, 95% CI: −1.05, −0.16), MBzP (beta: −0.28 ng/mL/year, 95% CI: −0.46, −0.10), and MEP (beta: −1.23 ng/mL/year, 95% CI: −2.32, −0.15) concentrations over time.

**Table 3:**
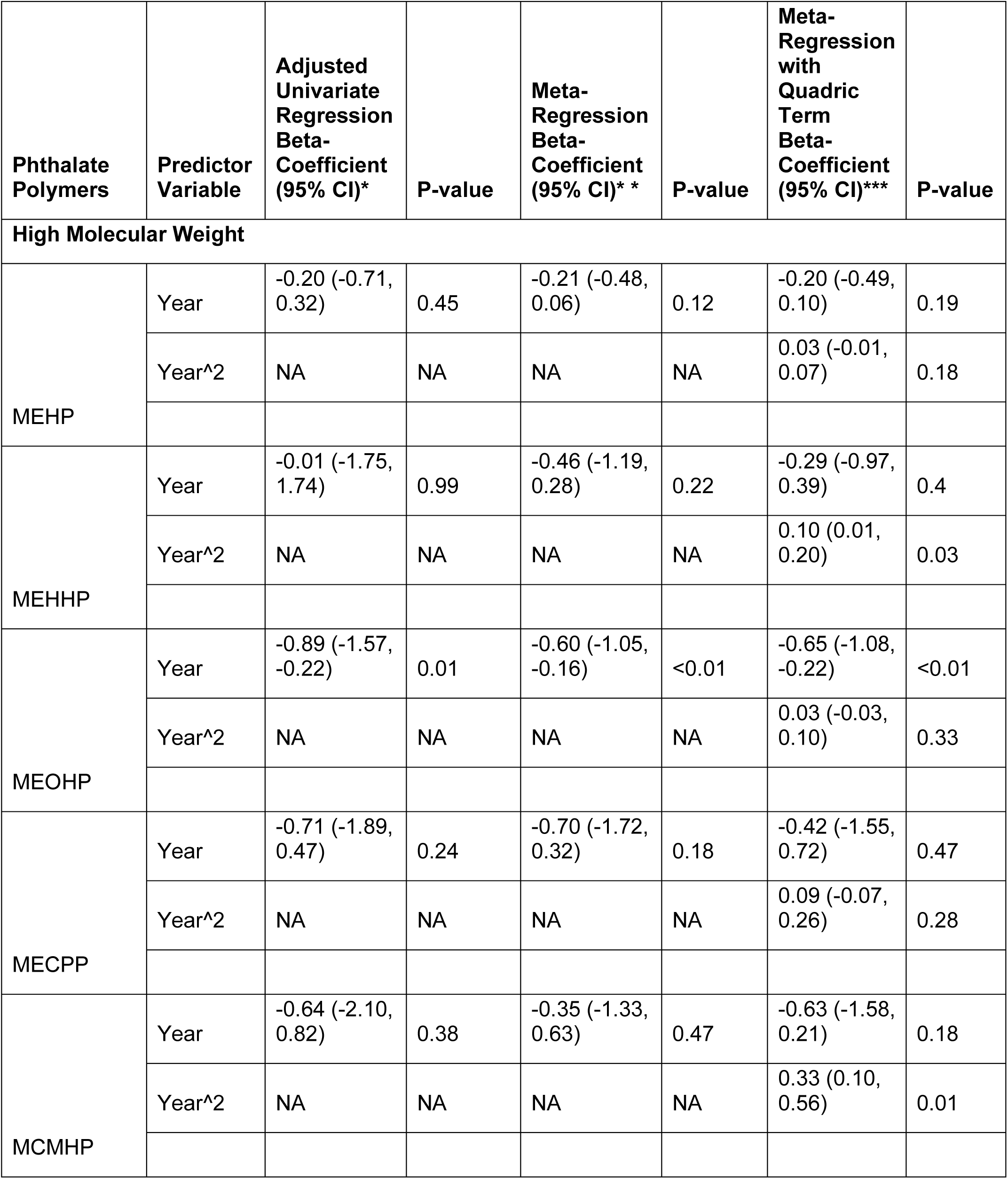
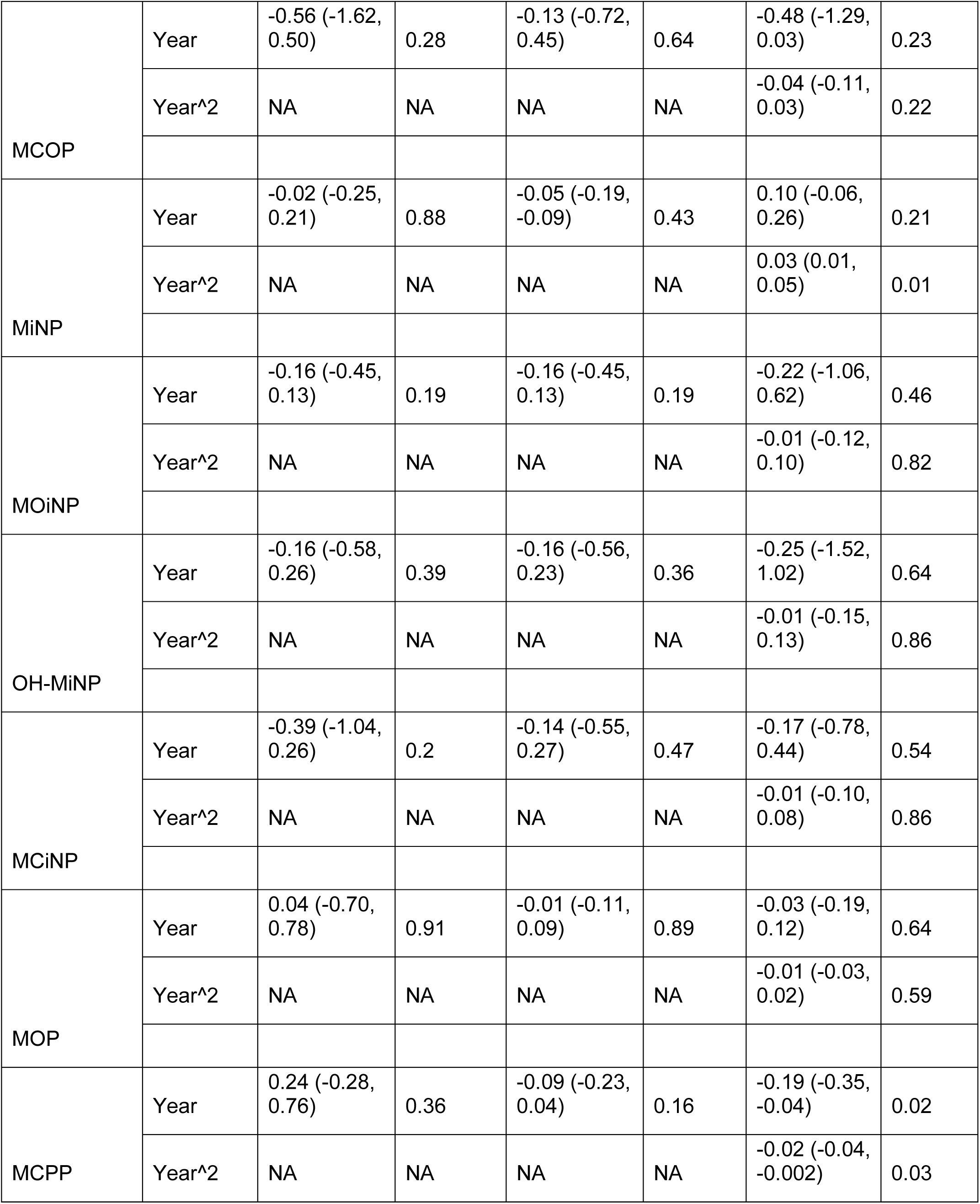
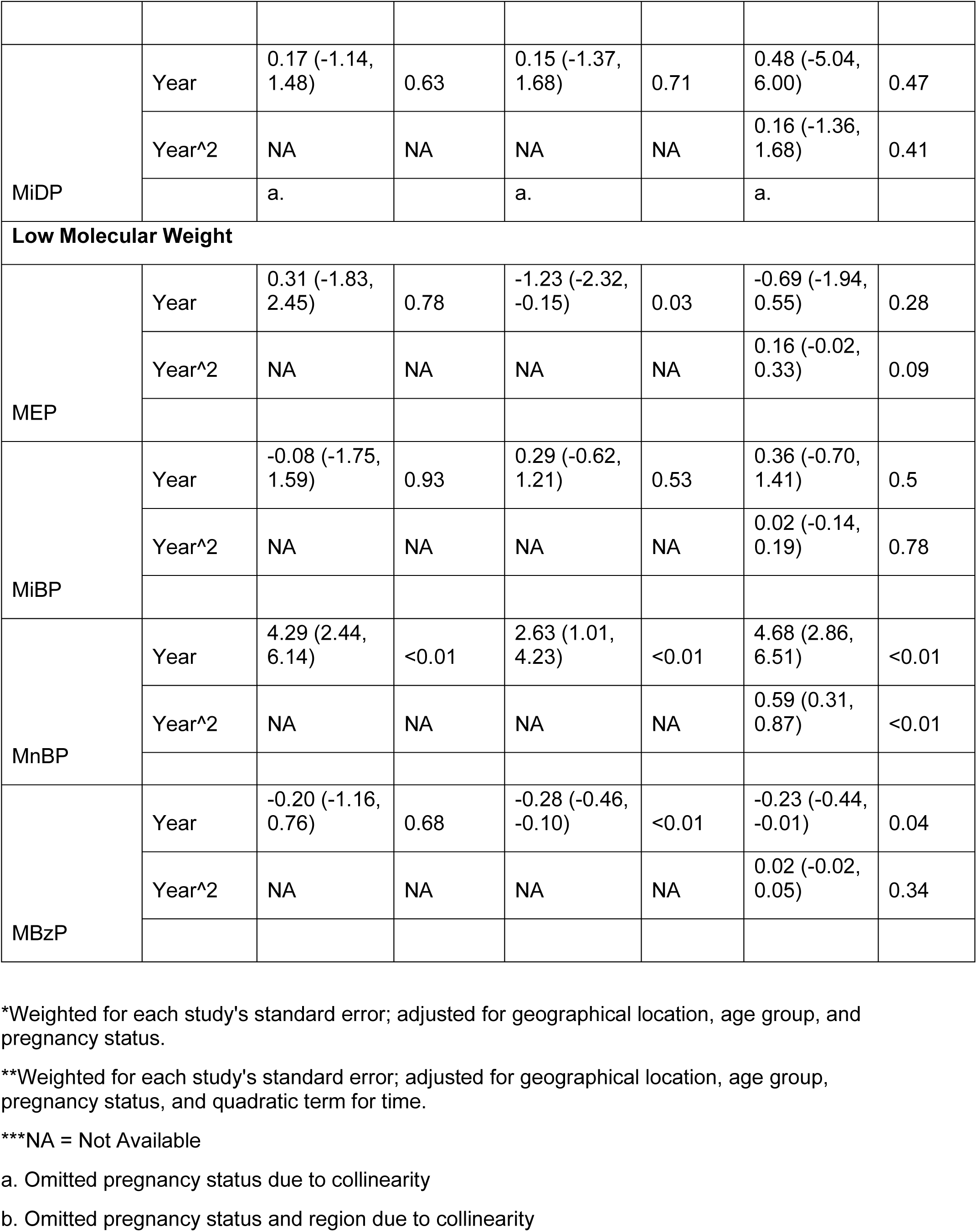
Adjusted Linear and Meta-Regressions for each monoester and year of sampling.

When we stratified and ran linear and quadratic meta-regression models by region (Table 4), many regions had insufficient information to provide a statistical finding. Of the regions that had sufficient data, we observed significant linear declines in MEOHP (beta: −0.80 ng/mL/year, 95% CI: [−1.17, −0.42]), MCPP (beta: −0.22 ng/mL/year, 95% CI: [−0.41, −0.03]), and MBzP (beta: −0.33 ng/mL/year, 95% CI: [−0.50, −0.15]) concentrations over time in the EPA region, and declines in MEP concentrations in Latin America (beta: −10.62 ng/ml/year; 95% CI: [−21.03, −0.21]) and Africa (beta: −8.98 ng/ml/year; 95% CI: [−9.69, −8.26]).

**Table 4:**
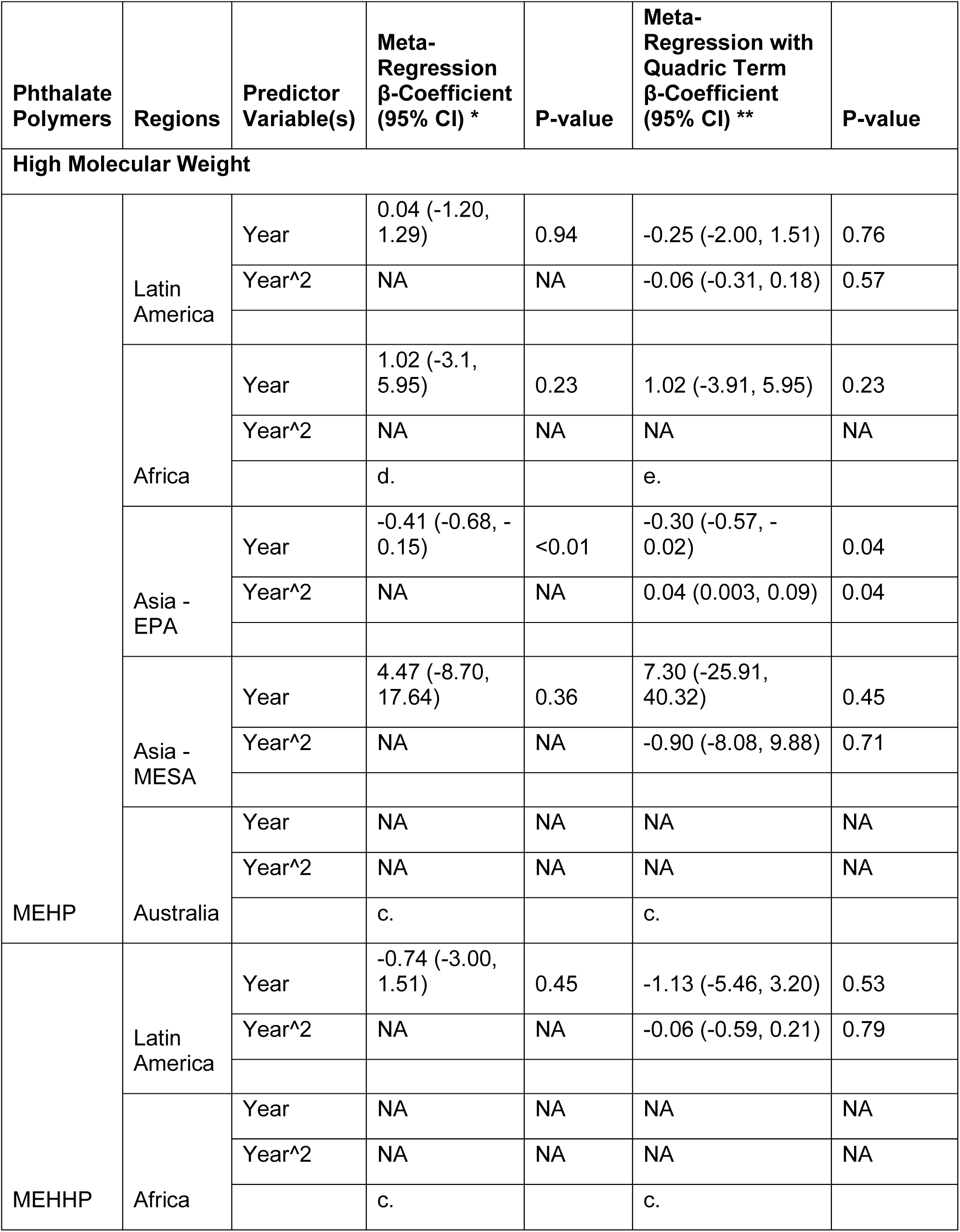
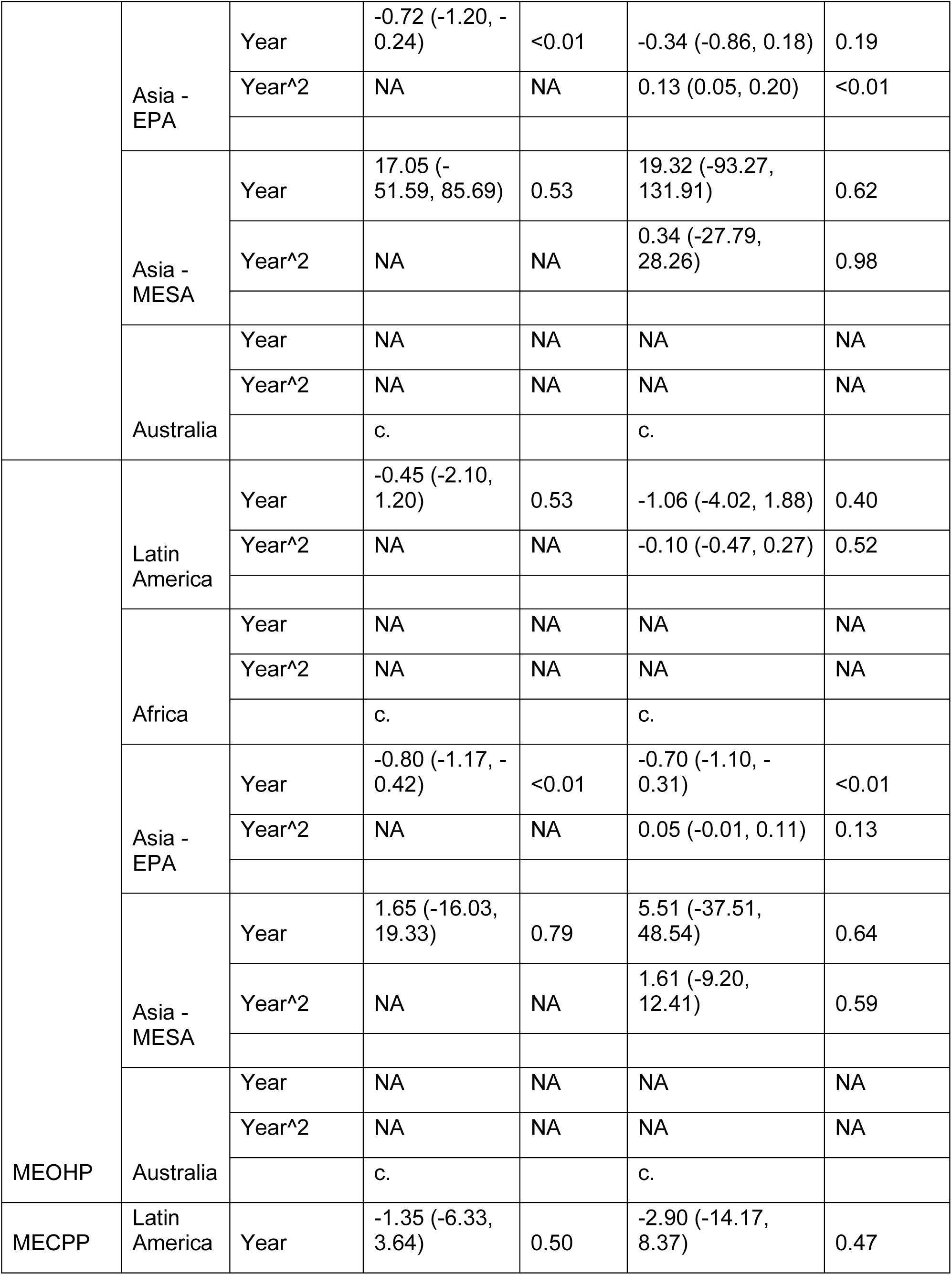
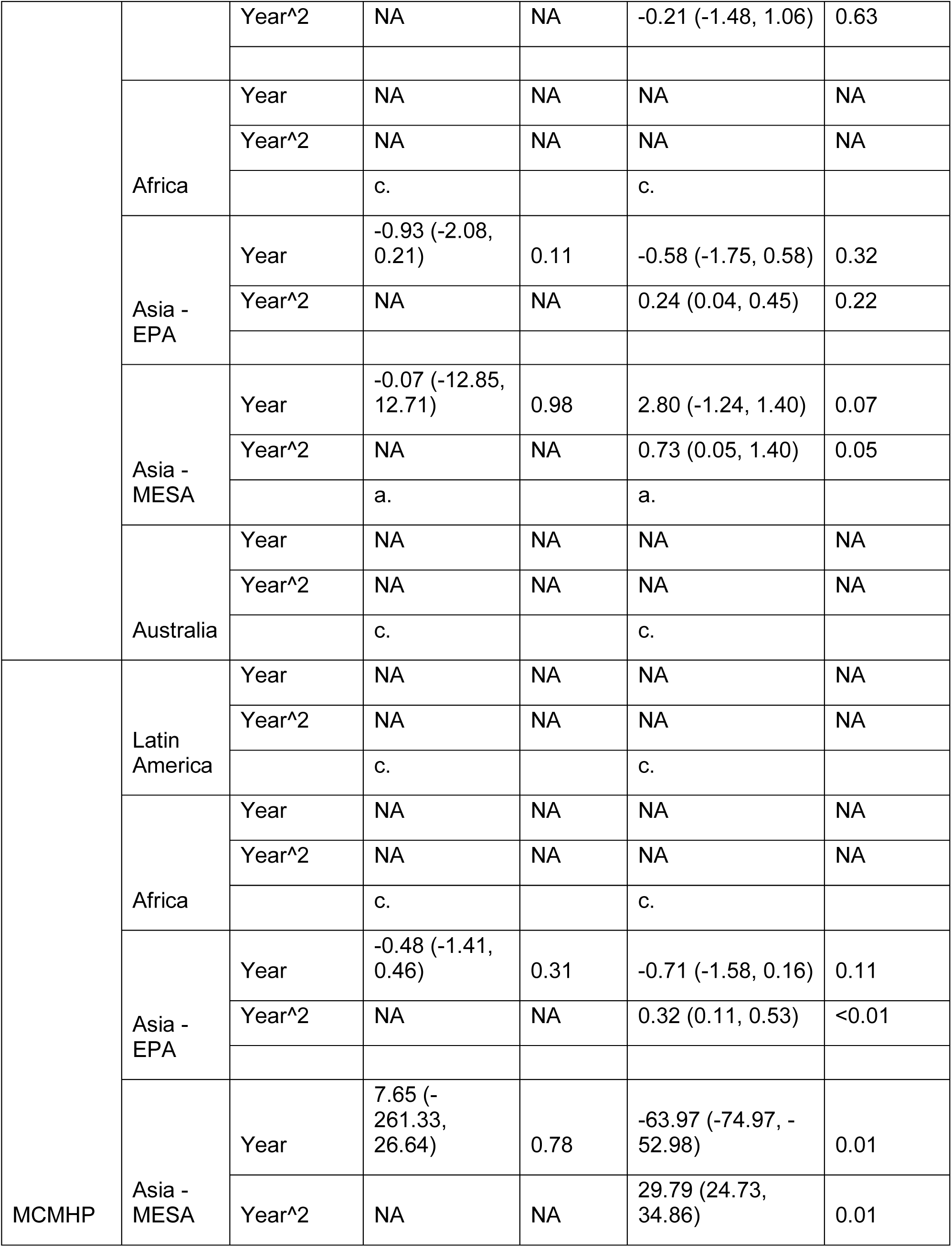
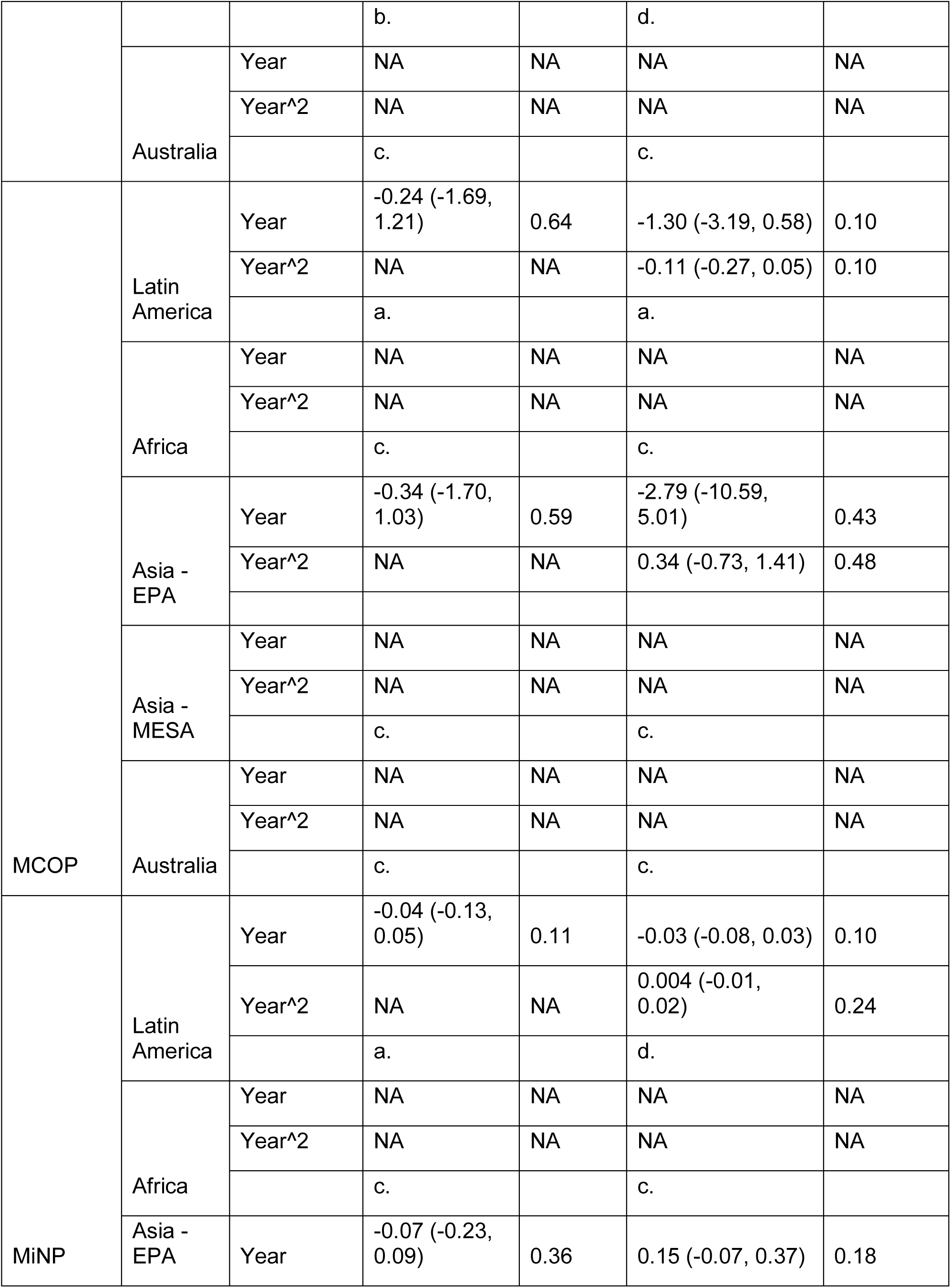
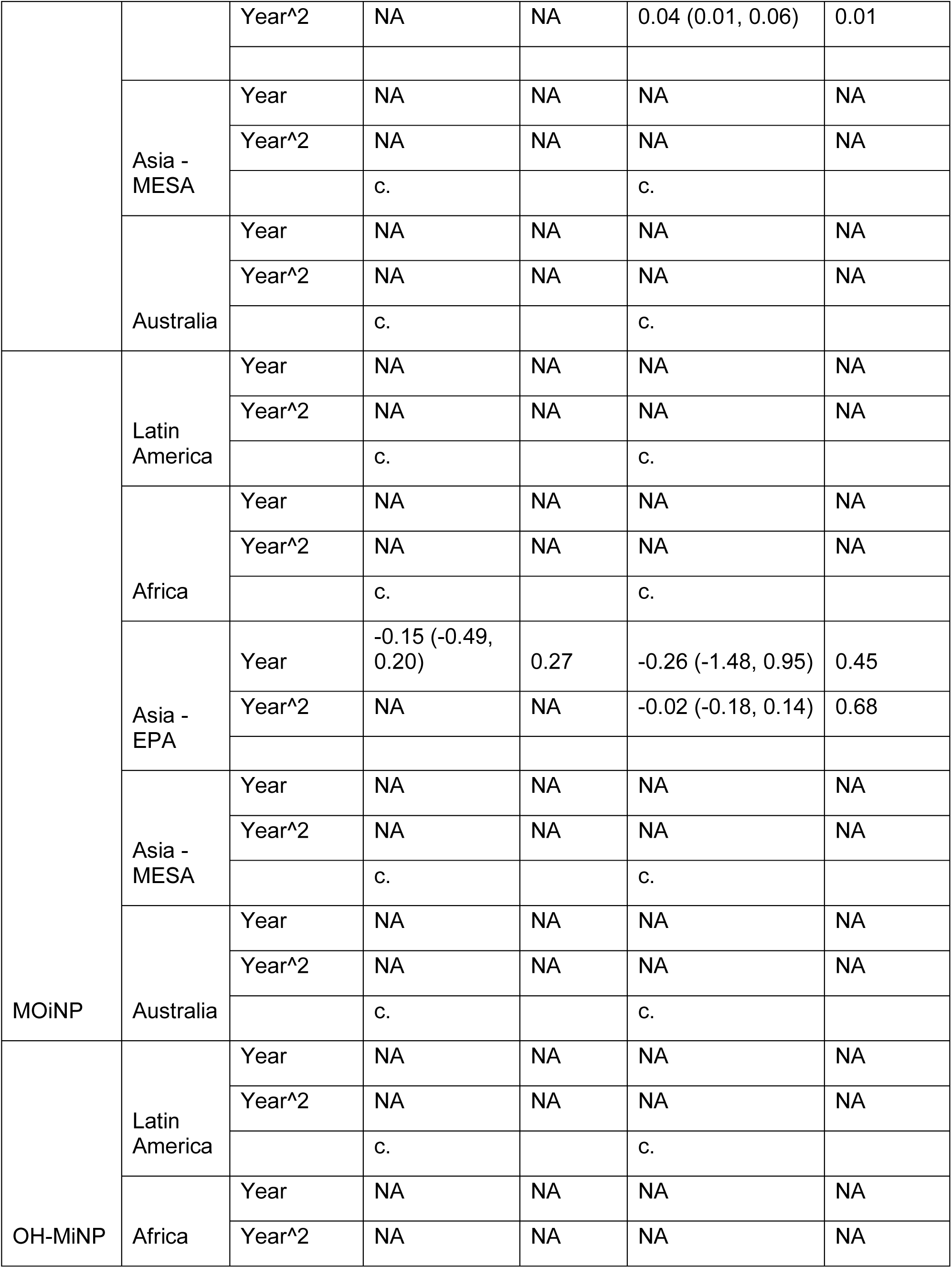
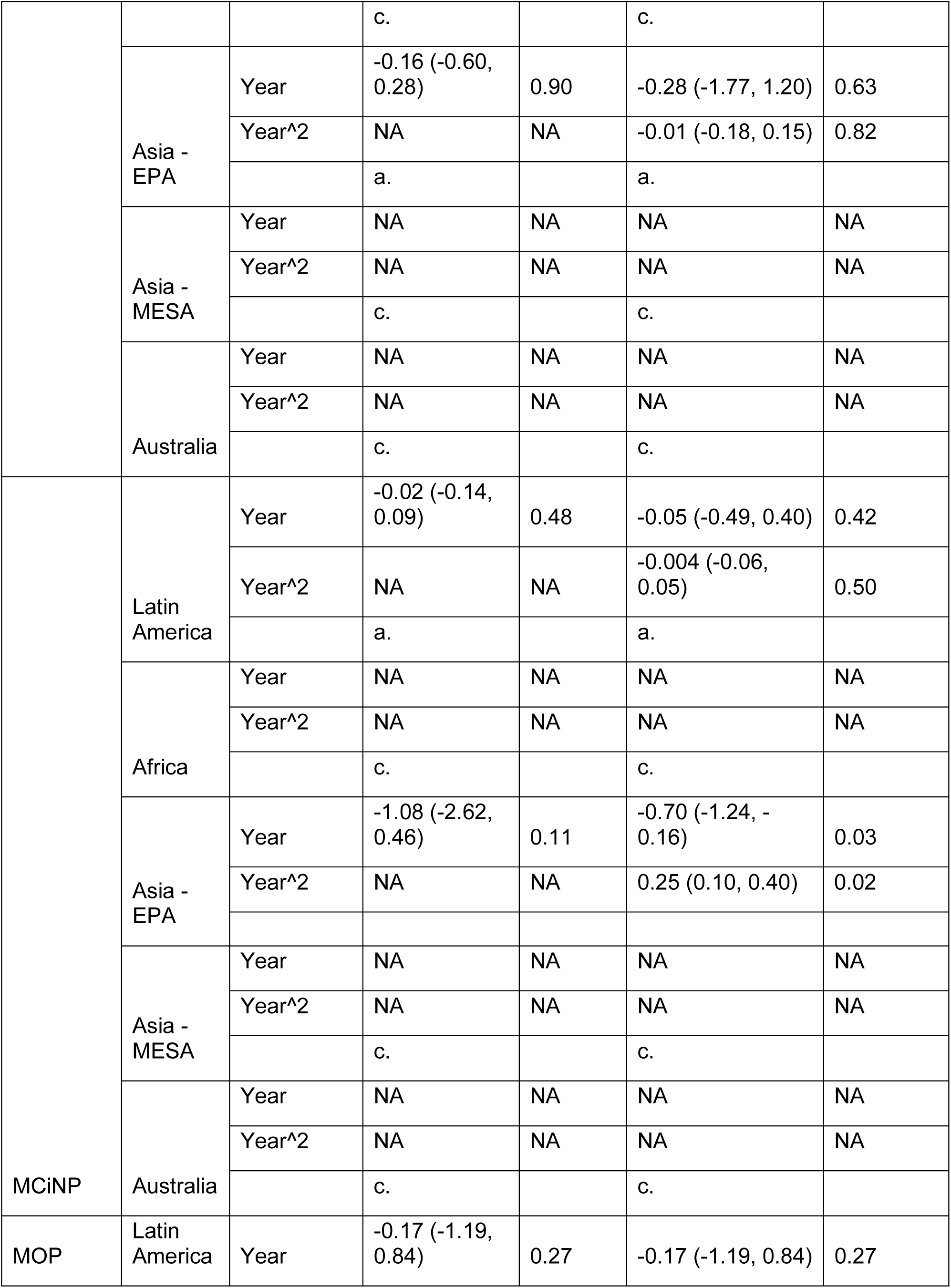
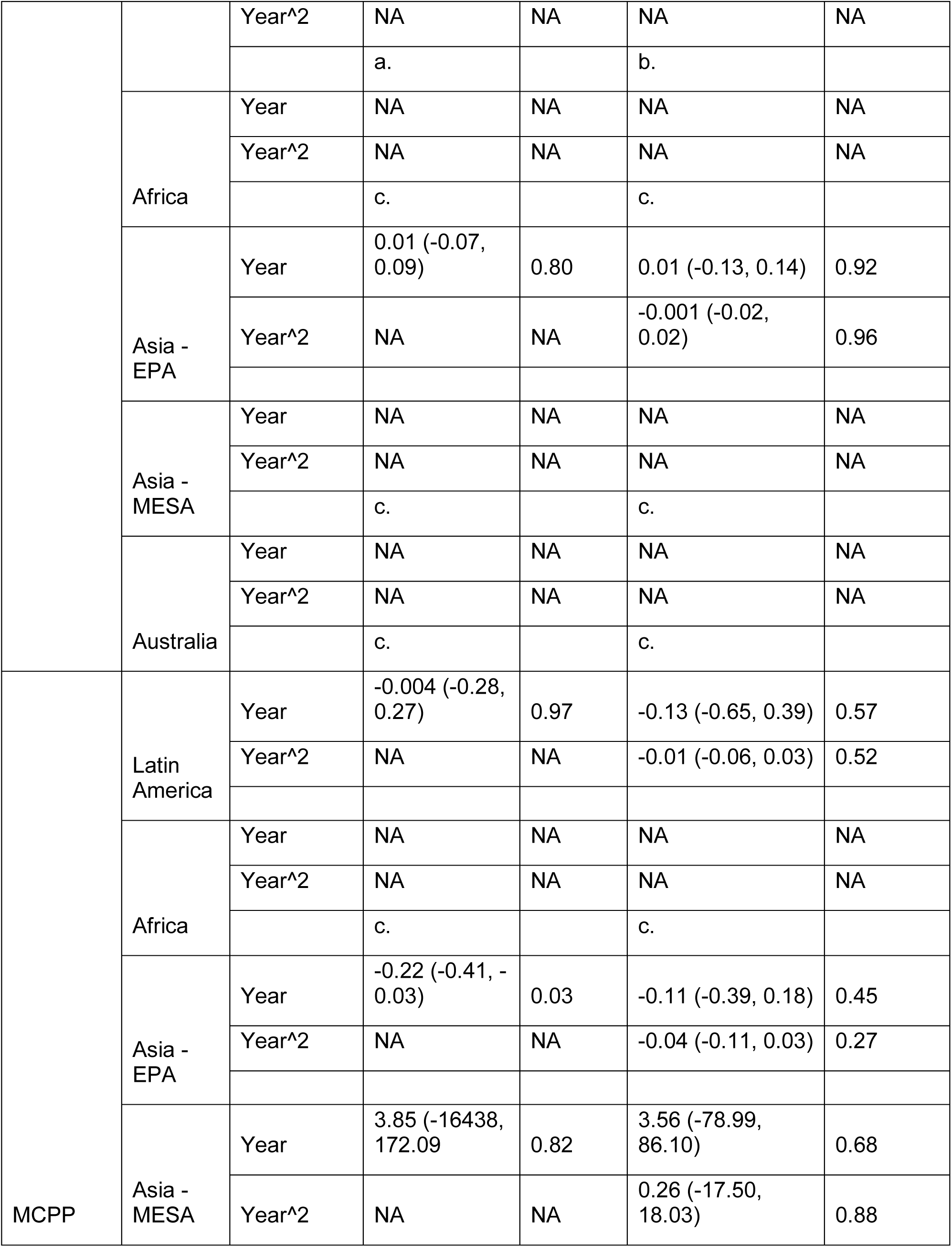
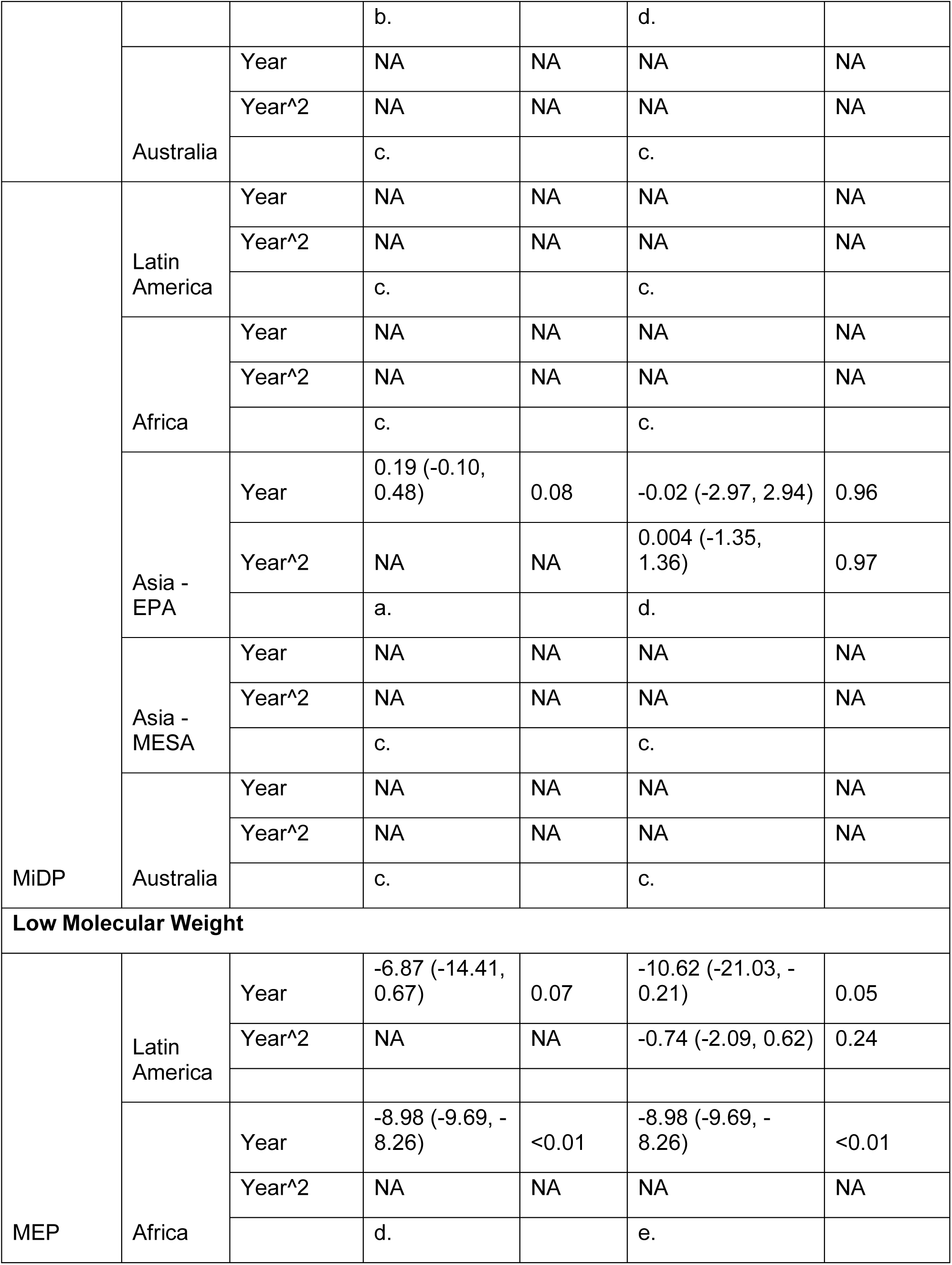
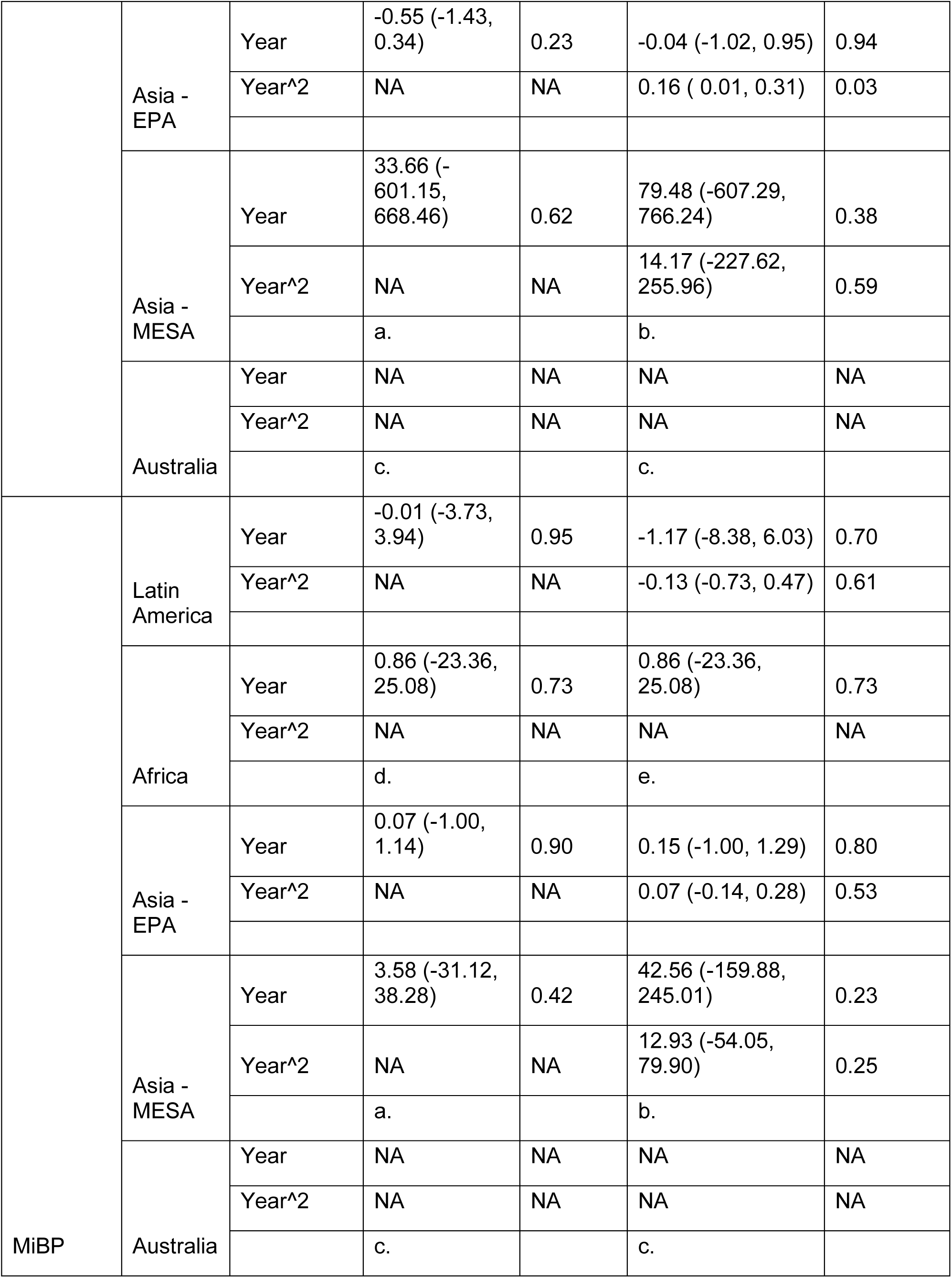
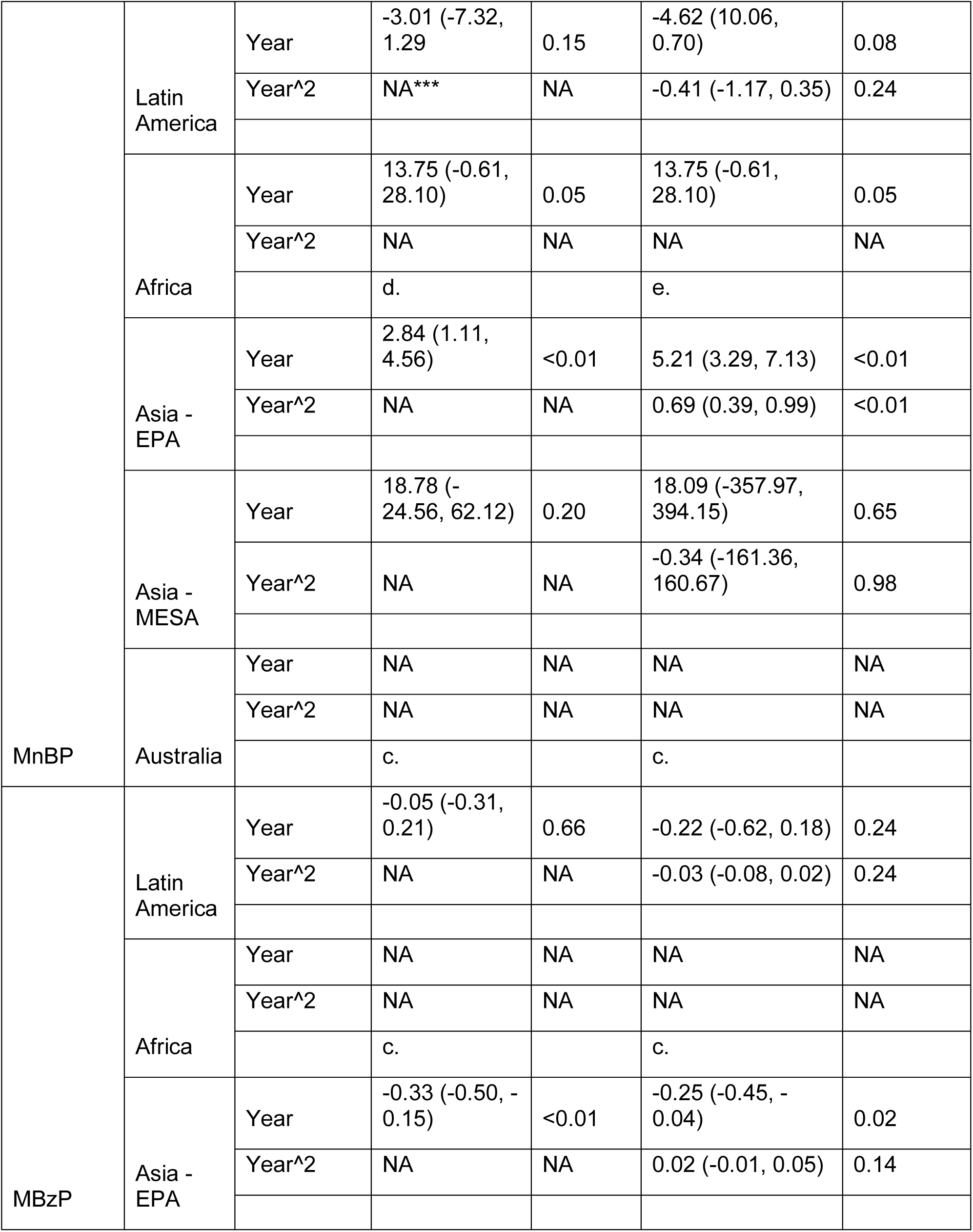
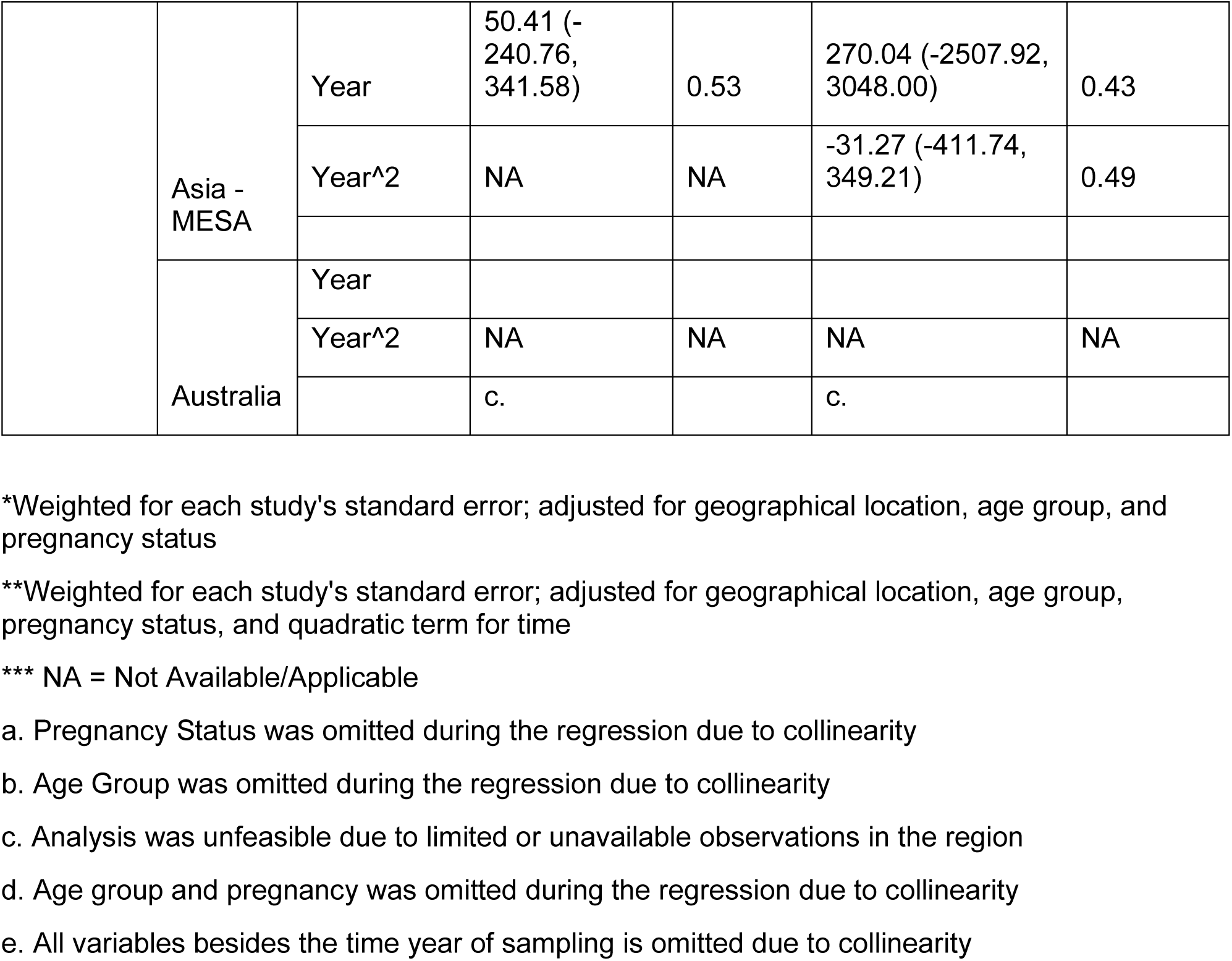
Meta-regressions between monoesters and year of sampling stratified for geographical regions.

We also observed significant non-linear associations between time and MnBP (beta: 0.69 ng/mL/year^2^, 95% CI: 0.39, 0.99), MEHP (beta: 0.04 ng/mL/year^2^, 95% CI: 0.003, 0.09), MEHHP (beta: 0.13 ng/mL/year^2^, 95% CI: 0.05, 0.20), MCMHP (beta: 0.32 ng/mL/year^2^, 95% CI: 0.11, 0.53), MiNP concentration (beta: 0.04 ng/mL/year^2^, 95% CI: 0.01, 0.06), MCiNP (beta: 0.25 ng/mL/year^2^, 95% CI: 0.10, 0.40), and MEP (beta: 0.16 ng/mL/year^2^, 95% CI: 0.01, 0.31) concentrations in the EPA region. Additionally, we observed significant non-linear associations between time and MECPP concentration (beta: 0.73 ng/mL/year^2^, 95% CI: [0.05, 1.40]) and MCMHP concentration (beta: 29.79 ng/ml/year^2^; 95% CI: [24.73, 34.86]) in the MESA region.

### 3.3 Tests for Heterogeneity

Overall, we detected high levels of heterogeneity across the study, as indicated by the large Cochran’s Q and I^2^ values (Supplement Table 4). By implementing the trim-and-fill method, we counteracted heterogeneity within each model’s pooled monoester phthalate concentration. However, several regions did not alter after the trim-and-fill method was implemented due to either insufficient data or low degrees of heterogeneity (Supplement Table 4).

### 3.4 Predicted Exposure Trends 2003 to 2023

Supplement Tables 5 and 6 indicate each phthalate monoester’s estimated predicted mean concentration at 5-year intervals based on linear and quadratic models and grouped by parent compound. Supplement Table 7 shows the calculation of the accompanying SDs. In summary, nearly all the phthalate monoesters besides MnBP, MiDP, and MiBP displayed declining linear concentration levels in combined and region-specific analyses from 2003 to 2023. In the quadratic models, most exhibited a U-shaped trend over the years, while MOiNP, OH-MiNP, MCOP, MCPP, and MOP exhibited an inverse U-shaped trend over the study period in both combined and region-specific analyses. Further visual aids and percentiles (10th, 25th, 50th, 75th, and 95th) for each year can be located in Figure 2, Supplement Tables 5 and 6, and Supplement Figures 1-5.

**Figure 2:**
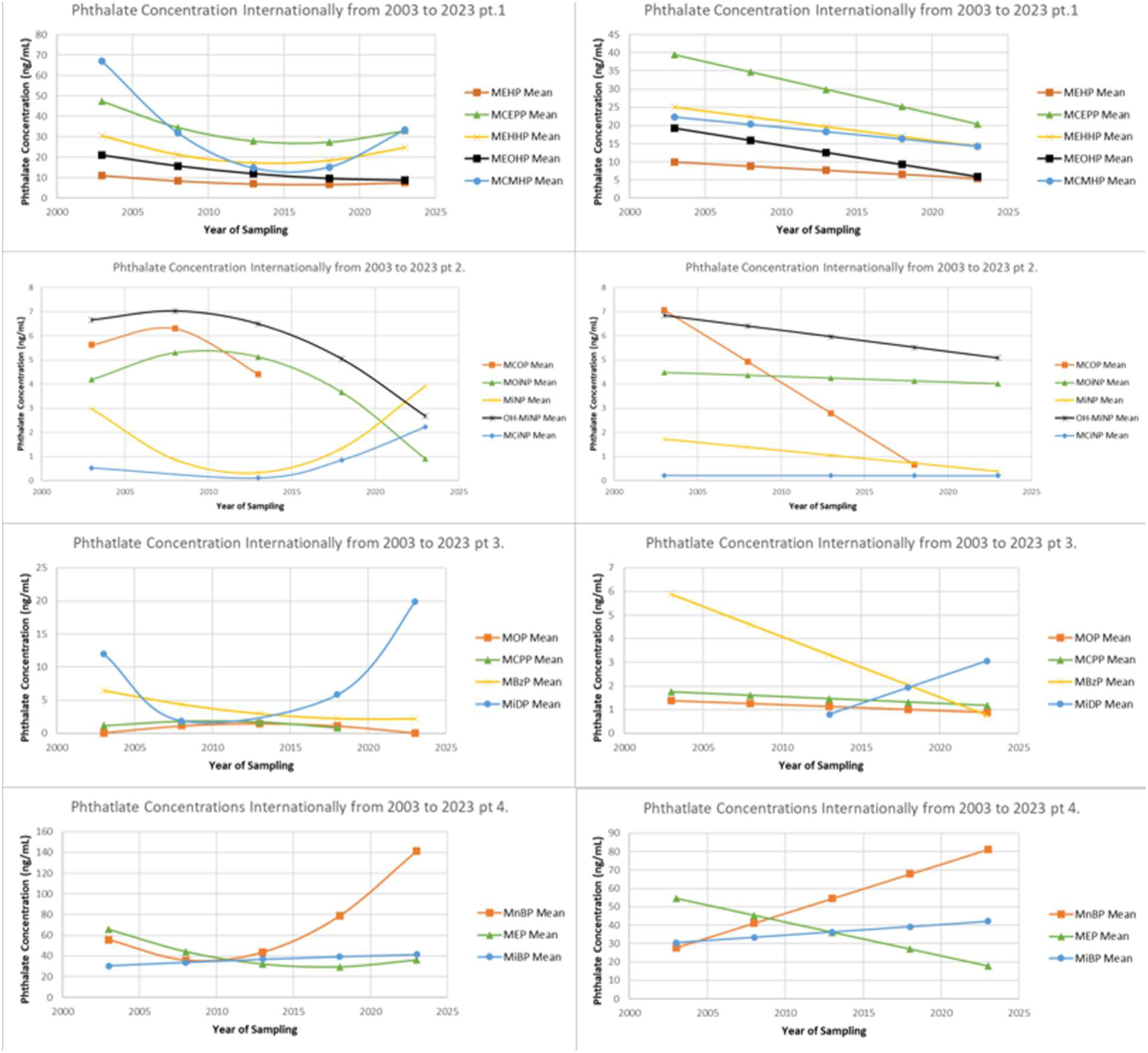
Linear and non-linear associations between year of sampling and each phthalate monoester, grouped by parent compound.

### 3.5 Sensitivity Meta-Analysis

When we excluded the articles that used GM values, we identified most beta-coefficients to have shifted by 15% or more compared to the overall regression outcome. Along with the shifts of the beta-coefficients, approximately half of the previously significant associations were no longer significant, and several novel associations became significant. We observed significant non-linear associations between time and MBzP concentration (beta: 0.04 ng/mL/year^2^; 95% CI: 0.01, 0.08) internationally, MBzP concentration (beta: 0.04 ng/mL/year^2^; 95% CI: 0.01, 0.08) in the EPA region, and MnBP concentration (beta: −31.16 ng/mL/year^2^; 95% CI: −36.57, −25.76) in the MESA region. Furthermore, we observed significant linear decreases in MOiNP (beta: −1.77 ng/mL/year; 95% CI: −2.25, −1.28) and OH-MiNP concentration (beta: −1.10 ng/mL/year; 95% CI: −1.89, −0.30) over time internationally and in the EPA region. Lastly, we observed significant linear increases in MiDP concentration (beta: 1.41 ng/mL/year; 95% CI: 1.00, 1.81) internationally and in MEP concentration (beta: 95.53 ng/mL/year; 95% CI: 51.29, 139.76) in MESA over time.

Similarly, when excluding articles with AM values, we observed a 15% or greater shift in beta-coefficient values, with many of the previously significant associations no longer significant. The exclusion of AM values also introduced several novel significant associations that weren’t present previously. We observed a non-linear association between time and MCOP concentration in EPA (beta: 2.13 ng/mL/year^2^, 95% CI: [1.16, 3.10]). Additionally, we observed significant negative linear associations between time and monoester concentration internationally (MECPP: −1.17 ng/mL/year, 95% CI: [−2.19, −0.15]; MCMHP: −0.70 ng/mL/year, 95% CI: [−1.39, −0.01]; OH-MiNP: −0.67 ng/mL/year, 95% CI: [−1.24, −0.09]) and in the EPA region (MECPP: −1.37 ng/mL/year, 95% CI: [−2.57, −0.16]; MCiNP: −0.58 ng/mL/year^2^, 95% CI: [−1.15, −0.01]).

Overall, the significance between time and each phthalate monoester varied depending on the use of GM and AM values. These sensitivity analysis outcomes warrant caution when substituting or combining results, as they could yield significant differences in their results. Additional details on significance levels and altered coefficient values are located in Supplement Tables 8 and 9.

## Discussion

### 4.1 Key Findings

This systematic review is among the first to examine urinary phthalate metabolite concentration trends in non-occupationally exposed populations outside the US, Europe, and Canada. Throughout the study period, we identified significant linear declines in many monoester concentrations across all regions combined (MEOHP, MBzP, and MEP) and in Latin America (MEP), Africa (MEP), and EPA (MEOHP, MCPP and MBzP) regions. When analyzing the quadratic models, we observed significant non-linear associations between time and many monoesters internationally (MnBP, MEHHP, MCMHP, MiNP, and MCPP), in the EPA region (MnBP, MEHP, MEHHP, MCMHP, MCiNP, MEP, and MiNP), and in the MESA region (MECPP and MCMHP). Lastly, we noticed consistent results for several monoesters and some new associations in a sensitivity analysis excluding studies that reported geometric rather than AM values

Prior studies have identified similar decreasing trends in phthalate over time within various regions (Domínguez-Romero et al., 2023; Lyu et al., 2022b). These studies attributed the observed declines to the regulation of DEHP, DiNP, and other phthalates. In 2003, the Ministry of Health in Japan prohibited the use of DEHP, DiNP, and other phthalates in food packaging and children’s toys (Hamamoto, 2002; Lyu et al., 2022a; Tsumura et al., 2003). China also restricted 16 phthalates in 2017 in food products and packaging and banned the export of children’s toys containing more than 1% by weight of DEHP, BBP, and Dibutyl phthalate (Wang and Qian, 2021). Along with China, in the early 2000’s, the Ministry of Health in some Latin American countries banned the manufacture, import, export, and any other trade or charitable use of toys, flexible chewable baby merchandise, or plastic childcare products containing greater than 0.1% of various phthalates (Tsang, 2008; Wu et al., 2020b). By implementing these regulations and bans on specific phthalates, such as DEHP and DiNP, population exposure to these EDCs will continue to decrease over time.

Several prior studies have also identified significant shifts within the plasticizer market, particularly in the substitution of phthalates (Lessmann et al., 2019; Silva et al., 2019; Zota Ami et al., 2014). DEHP, known for being the most used plasticizer globally, is being increasingly replaced by other phthalates such as DiNP and DiDP, which now represent approximately 30-60% of the US and Europe’s total plasticizer market. With alternative phthalates and substitutions, the concentration of DEHP secondary metabolites such as MEHP, MEOHP, and others has diminished throughout the years (Zota Ami et al., 2014). A prior study performed in Mexico showed similar results between DEHP and di(2-ethylhexyl) terephthalate (DEHTP), a well-documented alternative to DEHP. The study identified DEHTP to significantly increase metabolite concentration in the study’s participants, while DEHP metabolite concentration remained stagnant between 2007 and 2010 (Wu et al., 2020b).

Another study performed in China also displayed a similar outcome, suggesting that the substitution of phthalates may have influenced the results. However, instead of DEHP declining, DEP and DiBP phthalates, also prominent phthalates being used in industry practices, were shown to have a significant decrease, while DEHP increased between 2010 and 2019 (Domínguez-Romero et al., 2023). These changes in phthalates use seen in industry practices caused by consumer demands or regulations on chemical safety may seem to have some benefit; they simultaneously alter human exposure to other variations, which may increase the risk of different health outcomes.  

Previous studies have also identified increasing temporal trends for a few phthalates across different regions. These studies identified dietary consumption, especially of shellfish and seafood, and food packaging as primary sources, as phthalates leach out of plastic containers and wraps, thereby contaminating the item (Alak et al., 2024; Kang et al., 2023; Tian et al., 2022). Furthermore, the uses of agriculture films and pesticides have been identified as contributing to phthalate exposure. Prior studies revealed that agriculture films, a standard tool used to protect crops, contain 10 to 60% plasticizers by weight, added to extend their longevity in the fields (Ding et al., 2021; Zhou et al., 2020). Another study on pesticides analyzed the formula of 110 pesticides used in agriculture and found that phthalates accounted for 2 to 40% of the total organic solvent content (Tang et al., 2020b). Fertilizers were also noted as playing a critical role in amplifying these exposure rates. They are often packaged with high concentrations of plasticizers and create chemical deposits in the soil that accumulate over time (Li et al., 2023d; Mo et al., 2008; Zhang et al., 2015). These plastic additives contaminate soil, crops, and water sources, indirectly heightening the general public’s exposure to plasticizers (Li et al., 2023d).

Cosmetics and hygiene products have also emerged as a contributing factors for dermal phthalate exposure (Koniecki et al., 2011). Products like body wash, deodorant, shampoos, lotions, and oils, are infused with phthalate compounds to prolong fragrance longevity, have been associated with a 1.17 to 2.86 increase dermal exposure to various phthalate metabolites (Bloom et al., 2024). A study analyzing feminine hygiene products reported that women who used products that contained DEP had 1.37 to 2.92 time the urinary MEP concentration of those who didn’t use these products. The study also reported that participants who used perfumes that contained DnBP had 1.38-fold higher urinary MnBP concentration compared with participants who didn’t use perfume (Parlett et al., 2013). These studies show that basic hygienic care products that are used daily can increase the risk of phthalate exposure without public recognition.

Lastly, contaminated water reservoirs and improper waste management have been identified as contributors to phthalate exposure, specifically in LMICs. In these countries, water reservoirs are often polluted with waste containing traces of phthalates and many other EDCs due to limited regulation concerning waste management, and clean water and lack of adequate (drinking) water treatment processes is often not available (Abtahi et al., 2019; Chen et al., 2023c; Dada and Ikeh, 2018; Joubert et al., 2019; Li et al., 2023d).

### 4.2 Limitations

Several limitations should be considered when interpreting our results. First, most of the studies in our overall analysis originate from high-income countries, particularly in the EPA. The high number of studies from high-income countries may skew the combined analysis since high-income countries will have more funding to support stronger regulation and waste-management systems than LMICs. Second, several monoesters had insufficient articles for regional analyses, especially in regions with a high proportion of LMICs, such as Africa and Latin America. With such low representation for these countries, the data on each phthalate metabolite concentration may be skewed toward values similar to those of more documented countries. Third, while there are hundreds of different phthalates metabolites that are being used, our review focuses primarily on the most prevalent metabolites seen in industry practices internationally. This does limit our understanding of potential temporal and geographic variation seen in different phthalate metabolites. Finally, substituting the GM for the monoester phthalate arithmetic means may have introduced calculation bias in our results.

### 4.3 Strengths

This review is one of the few to analyze temporal and geographical differences in concentrations of various phthalate metabolites, specifically in Latin America, Africa, and Australia. In addition, it is the first to examine differences in exposure levels by age and pregnancy status. Together, these strengths offer the opportunity to capture the general population’s exposure levels to parent compounds of the included phthalate monoesters.

Additionally, our review examined individual phthalate monoesters without linking them to their parent compounds. By doing so, it offers a strong approximation of which monoesters are increasing and diminishing throughout the time and region. In addition to the examination of each monoester, our review highlighted the lack of significant research in regions that include many LMICs. To the degree that data were available, the review provides insight into phthalate exposure in these regions, allowing for meaningful comparisons in future research with more extensively studied regions like the US and Europe. Finally, using the multi-reviewer approach throughout each stage of screening and data extraction processes minimized selection bias.

### 4.4 Future Research Directions and Implications

Research documenting the adverse health outcomes associated with phthalate exposure is prominent across the environmental and medical fields. However, over the past two decades, there have been fewer than five articles focused on phthalate exposure from Africa, a continent with 54 countries, and Australia, with a population of over 26 million. While many regions, particularly LMICs where populations are known to be disproportionately exposed, are understandably focusing their resources on environmental contamination caused by pesticides and heavy metals, these same populations are also highly exposed to plastic waste. This plastic waste contains phthalates among other chemicals, all of which warrant greater monitoring and research within these regions.

Additionally, with increased regulation of the use of certain phthalate parent compounds such as DEHP, stronger assessment of DEHTP and other substitutes for DEHP are warranted. Furthermore, an increase in longitudinal studies or repeated panel studies is also crucial to understanding exposure levels, since measuring the same population over time will offer the best opportunity to document the influence of industrialization and the shift in policies and regulations regarding EDC use within specific regions.

Finally, studies on the attributable disease burden and costs associated with phthalate exposure at regional and international levels are needed. As previously stated, phthalates are associated with a plethora of adverse health outcomes, such as cancer and cardiovascular morbidity. Understanding the financial impact of phthalate-attributed health outcomes is essential for highlighting the economic losses and evaluating the tradeoffs of ongoing plasticizer use.

## Conclusion

In conclusion, this review reveals the widespread prevalence of exposure to some of the phthalates used in industry practices. Between 2000 and 2023, many phthalates had non-linear increases globally and in the EPA region. In Latin America, MESA, and Africa, few phthalates displayed significant evidence of a decline in concentration throughout the years.

Our study shed some light on the shift of exposure trends throughout time and reveals gaps in biomonitoring data in various regions, particularly in regions with limited research infrastructure. Additional studies filling these gaps and estimating attributable disease burden and costs at regional and global levels are needed to show these EDCs’ impact on the public. In addition, increased discussion on regulations to lower or prevent the use and consumption of any phthalate form is necessary in all regions to counteract the continuous growth of phthalate exposure. This, in turn, will contribute to aiding in the protection of human health through advocacy for measures to eliminate exposure to harmful environmental chemicals.

## Data Availability

All data produced in the present study are available upon reasonable request to the authors.

## Abbreviations

ANOVA: Analysis of Variance
BBP: Benzyl butyl phthalate
CI: Confidence Interval
DEHP: Di(2-ethylhexyl) phthalate
DEHTP: di(2-ethylhexyl) terephthalate
DEP: Diethyl phthalate
DiBP: Diisobutyl Phthalate
DiDP: Diisodecyl Phthalate
DiNP: Diisononyl Phthalate
DBP: Dibutyl phthalate
DnOP: Di-n-octyl Phthalate
EDCs: Endocrine-Disrupting Chemicals
EPA: Eastern and Pacific Asia
IQR: Interquartile Range
LMICs: Low- and Middle-Income Countries
MESA: Middle East and South Asia
MnBP: Mono(2-carboxymethylhexyl) phthalate
MEHP: Mono(2-ethyl-5-carboxypentyl) phthalate
MEHHP: Mono(2-ethyl-5-hydroxyhexyl) phthalate
MEOHP: Mono(2-ethyl-5-oxohexyl) phthalate
MECPP: Mono(2-ethyl-5-carboxypentyl) phthalate
MCMHP: Mono(2-carboxymethylhexyl) phthalate
MCOP: Mono-carboxy-isooctyl phthalate
MiNP: Mono-isononyl phthalate
MOiNP: Mono-oxo-isononyl phthalate
OH-MiNP: Mono-hydroxy-isononyl phthalate
MCiNP: mono-carboxy-isononyl phthalate
MOP: Mono-octyl phthalate
MCPP: Mono(3-carboxypropyl) phthalate
MBzP: Monobenzyl phthalate
MiDP: Mono-isodecyl phthalate
MEP: Mono-ethyl phthalate
MiBP: Mono-isobutyl phthalate
MECPTP: Mono-2-ethyl-5-carboxypentyl terephthalate
PRISMA: Preferred Reporting Items for Systematic Reviews and Meta-Analysis
PROSPERO: International Prospective Register of Systematic Reviews
PVC: Polyvinyl Chloride
SD: Standard Deviation
US: United States

