## Supplement Tables and Figures for "Temporal and Geographic Variability of International Urinary Phthalates in Humans: A Systematic Review and Meta-Analysis of Biomonitoring Data"

Supplement Materials:

Table of Contents:

| Table/Figures: | **Description:** |
| --- | --- |
| S. Table 1 | This table displays a breakdown of each parent compound into their monoester form |
| S. Table 2 | This displays the creatinine concentration at each age group and overall. |
| S. Table 3 | This table is a descriptive breakdown of the subsets seen throughout the review. |
| S. Table 4 | This table displays the pooled mean, I^2, Q, and adjusted pool mean values for each phthalate monoester internationally and stratified per region. |
| S. Table 5 | This table displayed the estimated percentiles of the linear shift in each phthalate monoester concentration from 2003 to 2023. |
| S. Table 6 | This table displayed the estimated percentiles of the non-linear shift in each phthalate monoester concentration from 2003 to 2023. |
| S. Table 7 | This table displays the Standard Deviation regression used to calculate the standard deviation for each bisphenol for S. Table 5 and 6. |
| S. Table 8 | This table displays a sensitivity analysis excluding articles that provided Geometric Mean values. |
| S. Table 9 | This table displays a sensitivity analysis excluding articles that provided Arithmetic mean values. |
| S. Figure 1 | This figure displays the linear and non-linear shift of each monoester phthalate in Latin America. |
| S. Figure 2 | This figure displays the linear and non-linear shift of each monoester phthalate in Africa. |
| S. Figure 3 | This figure displays the linear and non-linear shift of each monoester phthalate in the Eastern and Pacific Asia region. |
| S. Figure 4 | This figure displays the linear and non-linear shift of each monoester phthalate in the Middle East and Southern Asia regions. |
| S. Figure 5 | This figure displays the linear and non-linear shift of each monoester phthalate in Australia. |

Supplement Table 1: Phthalates of interest.

| **Parent Compound** | **Abbreviation of Parent Metabolites** | **Primary Metabolite** | **Abbreviation of Primary Metabolites** |
| --- | --- | --- | --- |
| **High Molecular Weight** | | | |
| Di(2-ethylhexyl) phthalate | DEHP | Mono(2-ethylhexyl) phthalate | MEHP |
|  |  | Mono(2-ethyl-5-hydroxyhexyl) phthalate | MEHHP |
|  |  | Mono(2-ethyl-5-oxohexyl) phthalate | MEOHP |
|  |  | Mono(2-ethyl-5-carboxypentyl) phthalate | MECPP |
|  |  | Mono(2-carboxymethylhexyl) phthalate | MCMHP |
| Diisononyl Phthalate | DiNP | Mono-carboxy-isooctyl phthalate | MCOP |
|  |  | Mono-isononyl phthalate | MiNP |
|  |  | Mono-oxo-isononyl phthalate | MOiNP |
|  |  | Mono-hydroxy-isononyl phthalate | OH-MiNP |
| Di-n-octyl phthalate | DnOP | Mono-octyl phthalate | MOP |
|  |  | Mono(3-carboxypropyl) phthalate | MCPP |
| Diisodecyl phthalate | DiDP | Mono-isodecyl phthalate | MiDP |
|  |  | Mono(carboxy-isononyl) phthalates | MCiNP |
| **Low Molecular Weight** | | | |
| Diethyl phthalate | DEP | Mono-ethyl phthalate | MEP |
| Diisobutyl phthalate | DiBP | Mono-isobutyl phthalate | MiBP |
| Benzyl butyl phthalate | BBP | Mono-n-butyl phthalate | MnBP |
| Dibutyl phthalate | DBP | Monobenzyl phthalate | MBzP |

Supplement Table 2: Creatinine-levels across the lifespan.

| **Age group (Years)** | **Creatinine concentration (mg/mL)** |
| --- | --- |
| Total | 0.91 |
| 0-2 | 0.52 |
| 3 - 6 | 0.74 |
| 7 - 12 | 0.9 |
| 13 - 18 | 1.44 |
| 19 - 29 | 1.25 |
| 30 - 39 | 0.94 |
| 40 - 49 | 0.93 |
| 50 - 59 | 0.85 |
| >=60 | 0.81 |

Note: These values used to adjust for creatinine-adjusted phthalate concentration are based on the conversions seen by Park, J. H. et al.

Supplement Table 3: Summary Descriptions of the Included Phthalate Articles.

| **Phthalate** | **# of Articles *** | **Total Sub-Cohorts *** | **Total Sample Size *** | **Sampling Period **** | **# of Sub-Cohorts that used AM Values** | **# of Sub-Cohorts that used GM Values** | **# of Sub-Cohorts that used Median Values** | **Regions** | | | | | **Life Cycle Stage **** | | | **Pregnant ***** | |
| --- | --- | --- | --- | --- | --- | --- | --- | --- | --- | --- | --- | --- | --- | --- | --- | --- | --- |
|  |  |  |  |  |  |  |  | **Latin America** | **Africa** | **Asia - EPA** | **Asia - MESA** | **Australia** | **Mixed** | **Youth (< 18 years)** | **Adult (18+ years)** | **No** | **Yes** |
| **High Molecular Weight** | | | | | | | | | | | | | | | | | |
| **MEHP (n)** | 172 | 201 | 77726 | 2000-2022 | 60 | 97 | 44 | 14 | 3 | 175 | 8 | 1 | 27 | 62 | 112 | 148 | 53 |
| **%** |  |  |  |  | 29.85 | 48.26 | 21.89 | 6.97 | 1.49 | 87.06 | 3.98 | 0.5 | 13.43 | 30.85 | 55.72 | 73.63 | 26.37 |
| **MEHHP (n)** | 190 | 215 | 100391 | 2000-2022 | 68 | 106 | 41 | 11 | 2 | 191 | 9 | 2 | 25 | 62 | 128 | 166 | 49 |
| **%** |  |  |  |  | 31.63 | 49.30 | 19.07 | 5.12 | 0.93 | 88.84 | 4.19 | 0.92 | 11.63 | 28.84 | 59.53 | 77.21 | 22.79 |
| **MEOHP (n)** | 185 | 216 | 99635 | 2000-2022 | 65 | 107 | 44 | 11 | 2 | 193 | 8 | 2 | 29 | 65 | 122 | 167 | 49 |
| **%** |  |  |  |  | 30.09 | 49.54 | 20.37 | 5.09 | 0.93 | 89.35 | 3.7 | 0.93 | 13.43 | 30.09 | 56.48 | 77.31 | 22.69 |
| **MECPP (n)** | 109 | 125 | 57699 | 2000-2022 | 31 | 68 | 26 | 9 | 2 | 106 | 6 | 2 | 14 | 36 | 75 | 93 | 32 |
| **%** |  |  |  |  | 24.80 | 54.40 | 20.80 | 7.2 | 1.6 | 84.8 | 4.8 | 1.6 | 11.2 | 28.8 | 60 | 74.4 | 25.6 |
| **MCMHP (n)** | 43 | 50 | 20734 | 2009-2022 | 12 | 26 | 12 | 2 | 0 | 44 | 4 | 0 | 6 | 13 | 31 | 41 | 9 |
| **%** |  |  |  |  | 24.00 | 52.00 | 24.00 | 4 | 0 | 88 | 8 | 0 | 12 | 26 | 62 | 82 | 18 |
| **MCOP (n)** | 18 | 24 | 13503 | 2002-2019 | 6 | 13 | 5 | 7 | 2 | 14 | 1 | 0 | 3 | 11 | 10 | 19 | 5 |
| **%** |  |  |  |  | 25.00 | 54.17 | 20.83 | 29.17 | 8.33 | 58.33 | 4.17 | 0 | 12.5 | 45.83 | 41.67 | 79.17 | 20.83 |
| **MiNP (n)** | 29 | 34 | 11858 | 2003-2020 | 9 | 17 | 8 | 4 | 0 | 29 | 1 | 0 | 4 | 29 | 1 | 27 | 7 |
| **%** |  |  |  |  | 26.47 | 50.00 | 23.53 | 11.76 | 0 | 85.29 | 2.94 | 0 | 11.76 | 85.29 | 2.94 | 79.41 | 20.59 |
| **MOiNP (n)** | 5 | 9 | 3677 | 2002-2018 | 1 | 6 | 2 | 0 | 0 | 7 | 2 | 0 | 0 | 5 | 4 | 6 | 3 |
| **%** |  |  |  |  | 11.11 | 66.67 | 22.22 | 0 | 0 | 77.78 | 22.22 | 0 | 0 | 55.56 | 44.44 | 66.67 | 33.33 |
| **OH-MiNP (n)** | 6 | 11 | 4130 | 2002-2018 | 1 | 8 | 2 | 0 | 0 | 9 | 2 | 0 | 0 | 9 | 2 | 6 | 5 |
| **%** |  |  |  |  | 9.09 | 72.73 | 18.18 | 0 | 0 | 81.81 | 18.18 | 0 | 0 | 81.82 | 18.18 | 54.55 | 45.45 |
| **MCiNP** | 11 | 17 | 7478 | 2002-2018 | 3 | 11 | 3 | 6 | 2 | 7 | 2 | 0 | 1 | 10 | 6 | 13 | 4 |
| **%** |  |  |  |  | 17.65 | 64.71 | 17.65 | 35.29 | 11.76 | 41.12 | 11.76 | 0 | 5.88 | 58.82 | 35.29 | 76.47 | 23.53 |
| **MOP (n)** | 22 | 24 | 8381 | 2005-2021 | 8 | 10 | 6 | 4 | 0 | 19 | 1 | 0 | 3 | 8 | 13 | 17 | 7 |
| **%** |  |  |  |  | 33.33 | 41.67 | 25.00 | 16.67 | 0 | 79.17 | 4.17 | 0 | 12.5 | 33.33 | 54.17 | 70.83 | 29.17 |
| **MCPP (n)** | 34 | 42 | 17336 | 2000-2020 | 12 | 25 | 5 | 12 | 0 | 25 | 4 | 1 | 11 | 12 | 19 | 31 | 11 |
| **%** |  |  |  |  | 28.57 | 59.52 | 11.90 | 28.57 | 0 | 59.52 | 9.52 | 2.38 | 26.19 | 28.57 | 45.24 | 73.81 | 26.19 |
| **MiDP (n)** | 6 | 6 | 2511 | 2010-2017 | 3 | 3 | 0 | 0 | 0 | 4 | 2 | 0 | 0 | 2 | 4 | 6 | 0 |
| **%** |  |  |  |  | 50.00 | 50.00 | 0.00 | 0 | 0 | 66.67 | 33.33 | 0 | 0 | 33.33 | 66.67 | 100 | 0 |
| **Low Molecular Weight** | | | | | | | | | | | | | | | | | |
| **MEP (n)** | 160 | 184 | 78221 | 2000-2021 | 60 | 86 | 38 | 14 | 3 | 160 | 5 | 2 | 24 | 53 | 107 | 134 | 50 |
| **%** |  |  |  |  | 32.61 | 46.74 | 20.65 | 7.61 | 1.63 | 86.96 | 2.72 | 1.09 | 13.04 | 28.8 | 58.15 | 72.83 | 27.17 |
| **MiBP (n)** | 115 | 135 | 52295 | 2000-2021 | 40 | 62 | 33 | 12 | 3 | 113 | 5 | 2 | 21 | 43 | 71 | 106 | 29 |
| **%** |  |  |  |  | 29.63 | 45.93 | 24.44 | 8.89 | 2.22 | 83.7 | 3.7 | 1.48 | 15.56 | 31.85 | 52.59 | 78.52 | 21.48 |
| **MnBP (n)** | 185 | 216 | 93634 | 2000 - 2021 | 64 | 101 | 51 | 13 | 3 | 191 | 7 | 2 | 26 | 65 | 125 | 167 | 49 |
| **%** |  |  |  |  | 29.63 | 46.76 | 23.61 | 6.02 | 1.39 | 88.43 | 3.24 | 0.93 | 12.04 | 30.09 | 57.87 | 77.31 | 22.69 |
| **MBzP (n)** | 146 | 168 | 81187 | 2000-2021 | 47 | 81 | 40 | 14 | 2 | 143 | 7 | 2 | 19 | 49 | 100 | 124 | 44 |
| **%** |  |  |  |  | 27.98 | 48.21 | 23.81 | 8.33 | 1.19 | 85.12 | 4.17 | 1.19 | 11.31 | 29.17 | 59.52 | 73.81 | 26.19 |

* These variables present the sum values.

**Note: There were no documented elderly (age > 65) cohort across the monoester subsets

*** The timeframe is recorded as the range during which samples were collected for each monoester.

a. %: Percentage of the total observations per monoester; n: the sum of data points for that category.

b. Due to the use of diverse cohorts in studies, with documentation for each separately, observations were recorded for each categorical variable.

Supplement Table 4: Statistical Pooled Means, Heterogeneity, and Imputed Articles for Phthalate Using the Trim-and-Fill Method.

| **Phthalate Polymers** | **Regions** | **Unadjusted Pooled Mean (95% CI)** | **Number of Imputed Articles** | **Adjusted Pooled Mean (95% CI)** | **Heterogeneity** |
| --- | --- | --- | --- | --- | --- |
| **High Molecular Weight** | | | | | |
| MEHP | Global | 7.67 (7.35, 8.00) | 6 | 8.43 (7.97, 8.89) | Q = 3.5e+06 |
|  |  |  |  |  | I^2 = 100 |
|  | Latin America | 6.19 (5.23, 7.15) | 4 | 9.18 (5.40, 12.97) | Q = 66783.36 |
|  |  |  |  |  | I^2 = 99.98 |
|  | Africa | 7.62 (4.05, 11.19) | 0 | 7.62 (4.05, 11.19) | Q = 5.53 |
|  |  |  |  |  | I^2 = 63.84 |
|  | Asia - EPA | 7.61 (7.25, 7.96) | 5 | 8.09 (7.66, 8.53) | Q = 3.3e+06 |
|  |  |  |  |  | I^2 = 99.99 |
|  | Asia - MESA | 48.49 (37.73, 59.25) | 1 | 36.79 (26.29, 47.28) | Q = 1495.59 |
|  |  |  |  |  | I^2 = 99.53 |
|  | Australia | 4.00 (3.99, 4.01) | 0 | 4.00 (3.99, 4.01) | Q = NA |
|  |  |  |  |  | I^2 = NA |
| MEHHP | Global | 18.25 (17.47, 19.04) | 14 | 19.75 (18.88, 20.63) | Q = 1.7e+07 |
|  |  |  |  |  | I^2 = 100 |
|  | Latin America | 21.19 (13.05, 29.33) | 1 | 22.92 (11.93, 33.91) | Q = 55876.12 |
|  |  |  |  |  | I^2 = 99.98 |
|  | Africa | 36.70 (19.86, 53.53) | 0 | 36.70 (19.86, 53.53) | Q = 2.22 |
|  |  |  |  |  | I^2 = 54.97 |
|  | Asia - EPA | 17.39 (16.57, 18.21) | 14 | 18.81 (17.91, 19.71) | Q = 1.6e+07 |
|  |  |  |  |  | I^2 = 100 |
|  | Asia - MESA | 95.09 (77.82, 112.36) | 3 | 189.47 (94.98, 283.96) | Q = 2943.24 |
|  |  |  |  |  | I^2 = 99.73 |
|  | Australia | 13.44 (5.71, 14.14) | 0 | 13.44 (5.71, 14.14) | Q = 733.68 |
|  |  |  |  |  | I^2 = 99.86 |
| MEOHP | Global | 13.02 (12.41, 13.64) | 20 | 14.89 (14.03, 15.74) | Q = 7.2e+06 |
|  |  |  |  |  | I^2 = 100 |
|  | Latin America | 14.31 (9.4, 19.22) | 2 | 16.83 (9.39, 24.27) | Q = 46550.78 |
|  |  |  |  |  | I^2 = 99.98 |
|  | Africa | 25.96 (16.01, 35.90) | 0 | 25.96 (16.01, 35.90) | Q = 1.56 |
|  |  |  |  |  | I^2 = 36.02 |
|  | Asia - EPA | 12.57 (11.92, 13.22) | 23 | 14.49 (13.60, 15.38) | Q = 7.1e+06 |
|  |  |  |  |  | I^2 = 100 |
|  | Asia - MESA | 48.75 (39.31, 58.19) | 3 | 88.54 (36.54, 140.54) | Q = 968.45 |
|  |  |  |  |  | I^2 = 99.28 |
|  | Australia | 10.60 (5.89, 12.30) | 1 | 8.20 (4.25, 12.05) | Q = 551.59 |
|  |  |  |  |  | I^2 = 99.82 |
| MECPP | Global | 25.74 (24.49, 26.98) | 6 | 27.66 (26.26, 29.06) | Q = 3.9e+06 |
|  |  |  |  |  | I^2 = 100 |
|  | Latin America | 41.05 (26.29, 55.82) | 2 | 49.98 (27.06, 72.91) | Q = 31532.56 |
|  |  |  |  |  | I^2 = 100 |
|  | Africa | 86.56 (63.76, 109.35) | 1 | 84.67 (65.29, 104.05) | Q = 0.04 |
|  |  |  |  |  | I^2 = 0.00 |
|  | Asia - EPA | 24.32 (22.96, 25.68) | 9 | 26.93 (25.45, 28.41) | Q = 3.8e+06 |
|  |  |  |  |  | I^2 = 100 |
|  | Asia - MESA | 36.97 (23.15, 50.80) | 0 | 36.97 (23.15, 50.80) | Q = 566.91 |
|  |  |  |  |  | I^2 = 99.12 |
|  | Australia | 18.34 (7.27, 29.42) | 1 | 12.70 (3.66, 21.75) | Q = 765 |
|  |  |  |  |  | I^2 = 99.87 |
| MCMHP | Global | 8.85 (8.01, 9.69) | 0 | 7.86 (7.31, 8.41) | Q = 1.1e+06 |
|  |  |  |  |  | I^2 = 100 |
|  | Latin America | 26.33 (24.91, 27.75) | 1 | 26.30 (24.91, 27.69) | Q = 0.03 |
|  |  |  |  |  | I^2 = 0.00 |
|  | Africa | NA | NA | NA | Q = NA |
|  |  |  |  |  | I^2 = NA |
|  | Asia - EPA | 8.05 (7.17, 8.93) | 2 | 9.19 (7.33, 11.04) | Q = 1.1e+06 |
|  |  |  |  |  | I^2 = 100 |
|  | Asia - MESA | 17.45 (11.56, 23.34) | 2 | 4.61 (ND, 11.70) | Q = 175.49 |
|  |  |  |  |  | I^2 = 98.29 |
|  | Australia | NA | NA | NA | Q = NA |
|  |  |  |  |  | I^2 = NA |
| MCOP | Global | 2.93 (2.71, 3.15) | 3 | 3.81 (3.23, 4.40) | Q = 2.1e+06 |
|  |  |  |  |  | I^2 = 100 |
|  | Latin America | 5.40 (2.33, 8.47) | 0 | 5.40 (2.33, 8.47) | Q = 9742.48 |
|  |  |  |  |  | I^2 = 99.99 |
|  | Africa | 4.00 (2.55, 5.45) | 1 | 3.50 (2.06, 4.94) | Q = 1.75 |
|  |  |  |  |  | I^2 = 42.97 |
|  | Asia - EPA | 1.49 (1.16, 1.72) | 0 | 1.49 (1.16, 1.72) | Q = 2.0e+06 |
|  |  |  |  |  | I^2 = 100 |
|  | Asia - MESA | 2.30 (1.39, 3.21) * | NA | 2.30 (1.386, 3.21) * | Q = NA |
|  |  |  |  |  | I^2 = NA |
|  | Australia | NA | NA | NA | Q = NA |
|  |  |  |  |  | I^2 = NA |
| MiNP | Global | 1.02 (0.94, 1.10) | 1 | 1.31 (0.99, 1.63) | Q = 5.3e+05 |
|  |  |  |  |  | I^2 = 100 |
|  | Latin America | 0.43 (0.32, 0.53) | 0 | 0.43 (0.32, 0.53) | Q = 1737.82 |
|  |  |  |  |  | I^2 = 99.83 |
|  | Africa | NA | NA | NA | Q = NA |
|  |  |  |  |  | I^2 = NA |
|  | Asia - EPA | 1.13 (1.04, 1.22) | 1 | 1.46 (1.07, 1.85) | Q = 5.1e+05 |
|  |  |  |  |  | I^2 = 99.99 |
|  | Asia - MESA | 4.96 (0.80, 9.13) * | NA | 4.96 (0.80, 9.13) * | Q = NA |
|  |  |  |  |  | I^2 = NA |
|  | Australia | NA | NA | NA | Q = NA |
|  |  |  |  |  | I^2 = NA |
| MOiNP | Global | 1.87 (1.66, 2.07) | 4 | 0.82 (0.62, 1.01) | Q = 8352.97 |
|  |  |  |  |  | I^2 = 99.90 |
|  | Latin America | NA | NA | NA | Q = NA |
|  |  |  |  |  | I^2 = NA |
|  | Africa | NA | NA | NA | Q = NA |
|  |  |  |  |  | I^2 = NA |
|  | Asia - EPA | 1.72 (1.50, 1.93) | 2 | 0.82 (0.61, 1.03) | Q = 8209.36 |
|  |  |  |  |  | I^2 = 99.93 |
|  | Asia - MESA | 6.06 (0.14, 11.99) | 1 | 3.15 (ND, 8.25) | Q = 23.62 |
|  |  |  |  |  | I^2 = 95.77 |
|  | Australia | NA | NA | NA | Q = NA |
|  |  |  |  |  | I^2 = NA |
| OH-MiNP | Global | 2.35 (2.06, 2.65) | 1 | 2.28 (1.99, 2.58) | Q = 7016.81 |
|  |  |  |  |  | I^2 = 99.86 |
|  | Latin America | NA | NA | NA | Q = NA |
|  |  |  |  |  | I^2 = NA |
|  | Africa | NA | NA | NA | Q = NA |
|  |  |  |  |  | I^2 = NA |
|  | Asia - EPA | 2.11 (1.8175, 2.41) |  | 2.11 (1.8175, 2.41) | Q = 6867.51 |
|  |  |  |  |  | I^2 = 99.88 |
|  | Asia - MESA | 9.48 (0.59, 18.37) | 1 | 5.12 (ND, 12.76) | Q = 22.00 |
|  |  |  |  |  | I^2 = 95.45 |
|  | Australia | NA | NA | NA | Q = NA |
|  |  |  |  |  | I^2 = NA |
| MCiNP | Global | 2.43 (2.25 2.62) | 1 | 2.63 (2.45, 2.81) | Q = 3.8e+06 |
|  |  |  |  |  | I^2 = 100 |
|  | Latin America | 0.82 (0.67, 0.97) | 2 | 0.68 (0.37, 1.00) | Q = 1.8e+05 |
|  |  |  |  |  | I^2 = 100 |
|  | Africa | 1.62 (1.01, 2.23) | 1 | 1.70 (1.18, 2.22) | Q = 0.1 |
|  |  |  |  |  | I^2 = 0.00 |
|  | Asia - EPA | 0.46 (0.44, 0.48) | 2 | 0.43 (0.40, 0.45) | Q =8966.97 |
|  |  |  |  |  | I^2 = 99.93 |
|  | Asia - MESA | 13.05 (ND, 28.64) | 1 | 21.00 (3.45, 38.53) | Q = 790.53 |
|  |  |  |  |  | I^2 = 99.87 |
|  | Australia | NA | NA | NA | Q = NA |
|  |  |  |  |  | I^2 = NA |
| MOP | Global | 0.46 (0.35, 0.57) | 2 | 0.57 (0.42, 0.73) | Q = 62887.76 |
|  |  |  |  |  | I^2 = 99.96 |
|  | Latin America | 0.81 (0.14, 1.48) | 0 | 0.81 (0.14, 1.48) | Q = 32983.48 |
|  |  |  |  |  | I^2 = 99.99 |
|  | Africa | NA | NA | NA | Q = NA |
|  |  |  |  |  | I^2 = NA |
|  | Asia - EPA | 0.31 (0.26, 0.36) | 4 | 0.39 (0.29, 0.49) | Q = 8400.00 |
|  |  |  |  |  | I^2 = 99.79 |
|  | Asia - MESA | 4.95 (4.16, 5.74) * | NA | 4.95 (4.16, 5.74) * | Q = NA |
|  |  |  |  |  | I^2 = NA |
|  | Australia | NA | NA | NA | Q = NA |
|  |  |  |  |  | I^2 = NA |
| MCPP | Global | 1.77 (1.66, 1.88) | 2 | 1.77 (1.66, 1.88) | Q = 3.7e+05 |
|  |  |  |  |  | I^2 = 100 |
|  | Latin America | 1.74 (1.05, 2.44) | 0 | 1.74 (1.05, 2.44) | Q =87557.62 |
|  |  |  |  |  | I^2 = 100 |
|  | Africa | NA | NA | NA | Q = NA |
|  |  |  |  |  | I^2 = NA |
|  | Asia - EPA | 1.88 (1.76, 2.01) | 1 | 1.88 (1.76, 2.01) | Q = 2.7e+05 |
|  |  |  |  |  | I^2 = 99.99 |
|  | Asia - MESA | 2.00 (0.42, 3.58) | 2 | 0.98 (ND, 3.04) | Q = 70.00 |
|  |  |  |  |  | I^2 = 95.71 |
|  | Australia | 0.33 (0.23, 0.43) | 0 | 0.33 (0.23, 0.43) | Q = NA |
|  |  |  |  |  | I^2 = NA |
| MiDP | Global | 0.68 (0.34, 1.02) | 0 | 0.68 (0.34, 1.02) | Q = 229.03 |
|  |  |  |  |  | I^2 = 97.82 |
|  | Latin America | NA | NA | NA | Q = NA |
|  |  |  |  |  | I^2 = NA |
|  | Africa | NA | NA | NA | Q = NA |
|  |  |  |  |  | I^2 = NA |
|  | Asia - EPA | 0.23 (ND, 0.50) | NA | 0.23 (ND, 0.50) | Q = 67.25 |
|  |  |  |  |  | I^2 = 95.54 |
|  | Asia - MESA | 1.45 (ND, 4.28) | 1 | ND (ND, 2.57) | Q = 147.08 |
|  |  |  |  |  | I^2 = 99.32 |
|  | Australia | NA | NA | NA | Q = NA |
|  |  |  |  |  | I^2 = NA |
| **Low Molecular Weight** | | | | | |
| MEP | Global | 24.84 (23.85, 25.83) | 8 | 32.87 (29.23, 36.52) | Q =6.4e+06 |
|  |  |  |  |  | I^2 = 100.00 |
|  | Latin America | 83.53 (73.08, 94.05) | 0 | 83.53 (73.08, 94.05) | Q =4.0e+05 |
|  |  |  |  |  | I^2 = 100.00 |
|  | Africa | 360.21 (133.10, 587.32) | 0 | 360.21 (133.10, 587.32) | Q =0.15 |
|  |  |  |  |  | I^2 = 0.00 |
|  | Asia - EPA | 19.80 (18.80, 20.80) | 12 | 24.11 (21.15, 27.06) | Q =5.2e+06 |
|  |  |  |  |  | I^2 = 100.00 |
|  | Asia - MESA | 353.84 (ND, 778.32) | 0 | 353.84 (ND, 778.32) | Q =941.28 |
|  |  |  |  |  | I^2 = 99.58 |
|  | Australia | 101.73 (7.16, 196.30) | 1 | 53.50 (ND, 130.78) | Q = 2232.47 |
|  |  |  |  |  | I^2 = 99.96 |
| MiBP | Global | 24.70 (23.45, 25.95) | 3 | 24.58 (23.33, 25.83) | Q =2.1e+09 |
|  |  |  |  |  | I^2 = 99.99 |
|  | Latin America | 13.54 (9.92, 17.17) | 4 | 20.90 (12.3783, 29.43) | Q =12906.83 |
|  |  |  |  |  | I^2 = 99.91 |
|  | Africa | 25.42 (9.96, 40.89) | 2 | 13.7 (0.01, 27.39) | Q =14.77 |
|  |  |  |  |  | I^2 = 86.46 |
|  | Asia - EPA | 25.78 (24.39, 27.16) | 3 | 25.56 (24.18, 26.94) | Q = 2.0e+06 |
|  |  |  |  |  | I^2 = 99.99 |
|  | Asia - MESA | 57.93 (28.90, 86.96) | 1 | 76.68 (41.67, 111.68) | Q = 649.00 |
|  |  |  |  |  | I^2 = 99.38 |
|  | Australia | 17.77 (17.52, 18.01) | 1 | 17.70 (17.48, 17.92) | Q =1.79 |
|  |  |  |  |  | I^2 = 43.98 |
| MnBP | Global | 54.08 (52.19, 55.97) | 7 | 62.16 (56.99, 67.32) | Q = 1.6e+09 |
|  |  |  |  |  | I^2 = 100 |
|  | Latin America | 36.96 (33.86, 40.06) | 2 | 29.78 (26.81, 32.73) | Q = 2.1e+07 |
|  |  |  |  |  | I^2 = 99.99 |
|  | Africa | 126.27 (58.99, 193.54) | 0 | 126.27 (58.99, 193.54) | Q = 8.39 |
|  |  |  |  |  | I^2 = 76.16 |
|  | Asia - EPA | 51.78 (50.11, 53.44) | 12 | 57.96 (55.61, 60.31) | Q = 7.3e+09 |
|  |  |  |  |  | I^2 = 100 |
|  | Asia - MESA | 159.16 (83.20, 235.11) | 0 | 159.16 (83.20, 235.11) | Q = 1458.07 |
|  |  |  |  |  | I^2 = 99.59 |
|  | Australia | 28.34 (15.31, 41.38) | 1 | 21.7 (11.03, 32.37) | Q = 799.90 |
|  |  |  |  |  | I^2 = 99.87 |
| MBzP | Global | 2.89 (2.71, 3.07) | 10 | 3.18 (2.98, 3.37) | Q = 1.1e+11 |
|  |  |  |  |  | I^2 = 100.00 |
|  | Latin America | 3.12 (2.21, 4.03) | 0 | 3.12 (2.21, 4.03) | Q = 34326.44 |
|  |  |  |  |  | I^2 = 99.96 |
|  | Africa | 2.98 (0.67, 5.29) | 1 | 2.00 (ND, 4.23) | Q = 3.20 |
|  |  |  |  |  | I^2 = 68.75 |
|  | Asia - EPA | 2.81 (2.62, 3.00) | 10 | 3.14 (2.93, 3.34) | Q = 3.0e+07 |
|  |  |  |  |  | I^2 = 100.00 |
|  | Asia - MESA | 17.20 (12.21, 22.19) | 0 | 17.20 (12.21, 22.19) | Q = 777.08 |
|  |  |  |  |  | I^2 = 99.23 |
|  | Australia | 3.95 (3.46, 4.44) | 1 | 4.20 (3.80, 4.60) | Q = 102.97 |
|  |  |  |  |  | I^2 = 99.03 |

*NA = Not Available/Applicable

**These entries were based on one article entry

***These values have values below limit of detection (LOD = 0.01)

NA = Not Available; ND = Non-Detectable; Q = Cochran's Q

Supplement Table 5: Estimated Linear Phthalate Concentrations, 2003 to 2023.

|  |  |  | **Estimate Concentrations per Year** | | | | |
| --- | --- | --- | --- | --- | --- | --- | --- |
| **Phthalate Polymers** | **Regions** | **Percentiles (%)** | **2003** | **2008** | **2013** | **2018** | **2023** |
| **High Molecular Weight** | | | | | | | |
| MEHP (ng/mL) | Global | 10 | 7.83 | 6.46 | 5.08 | 3.71 | 2.34 |
|  |  | 25 | 8.82 | 7.56 | 6.31 | 5.06 | 3.80 |
|  |  | 50 | 9.91 | 8.79 | 7.67 | 6.55 | 5.43 |
|  |  | 75 | 11.00 | 10.02 | 9.03 | 8.04 | 7.06 |
|  |  | 95 | 12.58 | 11.78 | 10.99 | 10.19 | 9.40 |
|  | Latin America | 10 | 7.29 | 5.77 | 4.24 | 2.72 | 1.20 |
|  |  | 25 | 7.81 | 6.34 | 4.87 | 3.40 | 1.93 |
|  |  | 50 | 8.39 | 6.98 | 5.57 | 4.16 | 2.75 |
|  |  | 75 | 8.97 | 7.62 | 6.27 | 4.92 | 3.57 |
|  |  | 95 | 9.80 | 8.54 | 7.27 | 6.01 | 4.74 |
|  | Africa | 10 | **<LOD** | **<LOD** | **<LOD** | **<LOD** | **<LOD** |
|  |  | 25 | 3.40 | 1.93 | 0.46 | **<LOD** | **<LOD** |
|  |  | 50 | 8.09 | 6.68 | 5.27 | 3.86 | 2.45 |
|  |  | 75 | 12.77 | 11.42 | 10.07 | 8.72 | 7.37 |
|  |  | 95 | 19.51 | 18.25 | 16.99 | 15.72 | 14.46 |
|  | Asia - EPA | 10 | 8.18 | 6.65 | 5.13 | 3.61 | 2.09 |
|  |  | 25 | 9.47 | 8.00 | 6.53 | 5.06 | 3.59 |
|  |  | 50 | 10.91 | 9.50 | 8.09 | 6.68 | 5.27 |
|  |  | 75 | 12.35 | 11.00 | 9.65 | 8.30 | 6.95 |
|  |  | 95 | 14.42 | 13.15 | 11.89 | 10.63 | 9.36 |
|  | Asia - MESA | 10 | 14.35 | 12.82 | 11.30 | 9.78 | 8.26 |
|  |  | 25 | 17.85 | 16.39 | 14.92 | 13.45 | 11.98 |
|  |  | 50 | 21.75 | 20.34 | 18.93 | 17.53 | 16.12 |
|  |  | 75 | 25.65 | 24.30 | 22.95 | 21.60 | 20.25 |
|  |  | 95 | 31.26 | 30.00 | 28.73 | 27.47 | 26.21 |
|  | Australia | 10 | **N.D.** | **N.D.** | **N.D.** | **N.D.** | **N.D.** |
|  |  | 25 | **N.D.** | **N.D.** | **N.D.** | **N.D.** | **N.D.** |
|  |  | 50 | **N.D.** | **N.D.** | **N.D.** | **N.D.** | **N.D.** |
|  |  | 75 | **N.D.** | **N.D.** | **N.D.** | **N.D.** | **N.D.** |
|  |  | 95 | **N.D.** | **N.D.** | **N.D.** | **N.D.** | **N.D.** |
| MEHHP (ng/mL) | Global | 10 | 20.659 | 17.385 | 14.111 | 10.837 | 7.563 |
|  |  | 25 | 22.762 | 19.757 | 16.752 | 13.747 | 10.742 |
|  |  | 50 | 25.098 | 22.392 | 19.686 | 16.981 | 14.275 |
|  |  | 75 | 27.434 | 25.027 | 22.621 | 20.214 | 17.807 |
|  |  | 95 | 30.795 | 28.819 | 26.842 | 24.866 | 22.889 |
|  | Latin America | 10 | 19.995 | 16.233 | 12.470 | 8.708 | 4.945 |
|  |  | 25 | 22.398 | 18.907 | 15.415 | 11.924 | 8.432 |
|  |  | 50 | 25.068 | 21.878 | 18.687 | 15.497 | 12.306 |
|  |  | 75 | 27.738 | 24.849 | 21.959 | 19.070 | 16.180 |
|  |  | 95 | 31.580 | 29.123 | 26.667 | 24.210 | 21.753 |
|  | Africa | 10 | **<LOD** | **<LOD** | **<LOD** | **<LOD** | **<LOD** |
|  |  | 25 | **<LOD** | **<LOD** | **<LOD** | **<LOD** | **<LOD** |
|  |  | 50 | 28.622 | 25.431 | 22.241 | 19.050 | 15.860 |
|  |  | 75 | 108.866 | 105.977 | 103.087 | 100.197 | 97.308 |
|  |  | 95 | 224.310 | 221.854 | 219.397 | 216.941 | 214.484 |
|  | Asia - EPA | 10 | 21.679 | 17.917 | 14.154 | 10.392 | 6.629 |
|  |  | 25 | 23.690 | 20.199 | 16.707 | 13.216 | 9.724 |
|  |  | 50 | 25.925 | 22.734 | 19.544 | 16.353 | 13.163 |
|  |  | 75 | 28.160 | 25.270 | 22.380 | 19.491 | 16.601 |
|  |  | 95 | 31.375 | 28.918 | 26.462 | 24.005 | 21.548 |
|  | Asia - MESA | 10 | 41.150 | 37.388 | 33.625 | 29.863 | 26.100 |
|  |  | 25 | 45.906 | 42.414 | 38.923 | 35.431 | 31.940 |
|  |  | 50 | 51.190 | 47.999 | 44.809 | 41.618 | 38.428 |
|  |  | 75 | 56.474 | 53.585 | 50.695 | 47.805 | 44.916 |
|  |  | 95 | 64.076 | 61.620 | 59.163 | 56.707 | 54.250 |
|  | Australia | 10 | 17.438 | 13.676 | 9.913 | 6.151 | 2.388 |
|  |  | 25 | 20.994 | 17.502 | 14.011 | 10.519 | 7.027 |
|  |  | 50 | 24.944 | 21.754 | 18.563 | 15.372 | 12.182 |
|  |  | 75 | 28.895 | 26.005 | 23.115 | 20.226 | 17.336 |
|  |  | 95 | 34.578 | 32.121 | 29.665 | 27.208 | 24.752 |
| MEOHP (ng/mL) | Global | 10 | 15.80 | 12.07 | 8.34 | 4.62 | 0.89 |
|  |  | 25 | 17.40 | 13.87 | 10.34 | 6.81 | 3.28 |
|  |  | 50 | 19.18 | 15.87 | 12.56 | 9.25 | 5.94 |
|  |  | 75 | 20.95 | 17.86 | 14.78 | 11.69 | 8.61 |
|  |  | 95 | 23.51 | 20.74 | 17.97 | 15.20 | 12.43 |
|  | Latin America | 10 | 14.61 | 10.62 | 6.64 | 2.65 | **<LOD** |
|  |  | 25 | 16.28 | 12.47 | 8.65 | 4.84 | 1.02 |
|  |  | 50 | 18.14 | 14.52 | 10.89 | 7.27 | 3.65 |
|  |  | 75 | 20.00 | 16.57 | 13.13 | 9.70 | 6.27 |
|  |  | 95 | 22.67 | 19.51 | 16.36 | 13.20 | 10.05 |
|  | Africa | 10 | **<LOD** | **<LOD** | **<LOD** | **<LOD** | **<LOD** |
|  |  | 25 | 6.75 | 2.93 | **<LOD** | **<LOD** | **<LOD** |
|  |  | 50 | 23.45 | 19.82 | 16.20 | 12.57 | 8.95 |
|  |  | 75 | 40.14 | 36.71 | 33.28 | 29.85 | 26.42 |
|  |  | 95 | 64.17 | 61.01 | 57.85 | 54.70 | 51.54 |
|  | Asia - EPA | 10 | 16.68 | 12.69 | 8.70 | 4.71 | 0.73 |
|  |  | 25 | 18.32 | 14.50 | 10.68 | 6.87 | 3.05 |
|  |  | 50 | 20.13 | 16.51 | 12.89 | 9.26 | 5.64 |
|  |  | 75 | 21.95 | 18.52 | 15.09 | 11.65 | 8.22 |
|  |  | 95 | 24.56 | 21.41 | 18.25 | 15.10 | 11.94 |
|  | Asia - MESA | 10 | 23.97 | 19.98 | 15.99 | 12.00 | 8.01 |
|  |  | 25 | 28.21 | 24.39 | 20.58 | 16.76 | 12.94 |
|  |  | 50 | 32.92 | 29.30 | 25.67 | 22.05 | 18.43 |
|  |  | 75 | 37.63 | 34.20 | 30.77 | 27.34 | 23.91 |
|  |  | 95 | 44.41 | 41.26 | 38.10 | 34.95 | 31.79 |
|  | Australia | 10 | 15.18 | 11.19 | 7.20 | 3.21 | **<LOD** |
|  |  | 25 | 17.57 | 13.76 | 9.94 | 6.13 | 2.31 |
|  |  | 50 | 20.23 | 16.61 | 12.98 | 9.36 | 5.73 |
|  |  | 75 | 22.89 | 19.46 | 16.02 | 12.59 | 9.16 |
|  |  | 95 | 26.71 | 23.55 | 20.40 | 17.24 | 14.09 |
| MECPP (ng/mL) | Global | 10 | 34.93 | 28.59 | 22.26 | 15.92 | 9.59 |
|  |  | 25 | 37.08 | 31.48 | 25.88 | 20.28 | 14.69 |
|  |  | 50 | 39.47 | 34.69 | 29.91 | 25.13 | 20.35 |
|  |  | 75 | 41.86 | 37.90 | 33.94 | 29.98 | 26.02 |
|  |  | 95 | 45.30 | 42.51 | 39.73 | 36.95 | 34.17 |
|  | Latin America | 10 | 37.75 | 31.29 | 24.82 | 18.35 | 11.88 |
|  |  | 25 | 42.02 | 36.97 | 31.91 | 26.86 | 21.80 |
|  |  | 50 | 46.77 | 43.28 | 39.80 | 36.32 | 32.83 |
|  |  | 75 | 51.51 | 49.60 | 47.69 | 45.77 | 43.86 |
|  |  | 95 | 58.33 | 58.68 | 59.03 | 59.38 | 59.73 |
|  | Africa | 10 | **<LOD** | **<LOD** | **<LOD** | **<LOD** | **<LOD** |
|  |  | 25 | 24.97 | 19.91 | 14.86 | 9.80 | 4.75 |
|  |  | 50 | 76.19 | 72.71 | 69.23 | 65.74 | 62.26 |
|  |  | 75 | 127.42 | 125.50 | 123.59 | 121.68 | 119.77 |
|  |  | 95 | 201.11 | 201.46 | 201.81 | 202.16 | 202.50 |
|  | Asia - EPA | 10 | **N.D.** | 29.64 | 23.17 | 16.70 | 10.23 |
|  |  | 25 | **N.D.** | 30.24 | 25.18 | 20.13 | 15.07 |
|  |  | 50 | **N.D.** | 30.90 | 27.42 | 23.93 | 20.45 |
|  |  | 75 | **N.D.** | 31.56 | 29.65 | 27.74 | 25.83 |
|  |  | 95 | **N.D.** | 32.52 | 32.87 | 33.21 | 33.56 |
|  | Asia - MESA | 10 | 40.33 | 33.86 | 27.39 | 20.93 | 14.46 |
|  |  | 25 | 42.72 | 37.66 | 32.61 | 27.55 | 22.50 |
|  |  | 50 | 45.37 | 41.88 | 38.40 | 34.92 | 31.43 |
|  |  | 75 | 48.01 | 46.10 | 44.19 | 42.28 | 40.37 |
|  |  | 95 | 51.83 | 52.17 | 52.52 | 52.87 | 53.22 |
|  | Australia | 10 | 26.14 | 19.67 | 13.20 | 6.73 | 0.26 |
|  |  | 25 | 28.35 | 23.29 | 18.24 | 13.18 | 8.13 |
|  |  | 50 | 30.80 | 27.32 | 23.84 | 20.35 | 16.87 |
|  |  | 75 | 33.26 | 31.34 | 29.43 | 27.52 | 25.61 |
|  |  | 95 | 36.78 | 37.13 | 37.48 | 37.83 | 38.18 |
| MCMHP (ng/mL) | Global | 10 | 12.71 | 11.79 | 10.88 | 9.96 | 9.04 |
|  |  | 25 | 17.26 | 15.83 | 14.40 | 12.98 | 11.55 |
|  |  | 50 | 22.31 | 20.32 | 18.32 | 16.33 | 14.33 |
|  |  | 75 | 27.36 | 24.80 | 22.24 | 19.68 | 17.12 |
|  |  | 95 | 34.63 | 31.26 | 27.88 | 24.51 | 21.13 |
|  | Latin America | 10 | 18.00 | 16.94 | 15.87 | 14.81 | 13.75 |
|  |  | 25 | 28.52 | 27.13 | 25.73 | 24.34 | 22.94 |
|  |  | 50 | 40.22 | 38.45 | 36.68 | 34.91 | 33.15 |
|  |  | 75 | 51.92 | 49.78 | 47.63 | 45.49 | 43.35 |
|  |  | 95 | 68.74 | 66.07 | 63.39 | 60.71 | 58.04 |
|  | Africa | 10 | **N.D.** | **N.D.** | **N.D.** | **N.D.** | **N.D.** |
|  |  | 25 | **N.D.** | **N.D.** | **N.D.** | **N.D.** | **N.D.** |
|  |  | 50 | **N.D.** | **N.D.** | **N.D.** | **N.D.** | **N.D.** |
|  |  | 75 | **N.D.** | **N.D.** | **N.D.** | **N.D.** | **N.D.** |
|  |  | 95 | **N.D.** | **N.D.** | **N.D.** | **N.D.** | **N.D.** |
|  | Asia - EPA | 10 | 13.10 | 12.04 | 10.98 | 9.91 | **<LOD** |
|  |  | 25 | 17.25 | 15.86 | 14.46 | 13.06 | 11.67 |
|  |  | 50 | 21.87 | 20.10 | 18.33 | 16.57 | 14.80 |
|  |  | 75 | 26.49 | 24.35 | 22.21 | 20.07 | 17.93 |
|  |  | 95 | 33.13 | 30.46 | 27.78 | 25.10 | 22.43 |
|  | Asia - MESA | 10 | 16.48 | 15.42 | 14.36 | 13.29 | 12.23 |
|  |  | 25 | 21.29 | 19.89 | 18.50 | 17.10 | 15.70 |
|  |  | 50 | 26.63 | 24.86 | 23.10 | 21.33 | 19.56 |
|  |  | 75 | 31.98 | 29.84 | 27.70 | 25.56 | 23.41 |
|  |  | 95 | 39.67 | 36.99 | 34.31 | 31.64 | 28.96 |
|  | Australia | 10 | **N.D.** | **N.D.** | **N.D.** | **N.D.** | **N.D.** |
|  |  | 25 | **N.D.** | **N.D.** | **N.D.** | **N.D.** | **N.D.** |
|  |  | 50 | **N.D.** | **N.D.** | **N.D.** | **N.D.** | **N.D.** |
|  |  | 75 | **N.D.** | **N.D.** | **N.D.** | **N.D..** | **N.D.** |
|  |  | 95 | **N.D.** | **N.D.** | **N.D.** | **N.D.** | **N.D.** |
| MCOP (ng/mL) | Global | 10 | 5.32 | 3.71 | 2.10 | 0.48 | **N.D.** |
|  |  | 25 | 6.15 | 4.29 | 2.42 | 0.56 | **N.D.** |
|  |  | 50 | 7.06 | 4.92 | 2.79 | 0.65 | **N.D.** |
|  |  | 75 | 7.98 | 5.56 | 3.15 | 0.74 | **N.D.** |
|  |  | 95 | 9.29 | 6.48 | 3.67 | 0.86 | **N.D.** |
|  | Latin America | 10 | 4.63 | 3.47 | 2.32 | 1.17 | **<LOD** |
|  |  | 25 | 5.03 | 4.11 | 3.19 | 2.26 | 1.34 |
|  |  | 50 | 5.48 | 4.81 | 4.14 | 3.47 | 2.81 |
|  |  | 75 | 5.93 | 5.51 | 5.10 | 4.69 | 4.27 |
|  |  | 95 | 6.58 | 6.53 | 6.48 | 6.43 | 6.38 |
|  | Africa | 10 | **<LOD** | **<LOD** | **<LOD** | **<LOD** | **<LOD** |
|  |  | 25 | **<LOD** | **<LOD** | **<LOD** | **<LOD** | **<LOD** |
|  |  | 50 | 1.83 | 1.16 | 0.50 | **<LOD** | **<LOD** |
|  |  | 75 | 3.64 | 3.22 | 2.81 | 2.39 | 1.98 |
|  |  | 95 | 6.23 | 6.18 | 6.13 | 6.08 | 6.04 |
|  | Asia - EPA | 10 | **N.D.** | **N.D.** | **N.D.** | **<LOD** | **N.D.** |
|  |  | 25 | **N.D.** | **N.D.** | **N.D.** | 0.07 | **N.D.** |
|  |  | 50 | **N.D.** | **N.D.** | **N.D.** | 0.23 | **N.D.** |
|  |  | 75 | **N.D.** | **N.D.** | **N.D.** | 0.38 | **N.D.** |
|  |  | 95 | **N.D.** | **N.D.** | **N.D.** | 0.60 | **N.D.** |
|  | Asia - MESA | 10 | 2.16 | 1.01 | **<LOD** | **<LOD** | **<LOD** |
|  |  | 25 | 3.21 | 2.29 | 1.37 | 0.45 | **<LOD** |
|  |  | 50 | 4.38 | 3.71 | 3.04 | 2.37 | 1.70 |
|  |  | 75 | 5.54 | 5.13 | 4.72 | 4.30 | 3.89 |
|  |  | 95 | 7.22 | 7.17 | 7.12 | 7.07 | 7.02 |
|  | Australia | 10 | **N.D.** | **N.D.** | **N.D.** | **N.D.** | **N.D.** |
|  |  | 25 | **N.D.** | **N.D.** | **N.D.** | **N.D.** | **N.D.** |
|  |  | 50 | **N.D.** | **N.D.** | **N.D.** | **N.D.** | **N.D.** |
|  |  | 75 | **N.D.** | **N.D.** | **N.D.** | **N.D.** | **N.D.** |
|  |  | 95 | **N.D.** | **N.D.** | **N.D.** | **N.D.** | **N.D.** |
| MiNP (ng/mL) | Global | 10 | 1.432 | 1.095 | 0.759 | 0.422 | **<LOD** |
|  |  | 25 | 1.569 | 1.234 | 0.898 | 0.562 | 0.227 |
|  |  | 50 | 1.722 | 1.388 | 1.053 | 0.718 | 0.384 |
|  |  | 75 | 1.875 | 1.541 | 1.208 | 0.874 | 0.540 |
|  |  | 95 | 2.095 | 1.763 | 1.430 | 1.098 | 0.766 |
|  | Latin America | 10 | **N.D.** | 0.228 | **<LOD** | **<LOD** | **<LOD** |
|  |  | 25 | **N.D.** | 0.234 | **<LOD** | **<LOD** | **<LOD** |
|  |  | 50 | **N.D.** | 0.240 | **<LOD** | **<LOD** | **<LOD** |
|  |  | 75 | **N.D.** | 0.247 | **<LOD** | **<LOD** | **<LOD** |
|  |  | 95 | **N.D.** | 0.257 | **<LOD** | **<LOD** | **<LOD** |
|  | Africa | 10 | **N.D.** | **N.D.** | **N.D.** | **N.D.** | **N.D.** |
|  |  | 25 | **N.D.** | **N.D.** | **N.D.** | **N.D.** | **N.D.** |
|  |  | 50 | **N.D.** | **N.D.** | **N.D.** | **N.D.** | **N.D.** |
|  |  | 75 | **N.D.** | **N.D.** | **N.D.** | **N.D.** | **N.D.** |
|  |  | 95 | **N.D.** | **N.D.** | **N.D.** | **N.D.** | **N.D.** |
|  | Asia - EPA | 10 | 1.286 | 0.995 | 0.704 | 0.414 | 0.123 |
|  |  | 25 | 1.400 | 1.118 | 0.837 | 0.555 | 0.273 |
|  |  | 50 | 1.528 | 1.256 | 0.984 | 0.711 | 0.439 |
|  |  | 75 | 1.656 | 1.393 | 1.131 | 0.868 | 0.606 |
|  |  | 95 | 1.839 | 1.590 | 1.342 | 1.093 | 0.845 |
|  | Asia - MESA | 10 | **<LOD** | **<LOD** | **<LOD** | **<LOD** | **<LOD** |
|  |  | 25 | **<LOD** | **<LOD** | **<LOD** | **<LOD** | **<LOD** |
|  |  | 50 | 5.559 | 5.287 | 5.014 | 4.742 | 4.470 |
|  |  | 75 | 21.880 | 21.618 | 21.355 | 21.093 | 20.830 |
|  |  | 95 | 45.361 | 45.112 | 44.864 | 44.615 | 44.367 |
|  | Australia | 10 | **N.D.** | **N.D.** | **N.D.** | **N.D.** | **N.D.** |
|  |  | 25 | **N.D.** | **N.D.** | **N.D.** | **N.D.** | **N.D.** |
|  |  | 50 | **N.D.** | **N.D.** | **N.D.** | **N.D.** | **N.D.** |
|  |  | 75 | **N.D.** | **N.D.** | **N.D.** | **N.D.** | **N.D.** |
|  |  | 95 | **N.D.** | **N.D.** | **N.D.** | **N.D.** | **N.D.** |
| MOiNP (ng/mL) | Global | 10 | 3.838 | 2.425 | 1.011 | **<LOD** | **<LOD** |
|  |  | 25 | 4.137 | 3.340 | 2.542 | 1.745 | 0.948 |
|  |  | 50 | 4.469 | 4.357 | 4.244 | 4.132 | 4.020 |
|  |  | 75 | 4.801 | 5.374 | 5.946 | 6.519 | 7.091 |
|  |  | 95 | 5.279 | 6.837 | 8.395 | 9.953 | 11.510 |
|  | Latin America | 10 | **N.D.** | **N.D.** | **N.D.** | **N.D.** | **N.D.** |
|  |  | 25 | **N.D.** | **N.D.** | **N.D.** | **N.D.** | **N.D.** |
|  |  | 50 | **N.D.** | **N.D.** | **N.D.** | **N.D.** | **N.D.** |
|  |  | 75 | **N.D.** | **N.D.** | **N.D.** | **N.D.** | **N.D.** |
|  |  | 95 | **N.D.** | **N.D.** | **N.D.** | **N.D.** | **N.D.** |
|  | Africa | 10 | **N.D.** | **N.D.** | **N.D.** | **N.D.** | **N.D.** |
|  |  | 25 | **N.D.** | **N.D.** | **N.D.** | **N.D.** | **N.D.** |
|  |  | 50 | **N.D.** | **N.D.** | **N.D.** | **N.D.** | **N.D.** |
|  |  | 75 | **N.D.** | **N.D.** | **N.D.** | **N.D.** | **N.D.** |
|  |  | 95 | **N.D.** | **N.D.** | **N.D.** | **N.D.** | **N.D.** |
|  | Asia - EPA | 10 | 3.512 | 2.483 | 1.454 | 0.425 | **<LOD** |
|  |  | 25 | 4.361 | 3.438 | 2.514 | 1.591 | 0.667 |
|  |  | 50 | 5.306 | 4.499 | 3.692 | 2.885 | 2.079 |
|  |  | 75 | 6.250 | 5.560 | 4.870 | 4.180 | 3.490 |
|  |  | 95 | 7.609 | 7.087 | 6.565 | 6.043 | 5.521 |
|  | Asia - MESA | 10 | **<LOD** | **<LOD** | **<LOD** | **<LOD** | **<LOD** |
|  |  | 25 | 5.325 | 4.401 | 3.478 | 2.554 | 1.630 |
|  |  | 50 | 10.426 | 9.619 | 8.812 | 8.005 | 7.199 |
|  |  | 75 | 15.527 | 14.837 | 14.147 | 13.457 | 12.767 |
|  |  | 95 | 22.865 | 22.344 | 21.822 | 21.300 | 20.778 |
|  | Australia | 10 | **N.D.** | **N.D.** | **N.D.** | **N.D.** | **N.D.** |
|  |  | 25 | **N.D.** | **N.D.** | **N.D.** | **N.D.** | **N.D.** |
|  |  | 50 | **N.D.** | **N.D.** | **N.D.** | **N.D.** | **N.D.** |
|  |  | 75 | **N.D.** | **N.D.** | **N.D.** | **N.D.** | **N.D.** |
|  |  | 95 | **N.D.** | **N.D.** | **N.D.** | **N.D.** | **N.D.** |
| OH-MiNP (ng/mL) | Global | 10 | 5.430 | 4.059 | 2.689 | 1.318 | **<LOD** |
|  |  | 25 | 6.100 | 5.168 | 4.237 | 3.306 | 2.374 |
|  |  | 50 | 6.844 | 6.400 | 5.957 | 5.513 | 5.070 |
|  |  | 75 | 7.588 | 7.632 | 7.677 | 7.721 | 7.765 |
|  |  | 95 | 8.658 | 9.404 | 10.151 | 10.897 | 11.644 |
|  | Latin America | 10 | **N.D.** | **N.D.** | **N.D.** | **N.D.** | **N.D.** |
|  |  | 25 | **N.D.** | **N.D.** | **N.D.** | **N.D.** | **N.D.** |
|  |  | 50 | **N.D.** | **N.D.** | **N.D.** | **N.D.** | **N.D.** |
|  |  | 75 | **N.D.** | **N.D.** | **N.D.** | **N.D.** | **N.D.** |
|  |  | 95 | **N.D.** | **N.D.** | **N.D.** | **N.D.** | **N.D.** |
|  | Africa | 10 | **N.D.** | **N.D.** | **N.D.** | **N.D.** | **N.D.** |
|  |  | 25 | **N.D.** | **N.D.** | **N.D.** | **N.D.** | **N.D.** |
|  |  | 50 | **N.D.** | **N.D.** | **N.D.** | **N.D.** | **N.D.** |
|  |  | 75 | **N.D.** | **N.D.** | **N.D.** | **N.D.** | **N.D.** |
|  |  | 95 | **N.D.** | **N.D.** | **N.D.** | **N.D.** | **N.D.** |
|  | Asia - EPA | 10 | 5.327 | 4.132 | 2.936 | 1.741 | 0.545 |
|  |  | 25 | 6.265 | 5.253 | 4.241 | 3.230 | 2.218 |
|  |  | 50 | 7.306 | 6.499 | 5.692 | 4.885 | 4.078 |
|  |  | 75 | 8.348 | 7.745 | 7.142 | 6.539 | 5.937 |
|  |  | 95 | 9.846 | 9.537 | 9.229 | 8.920 | 8.611 |
|  | Asia - MESA | 10 | **<LOD** | **<LOD** | **<LOD** | **<LOD** | **<LOD** |
|  |  | 25 | 6.761 | 5.750 | 4.738 | 3.727 | 2.715 |
|  |  | 50 | 13.382 | 12.575 | 11.768 | 10.961 | 10.154 |
|  |  | 75 | 20.003 | 19.401 | 18.798 | 18.195 | 17.592 |
|  |  | 95 | 29.529 | 29.220 | 28.911 | 28.603 | 28.294 |
|  | Australia | 10 | **N.D.** | **N.D.** | **N.D.** | **N.D.** | **N.D.** |
|  |  | 25 | **N.D.** | **N.D.** | **N.D.** | **N.D.** | **N.D.** |
|  |  | 50 | **N.D.** | **N.D.** | **N.D.** | **N.D.** | **N.D.** |
|  |  | 75 | **N.D.** | **N.D.** | **N.D.** | **N.D.** | **N.D.** |
|  |  | 95 | **N.D.** | **N.D.** | **N.D.** | **N.D.** | **N.D.** |
| MCiNP (ng/mL) | Global | 10 | **N.D.** | **N.D.** | **<LOD** | **<LOD** | **<LOD** |
|  |  | 25 | **N.D.** | **N.D.** | 0.138 | **<LOD** | **<LOD** |
|  |  | 50 | **N.D.** | **N.D.** | 0.200 | 0.199 | 0.199 |
|  |  | 75 | **N.D.** | **N.D.** | 0.262 | 0.351 | 0.440 |
|  |  | 95 | **N.D.** | **N.D.** | 0.351 | 0.569 | 0.786 |
|  | Latin America | 10 | **N.D.** | **N.D.** | 0.261 | **<LOD** | **<LOD** |
|  |  | 25 | **N.D.** | **N.D.** | 0.297 | **<LOD** | **<LOD** |
|  |  | 50 | **N.D.** | **N.D.** | 0.337 | **<LOD** | **<LOD** |
|  |  | 75 | **N.D.** | **N.D.** | 0.377 | **<LOD** | **<LOD** |
|  |  | 95 | **N.D.** | **N.D.** | 0.434 | 0.239 | **<LOD** |
|  | Africa | 10 | **<LOD** | **<LOD** | **<LOD** | **<LOD** | **<LOD** |
|  |  | 25 | 0.535 | **<LOD** | **<LOD** | **<LOD** | **<LOD** |
|  |  | 50 | 1.737 | 1.053 | 0.370 | **<LOD** | **<LOD** |
|  |  | 75 | 2.938 | 2.455 | 1.972 | 1.489 | 1.006 |
|  |  | 95 | 4.667 | 4.471 | 4.276 | 4.081 | 3.885 |
|  | Asia - EPA | 10 | **N.D.** | **N.D.** | **N.D.** | **<LOD** | **<LOD** |
|  |  | 25 | **N.D.** | **N.D.** | **N.D.** | **<LOD** | **<LOD** |
|  |  | 50 | **N.D.** | **N.D.** | **N.D.** | **<LOD** | **<LOD** |
|  |  | 75 | **N.D.** | **N.D.** | **N.D.** | 0.114 | **<LOD** |
|  |  | 95 | **N.D.** | **N.D.** | **N.D.** | 0.176 | **<LOD** |
|  | Asia - MESA | 10 | 10.601 | 9.538 | 8.474 | 7.411 | 6.348 |
|  |  | 25 | 12.967 | 12.084 | 11.200 | 10.317 | 9.434 |
|  |  | 50 | 15.596 | 14.912 | 14.229 | 13.546 | 12.863 |
|  |  | 75 | 18.224 | 17.741 | 17.258 | 16.775 | 16.292 |
|  |  | 95 | 22.006 | 21.811 | 21.615 | 21.420 | 21.225 |
|  | Australia | 10 | **N.D.** | **N.D.** | **N.D.** | **N.D.** | **N.D.** |
|  |  | 25 | **N.D.** | **N.D.** | **N.D.** | **N.D.** | **N.D.** |
|  |  | 50 | **N.D.** | **N.D.** | **N.D.** | **N.D.** | **N.D.** |
|  |  | 75 | **N.D.** | **N.D.** | **N.D.** | **N.D.** | **N.D.** |
|  |  | 95 | **N.D.** | **N.D.** | **N.D.** | **N.D.** | **N.D.** |
| MOP (ng/mL) | Global | 10 | 1.234 | 1.072 | 0.910 | **<LOD** | **<LOD** |
|  |  | 25 | 1.303 | 1.159 | 1.015 | 0.872 | 0.728 |
|  |  | 50 | 1.380 | 1.257 | 1.133 | 1.009 | 0.885 |
|  |  | 75 | 1.458 | 1.354 | 1.250 | 1.146 | 1.042 |
|  |  | 95 | 1.569 | 1.494 | 1.419 | 1.344 | 1.269 |
|  | Latin America | 10 | **N.D.** | **N.D.** | **N.D.** | **N.D.** | **N.D.** |
|  |  | 25 | **N.D.** | **N.D.** | **N.D.** | **N.D.** | **N.D.** |
|  |  | 50 | **N.D.** | **N.D.** | **N.D.** | **N.D.** | **N.D.** |
|  |  | 75 | **N.D.** | **N.D.** | **N.D.** | **N.D.** | **N.D.** |
|  |  | 95 | **N.D.** | **N.D.** | **N.D.** | **N.D.** | **N.D.** |
|  | Africa | 10 | **N.D.** | **N.D.** | **N.D.** | **N.D.** | **N.D.** |
|  |  | 25 | **N.D.** | **N.D.** | **N.D.** | **N.D.** | **N.D.** |
|  |  | 50 | **N.D.** | **N.D.** | **N.D.** | **N.D.** | **N.D.** |
|  |  | 75 | **N.D.** | **N.D.** | **N.D.** | **N.D.** | **N.D.** |
|  |  | 95 | **N.D.** | **N.D.** | **N.D.** | **N.D.** | **N.D.** |
|  | Asia - EPA | 10 | **N.D.** | **N.D.** | 0.106 | **<LOD** | **<LOD** |
|  |  | 25 | **N.D.** | **N.D.** | 0.126 | **<LOD** | **<LOD** |
|  |  | 50 | **N.D.** | **N.D.** | 0.148 | 0.115 | **<LOD** |
|  |  | 75 | **N.D.** | **N.D.** | 0.170 | 0.179 | 0.187 |
|  |  | 95 | **N.D.** | **N.D.** | 0.202 | 0.270 | 0.337 |
|  | Asia - MESA | 10 | **<LOD** | **<LOD** | **<LOD** | **<LOD** | **<LOD** |
|  |  | 25 | 2.020 | 1.946 | 1.871 | 1.797 | 1.722 |
|  |  | 50 | 5.023 | 4.990 | 4.957 | 4.924 | 4.890 |
|  |  | 75 | 8.026 | 8.034 | 8.042 | 8.050 | 8.059 |
|  |  | 95 | 12.346 | 12.414 | 12.481 | 12.549 | 12.616 |
|  | Australia | 10 | **N.D.** | **N.D.** | **N.D.** | **N.D.** | **N.D.** |
|  |  | 25 | **N.D.** | **N.D.** | **N.D.** | **N.D.** | **N.D.** |
|  |  | 50 | **N.D.** | **N.D.** | **N.D.** | **N.D.** | **N.D.** |
|  |  | 75 | **N.D.** | **N.D.** | **N.D.** | **N.D.** | **N.D.** |
|  |  | 95 | **N.D.** | **N.D.** | **N.D.** | **N.D.** | **N.D.** |
| MCPP (ng/mL) | Global | 10 | 1.552 | 1.327 | 1.102 | 0.877 | 0.652 |
|  |  | 25 | 1.644 | 1.457 | 1.271 | 1.084 | 0.898 |
|  |  | 50 | 1.747 | 1.603 | 1.459 | 1.315 | 1.171 |
|  |  | 75 | 1.849 | 1.748 | 1.647 | 1.546 | 1.444 |
|  |  | 95 | 1.997 | 1.957 | 1.917 | 1.878 | 1.838 |
|  | Latin America | 10 | **N.D.** | 1.462 | 0.834 | 0.206 | **<LOD** |
|  |  | 25 | **N.D.** | 1.485 | 0.934 | 0.382 | **<LOD** |
|  |  | 50 | **N.D.** | 1.512 | 1.044 | 0.577 | 0.110 |
|  |  | 75 | **N.D.** | 1.538 | 1.155 | 0.772 | 0.390 |
|  |  | 95 | **N.D.** | 1.575 | 1.314 | 1.053 | 0.792 |
|  | Africa | 10 | **N.D.** | **N.D.** | **N.D.** | **N.D.** | **N.D.** |
|  |  | 25 | **N.D.** | **N.D.** | **N.D.** | **N.D.** | **N.D.** |
|  |  | 50 | **N.D.** | **N.D.** | **N.D.** | **N.D.** | **N.D.** |
|  |  | 75 | **N.D.** | **N.D.** | **N.D.** | **N.D.** | **N.D.** |
|  |  | 95 | **N.D.** | **N.D.** | **N.D.** | **N.D.** | **N.D.** |
|  | Asia - EPA | 10 | **N.D.** | **N.D.** | **N.D.** | 1.489 | 0.861 |
|  |  | 25 | **N.D.** | **N.D.** | **N.D.** | 1.512 | 0.960 |
|  |  | 50 | **N.D.** | **N.D.** | **N.D.** | 1.537 | 1.070 |
|  |  | 75 | **N.D.** | **N.D.** | **N.D.** | 1.562 | 1.179 |
|  |  | 95 | **N.D.** | **N.D.** | **N.D.** | 1.598 | 1.337 |
|  | Asia - MESA | 10 | **N.D.** | **N.D.** | **N.D.** | **N.D.** | 0.127 |
|  |  | 25 | **N.D.** | **N.D.** | **N.D.** | **N.D.** | 0.188 |
|  |  | 50 | **N.D.** | **N.D.** | **N.D.** | **N.D.** | 0.255 |
|  |  | 75 | **N.D.** | **N.D.** | **N.D.** | **N.D.** | 0.323 |
|  |  | 95 | **N.D.** | **N.D.** | **N.D.** | **N.D.** | 0.420 |
|  | Australia | 10 | **<LOD** | **<LOD** | **<LOD** | **<LOD** | **<LOD** |
|  |  | 25 | **<LOD** | **<LOD** | **<LOD** | **<LOD** | **<LOD** |
|  |  | 50 | 1.451 | 0.984 | 0.517 | **<LOD** | **<LOD** |
|  |  | 75 | 2.867 | 2.485 | 2.102 | 1.719 | 1.336 |
|  |  | 95 | 4.904 | 4.643 | 4.382 | 4.121 | 3.860 |
| MiDP (ng/mL) a. | Global | 10 | **N.D.** | **N.D.** | **N.D.** | **<LOD** | **<LOD** |
|  |  | 25 | **N.D.** | **N.D.** | **N.D.** | 0.448 | **<LOD** |
|  |  | 50 | **N.D.** | **N.D.** | **N.D.** | 1.933 | 3.066 |
|  |  | 75 | **N.D.** | **N.D.** | **N.D.** | 3.418 | 6.234 |
|  |  | 95 | **N.D.** | **N.D.** | **N.D.** | 5.554 | 10.791 |
|  | Latin America | 10 | **N.D.** | **N.D.** | **N.D.** | **N.D.** | **N.D.** |
|  |  | 25 | **N.D.** | **N.D.** | **N.D.** | **N.D.** | **N.D.** |
|  |  | 50 | **N.D.** | **N.D.** | **N.D.** | **N.D.** | **N.D.** |
|  |  | 75 | **N.D.** | **N.D.** | **N.D.** | **N.D.** | **N.D.** |
|  |  | 95 | **N.D.** | **N.D.** | **N.D.** | **N.D.** | **N.D.** |
|  | Africa | 10 | **N.D.** | **N.D.** | **N.D.** | **N.D.** | **N.D.** |
|  |  | 25 | **N.D.** | **N.D.** | **N.D.** | **N.D.** | **N.D.** |
|  |  | 50 | **N.D.** | **N.D.** | **N.D.** | **N.D.** | **N.D.** |
|  |  | 75 | **N.D.** | **N.D.** | **N.D.** | **N.D.** | **N.D.** |
|  |  | 95 | **N.D.** | **N.D.** | **N.D.** | **N.D.** | **N.D.** |
|  | Asia - EPA | 10 | **N.D.** | **N.D.** | **N.D.** | **<LOD** | **<LOD** |
|  |  | 25 | **N.D.** | **N.D.** | **N.D.** | **<LOD** | **<LOD** |
|  |  | 50 | **N.D.** | **N.D.** | **N.D.** | 1.517 | 2.277 |
|  |  | 75 | **N.D.** | **N.D.** | **N.D.** | 3.465 | 6.371 |
|  |  | 95 | **N.D.** | **N.D.** | **N.D.** | 6.266 | 12.260 |
|  | Asia - MESA | 10 | **N.D.** | **N.D.** | **N.D.** | **<LOD** | **<LOD** |
|  |  | 25 | **N.D.** | **N.D.** | **N.D.** | **<LOD** | **<LOD** |
|  |  | 50 | **N.D.** | **N.D.** | **N.D.** | 1.460 | 2.220 |
|  |  | 75 | **N.D.** | **N.D.** | **N.D.** | 2.850 | 5.756 |
|  |  | 95 | **N.D.** | **N.D.** | **N.D.** | 4.850 | 10.844 |
|  | Australia | 10 | **N.D.** | **N.D.** | **N.D.** | **N.D.** | **N.D.** |
|  |  | 25 | **N.D.** | **N.D.** | **N.D.** | **N.D.** | **N.D.** |
|  |  | 50 | **N.D.** | **N.D.** | **N.D.** | **N.D.** | **N.D.** |
|  |  | 75 | **N.D.** | **N.D.** | **N.D.** | **N.D.** | **N.D.** |
|  |  | 95 | **N.D.** | **N.D.** | **N.D.** | **N.D.** | **N.D.** |
| **Low Molecular Weight** | | | | | | | |
| MEP (ng/mL) | Global | 10 | 45.680 | 35.547 | 25.413 | 15.280 | 5.146 |
|  |  | 25 | 49.847 | 40.179 | 30.510 | 20.841 | 11.173 |
|  |  | 50 | 54.477 | 45.325 | 36.173 | 27.021 | 17.869 |
|  |  | 75 | 59.107 | 50.471 | 41.836 | 33.200 | 24.565 |
|  |  | 95 | 65.767 | 57.875 | 49.983 | 42.091 | 34.198 |
|  | Latin America | 10 | 74.380 | 67.343 | 60.306 | 53.269 | 46.232 |
|  |  | 25 | 77.799 | 71.177 | 64.555 | 57.932 | 51.310 |
|  |  | 50 | 81.598 | 75.437 | 69.275 | 63.114 | 56.952 |
|  |  | 75 | 85.397 | 79.696 | 73.995 | 68.295 | 62.594 |
|  |  | 95 | 90.862 | 85.824 | 80.786 | 75.749 | 70.711 |
|  | Africa | 10 | **<LOD** | **<LOD** | **<LOD** | **<LOD** | **<LOD** |
|  |  | 25 | **<LOD** | **<LOD** | **<LOD** | **<LOD** | **<LOD** |
|  |  | 50 | 376.278 | 370.117 | 363.955 | 357.794 | 351.632 |
|  |  | 75 | 1008.668 | 1002.967 | 997.266 | 991.566 | 985.865 |
|  |  | 95 | 1918.464 | 1913.426 | 1908.388 | 1903.351 | 1898.313 |
|  | Asia - EPA | 10 | 25.806 | 18.769 | 11.732 | 4.695 | -2.342 |
|  |  | 25 | 30.556 | 23.934 | 17.312 | 10.689 | 4.067 |
|  |  | 50 | 35.834 | 29.672 | 23.511 | 17.349 | 11.188 |
|  |  | 75 | 41.112 | 35.411 | 29.710 | 24.010 | 18.309 |
|  |  | 95 | 48.705 | 43.667 | 38.629 | 33.592 | 28.554 |
|  | Asia - MESA | 10 | 70.092 | 63.055 | 56.018 | 48.981 | 41.944 |
|  |  | 25 | 74.601 | 67.979 | 61.357 | 54.734 | 48.112 |
|  |  | 50 | 79.611 | 73.450 | 67.288 | 61.127 | 54.965 |
|  |  | 75 | 84.621 | 78.920 | 73.220 | 67.519 | 61.818 |
|  |  | 95 | 91.829 | 86.791 | 81.753 | 76.715 | 71.678 |
|  | Australia | 10 | 94.858 | 87.821 | 80.784 | 73.747 | 66.710 |
|  |  | 25 | 102.890 | 96.268 | 89.646 | 83.023 | 76.401 |
|  |  | 50 | 111.815 | 105.653 | 99.492 | 93.330 | 87.169 |
|  |  | 75 | 120.740 | 115.039 | 109.338 | 103.638 | 97.937 |
|  |  | 95 | 133.579 | 128.542 | 123.504 | 118.466 | 113.428 |
| MiBP (ng/mL) | Global | 10 | 22.875 | 22.441 | 22.007 | 21.573 | 21.139 |
|  |  | 25 | 26.457 | 27.614 | 28.772 | 29.929 | 31.087 |
|  |  | 50 | 30.436 | 33.362 | 36.288 | 39.214 | 42.140 |
|  |  | 75 | 34.416 | 39.110 | 43.804 | 48.498 | 53.193 |
|  |  | 95 | 40.141 | 47.380 | 54.618 | 61.856 | 69.094 |
|  | Latin America | 10 | 15.882 | 14.149 | 12.416 | 10.683 | 8.950 |
|  |  | 25 | 19.351 | 19.136 | 18.920 | 18.704 | 18.488 |
|  |  | 50 | 23.206 | 24.676 | 26.146 | 27.616 | 29.085 |
|  |  | 75 | 27.061 | 30.216 | 33.372 | 36.527 | 39.683 |
|  |  | 95 | 32.607 | 38.187 | 43.768 | 49.348 | 54.929 |
|  | Africa | 10 | 14.614 | 12.881 | 11.147 | 9.414 | **<LOD** |
|  |  | 25 | 22.989 | 22.773 | 22.557 | 22.341 | 22.125 |
|  |  | 50 | 32.294 | 33.764 | 35.234 | 36.704 | 38.173 |
|  |  | 75 | 41.600 | 44.755 | 47.911 | 51.066 | 54.222 |
|  |  | 95 | 54.987 | 60.568 | 66.149 | 71.729 | 77.310 |
|  | Asia - EPA | 10 | 26.018 | 24.285 | 22.552 | 20.819 | 19.086 |
|  |  | 25 | 30.467 | 30.252 | 30.036 | 29.820 | 29.604 |
|  |  | 50 | 35.411 | 36.881 | 38.351 | 39.821 | 41.290 |
|  |  | 75 | 40.355 | 43.511 | 46.666 | 49.821 | 52.977 |
|  |  | 95 | 47.468 | 53.048 | 58.629 | 64.209 | 69.790 |
|  | Asia - MESA | 10 | 35.882 | 34.149 | 32.416 | 30.683 | 28.950 |
|  |  | 25 | 41.982 | 41.766 | 41.550 | 41.334 | 41.118 |
|  |  | 50 | 48.758 | 50.228 | 51.698 | 53.168 | 54.638 |
|  |  | 75 | 55.535 | 58.691 | 61.846 | 65.002 | 68.157 |
|  |  | 95 | 65.285 | 70.866 | 76.446 | 82.027 | 87.607 |
|  | Australia | 10 | 22.038 | 20.305 | 18.571 | 16.838 | 15.105 |
|  |  | 25 | 23.719 | 23.503 | 23.287 | 23.072 | 22.856 |
|  |  | 50 | 25.588 | 27.058 | 28.527 | 29.997 | 31.467 |
|  |  | 75 | 27.456 | 30.612 | 33.767 | 36.923 | 40.078 |
|  |  | 95 | 30.144 | 35.725 | 41.305 | 46.886 | 52.466 |
| MnBP (ng/mL) | Global | 10 | 18.86 | 30.33 | 41.79 | 53.25 | 64.72 |
|  |  | 25 | 23.08 | 35.45 | 47.81 | 60.18 | 72.55 |
|  |  | 50 | 27.77 | 41.14 | 54.51 | 67.88 | 81.24 |
|  |  | 75 | 32.46 | 46.83 | 61.20 | 75.57 | 89.94 |
|  |  | 95 | 39.20 | 55.02 | 70.83 | 86.64 | 102.46 |
|  | Latin America | 10 | 15.23 | 26.54 | 37.84 | 49.15 | 60.46 |
|  |  | 25 | 16.96 | 29.13 | 41.29 | 53.46 | 65.63 |
|  |  | 50 | 18.88 | 32.00 | 45.13 | 58.26 | 71.38 |
|  |  | 75 | 20.79 | 34.88 | 48.96 | 63.05 | 77.13 |
|  |  | 95 | 23.55 | 39.02 | 54.48 | 69.94 | 85.40 |
|  | Africa | 10 | **<LOD** | **<LOD** | **<LOD** | **<LOD** | **<LOD** |
|  |  | 25 | 8.52 | 20.69 | 32.86 | 45.03 | 57.20 |
|  |  | 50 | 90.15 | 103.28 | 116.40 | 129.53 | 142.65 |
|  |  | 75 | 171.78 | 185.86 | 199.94 | 214.03 | 228.11 |
|  |  | 95 | 289.21 | 304.67 | 320.14 | 335.60 | 351.06 |
|  | Asia - EPA | 10 | 16.77 | 28.07 | 39.38 | 50.69 | 61.99 |
|  |  | 25 | 21.96 | 34.13 | 46.30 | 58.46 | 70.63 |
|  |  | 50 | 27.73 | 40.85 | 53.98 | 67.11 | 80.23 |
|  |  | 75 | 33.49 | 47.58 | 61.66 | 75.75 | 89.83 |
|  |  | 95 | 41.79 | 57.25 | 72.72 | 88.18 | 103.64 |
|  | Asia - MESA | 10 | 65.42 | 76.72 | 88.03 | 99.33 | 110.64 |
|  |  | 25 | 78.78 | 90.95 | 103.12 | 115.29 | 127.46 |
|  |  | 50 | 93.63 | 106.76 | 119.89 | 133.01 | 146.14 |
|  |  | 75 | 108.49 | 122.57 | 136.66 | 150.74 | 164.82 |
|  |  | 95 | 129.85 | 145.32 | 160.78 | 176.24 | 191.71 |
|  | Australia | 10 | **<LOD** | 1.76 | 13.06 | 24.37 | 35.67 |
|  |  | 25 | **<LOD** | 7.46 | 19.63 | 31.79 | 43.96 |
|  |  | 50 | 0.66 | 13.79 | 26.92 | 40.04 | 53.17 |
|  |  | 75 | 6.04 | 20.13 | 34.21 | 48.29 | 62.38 |
|  |  | 95 | 13.77 | 29.24 | 44.70 | 60.16 | 75.63 |
| MBzP (ng/mL) | Global | 10 | 4.569 | 3.516 | 2.463 | 1.409 | 0.356 |
|  |  | 25 | 5.195 | 4.033 | 2.871 | 1.709 | 0.547 |
|  |  | 50 | 5.891 | 4.608 | 3.326 | 2.043 | 0.761 |
|  |  | 75 | 6.586 | 5.183 | 3.780 | 2.377 | 0.974 |
|  |  | 95 | 7.586 | 6.010 | 4.433 | 2.857 | 1.280 |
|  | Latin America | 10 | 4.154 | 3.003 | 1.853 | 0.703 | **<LOD** |
|  |  | 25 | 4.699 | 3.428 | 2.156 | 0.884 | **<LOD** |
|  |  | 50 | 5.306 | 3.899 | 2.492 | 1.086 | **<LOD** |
|  |  | 75 | 5.912 | 4.370 | 2.829 | 1.287 | **<LOD** |
|  |  | 95 | 6.784 | 5.049 | 3.313 | 1.577 | **<LOD** |
|  | Africa | 10 | **<LOD** | **<LOD** | **<LOD** | **<LOD** | **<LOD** |
|  |  | 25 | 1.904 | 0.632 | **<LOD** | **<LOD** | **<LOD** |
|  |  | 50 | 4.722 | 3.315 | 1.908 | 0.502 | **<LOD** |
|  |  | 75 | 7.540 | 5.998 | 4.456 | 2.915 | 1.373 |
|  |  | 95 | 11.594 | 9.858 | 8.122 | 6.386 | 4.651 |
|  | Asia - EPA | 10 | 4.916 | 3.766 | 2.616 | 1.465 | 0.315 |
|  |  | 25 | 5.580 | 4.309 | 3.037 | 1.765 | 0.493 |
|  |  | 50 | 6.318 | 4.912 | 3.505 | 2.098 | 0.692 |
|  |  | 75 | 7.056 | 5.515 | 3.973 | 2.431 | 0.890 |
|  |  | 95 | 8.118 | 6.382 | 4.646 | 2.911 | 1.175 |
|  | Asia - MESA | 10 | 4.541 | 3.391 | 2.241 | 1.091 | -0.060 |
|  |  | 25 | 6.267 | 4.996 | 3.724 | 2.452 | 1.180 |
|  |  | 50 | 8.185 | 6.778 | 5.372 | 3.965 | 2.558 |
|  |  | 75 | 10.103 | 8.561 | 7.020 | 5.478 | 3.936 |
|  |  | 95 | 12.862 | 11.126 | 9.390 | 7.654 | 5.919 |
|  | Australia | 10 | 5.120 | 3.970 | 2.820 | 1.669 | 0.519 |
|  |  | 25 | 6.047 | 4.775 | 3.504 | 2.232 | 0.960 |
|  |  | 50 | 7.077 | 5.671 | 4.264 | 2.857 | 1.451 |
|  |  | 75 | 8.107 | 6.566 | 5.024 | 3.483 | 1.941 |
|  |  | 95 | 9.589 | 7.854 | 6.118 | 4.382 | 2.647 |

a. Predictive equations used to obtain the predicted values did not adjust for international regions.

b. Predictive equations used to obtain the predicted values were adjusted for international regions, pregnancy status, and age groups.

c. ND: Not detectable at the given time; <LOD: Value was below the limit of detection (pre-defined limit: 0.1 ng/mL).

Supplement Table 6: Estimated Quadratic Phthalate Concentrations, 2003 to 2023.

|  |  |  | **Estimate Concentrations per Year** | | | | |
| --- | --- | --- | --- | --- | --- | --- | --- |
| **Phthalate Polymers** | **Regions** | **Percentiles (%)** | **2003** | **2008** | **2013** | **2018** | **2023** |
| **High Molecular Weight** | | | | | | | |
| MEHP (ng/mL) | Global | 10 | 9.14 | 6.12 | 4.35 | 3.84 | 4.57 |
|  |  | 25 | 10.12 | 7.24 | 5.60 | 5.19 | 6.03 |
|  |  | 50 | 11.21 | 8.48 | 6.98 | 6.70 | 7.65 |
|  |  | 75 | 12.30 | 9.72 | 8.36 | 8.21 | 9.27 |
|  |  | 95 | 13.87 | 11.51 | 10.35 | 10.38 | 11.61 |
|  | Latin America | 10 | 8.32 | 4.94 | 2.95 | 2.33 | 3.11 |
|  |  | 25 | 8.89 | 5.49 | 3.54 | 3.02 | 3.93 |
|  |  | 50 | 9.53 | 6.11 | 4.19 | 3.77 | 4.86 |
|  |  | 75 | 10.16 | 6.72 | 4.85 | 4.53 | 5.78 |
|  |  | 95 | 11.07 | 7.61 | 5.79 | 5.62 | 7.10 |
|  | Africa | 10 | 0.712 | **<LOD** | **<LOD** | **<LOD** | **<LOD** |
|  |  | 25 | 5.000 | 1.601 | **<LOD** | **<LOD** | **<LOD** |
|  |  | 50 | 9.764 | 6.347 | 4.429 | 4.011 | 5.094 |
|  |  | 75 | 14.529 | 11.092 | 9.216 | 8.900 | 10.146 |
|  |  | 95 | 21.383 | 17.919 | 16.103 | 15.935 | 17.414 |
|  | Asia - EPA | 10 | 9.673 | 6.291 | 4.294 | 3.682 | 4.455 |
|  |  | 25 | 11.045 | 7.646 | 5.686 | 5.166 | 6.086 |
|  |  | 50 | 12.569 | 9.151 | 7.233 | 6.815 | 7.898 |
|  |  | 75 | 14.093 | 10.656 | 8.780 | 8.465 | 9.710 |
|  |  | 95 | 16.285 | 12.821 | 11.005 | 10.837 | 12.317 |
|  | Asia - MESA | 10 | 15.115 | 11.733 | 9.736 | 9.124 | 9.896 |
|  |  | 25 | 18.708 | 15.309 | 13.349 | 12.829 | 13.749 |
|  |  | 50 | 22.700 | 19.282 | 17.365 | 16.947 | 18.029 |
|  |  | 75 | 26.693 | 23.256 | 21.380 | 21.065 | 22.310 |
|  |  | 95 | 32.437 | 28.972 | 27.156 | 26.988 | 28.468 |
|  | Australia | 10 | **ND** | **ND** | **ND** | **ND** | **ND** |
|  |  | 25 | **ND** | **ND** | **ND** | **ND** | **ND** |
|  |  | 50 | **ND** | **ND** | **ND** | **ND** | **ND** |
|  |  | 75 | **ND** | **ND** | **ND** | **ND** | **ND** |
|  |  | 95 | **ND** | **ND** | **ND** | **ND** | **ND** |
| MEHHP (ng/mL) | Global | 10 | 25.68 | 16.12 | 11.52 | 11.88 | 17.20 |
|  |  | 25 | 28.03 | 18.56 | 14.19 | 14.94 | 20.80 |
|  |  | 50 | 30.65 | 21.27 | 17.16 | 18.34 | 24.80 |
|  |  | 75 | 33.27 | 23.97 | 20.13 | 21.74 | 28.80 |
|  |  | 95 | 37.04 | 27.87 | 24.40 | 26.63 | 34.56 |
|  | Latin America | 10 | 23.83 | 13.90 | 8.77 | 8.47 | 12.98 |
|  |  | 25 | 26.47 | 16.61 | 11.72 | 11.81 | 16.87 |
|  |  | 50 | 29.40 | 19.61 | 14.99 | 15.51 | 21.19 |
|  |  | 75 | 32.32 | 22.62 | 18.26 | 19.22 | 25.52 |
|  |  | 95 | 36.53 | 26.95 | 22.96 | 24.56 | 31.74 |
|  | Africa | 10 | **<LOD** | **<LOD** | **<LOD** | **<LOD** | **<LOD** |
|  |  | 25 | **<LOD** | **<LOD** | **<LOD** | **<LOD** | **<LOD** |
|  |  | 50 | 35.13 | 25.34 | 20.72 | 21.24 | 26.92 |
|  |  | 75 | 115.54 | 105.84 | 101.47 | 102.44 | 108.74 |
|  |  | 95 | 231.23 | 221.65 | 217.65 | 219.25 | 226.44 |
|  | Asia - EPA | 10 | 26.81 | 16.87 | 11.75 | 11.44 | 15.95 |
|  |  | 25 | 29.06 | 19.20 | 14.31 | 14.40 | 19.46 |
|  |  | 50 | 31.57 | 21.79 | 17.16 | 17.69 | 23.37 |
|  |  | 75 | 34.07 | 24.37 | 20.01 | 20.97 | 27.27 |
|  |  | 95 | 37.68 | 28.10 | 24.10 | 25.70 | 32.88 |
|  | Asia - MESA | 10 | 46.00 | 36.06 | 30.94 | 30.63 | 35.14 |
|  |  | 25 | 51.00 | 41.14 | 36.25 | 36.34 | 41.40 |
|  |  | 50 | 56.56 | 46.77 | 42.15 | 42.67 | 48.35 |
|  |  | 75 | 62.11 | 52.41 | 48.05 | 49.01 | 55.31 |
|  |  | 95 | 70.11 | 60.53 | 56.53 | 58.13 | 65.32 |
|  | Australia | 10 | 24.80 | 14.86 | 9.74 | 9.43 | 13.94 |
|  |  | 25 | 28.65 | 18.79 | 13.90 | 13.99 | 19.05 |
|  |  | 50 | 32.94 | 23.15 | 18.53 | 19.05 | 24.73 |
|  |  | 75 | 37.22 | 27.52 | 23.15 | 24.12 | 30.42 |
|  |  | 95 | 43.38 | 33.80 | 29.81 | 31.40 | 38.59 |
| MEOHP (ng/mL) | Global | 10 | 16.882 | 11.947 | 7.936 | 4.850 | 2.688 |
|  |  | 25 | 18.820 | 13.750 | 9.855 | 7.134 | 5.588 |
|  |  | 50 | 20.974 | 15.754 | 11.987 | 9.672 | 8.809 |
|  |  | 75 | 23.128 | 17.758 | 14.119 | 12.210 | 12.031 |
|  |  | 95 | 26.226 | 20.641 | 17.186 | 15.861 | 16.665 |
|  | Latin America | 10 | 21.452 | 16.131 | 11.822 | 8.527 | 6.244 |
|  |  | 25 | 23.406 | 17.870 | 13.626 | 10.675 | 9.016 |
|  |  | 50 | 25.578 | 19.803 | 15.631 | 13.062 | 12.096 |
|  |  | 75 | 27.750 | 21.736 | 17.636 | 15.449 | 15.176 |
|  |  | 95 | 30.874 | 24.517 | 20.520 | 18.883 | 19.607 |
|  | Africa | 10 | **<LOD** | **<LOD** | **<LOD** | **<LOD** | **<LOD** |
|  |  | 25 | 8.665 | 3.221 | **<LOD** | **<LOD** | **<LOD** |
|  |  | 50 | 25.788 | 20.106 | 15.887 | 13.132 | 11.841 |
|  |  | 75 | 42.911 | 36.990 | 32.843 | 30.471 | 29.873 |
|  |  | 95 | 67.546 | 61.281 | 57.238 | 55.415 | 55.814 |
|  | Asia - EPA | 10 | 17.891 | 12.570 | 8.262 | 4.966 | 2.683 |
|  |  | 25 | 19.951 | 14.415 | 10.171 | 7.220 | 5.561 |
|  |  | 50 | 22.240 | 16.465 | 12.293 | 9.724 | 8.758 |
|  |  | 75 | 24.529 | 18.515 | 14.415 | 12.228 | 11.955 |
|  |  | 95 | 27.821 | 21.464 | 17.467 | 15.830 | 16.554 |
|  | Asia - MESA | 10 | 25.500 | 20.179 | 15.871 | 12.575 | 10.293 |
|  |  | 25 | 30.152 | 24.615 | 20.372 | 17.420 | 15.761 |
|  |  | 50 | 35.320 | 29.545 | 25.373 | 22.804 | 21.838 |
|  |  | 75 | 40.488 | 34.474 | 30.374 | 28.187 | 27.914 |
|  |  | 95 | 47.923 | 41.565 | 37.568 | 35.932 | 36.655 |
|  | Australia | 10 | 16.851 | 11.529 | 7.221 | 3.926 | 1.643 |
|  |  | 25 | 19.754 | 14.217 | 9.974 | 7.022 | 5.363 |
|  |  | 50 | 22.979 | 17.204 | 13.032 | 10.463 | 9.497 |
|  |  | 75 | 26.204 | 20.190 | 16.090 | 13.903 | 13.630 |
|  |  | 95 | 30.844 | 24.487 | 20.490 | 18.853 | 19.577 |
| MECPP (ng/mL) | Global | 10 | 43.05 | 28.43 | 20.13 | 18.14 | 22.47 |
|  |  | 25 | 45.15 | 31.36 | 23.81 | 22.50 | 27.43 |
|  |  | 50 | 47.48 | 34.62 | 27.91 | 27.35 | 32.94 |
|  |  | 75 | 49.82 | 37.88 | 32.01 | 32.19 | 38.45 |
|  |  | 95 | 53.17 | 42.57 | 37.90 | 39.17 | 46.37 |
|  | Latin America | 10 | 42.82 | 27.52 | 18.72 | 16.43 | 20.64 |
|  |  | 25 | 46.70 | 34.04 | 26.95 | 25.43 | 29.48 |
|  |  | 50 | 51.01 | 41.28 | 36.08 | 35.43 | 39.31 |
|  |  | 75 | 55.31 | 48.51 | 45.22 | 45.43 | 49.13 |
|  |  | 95 | 61.51 | 58.93 | 58.36 | 59.81 | 63.27 |
|  | Africa | 10 | **<LOD** | **<LOD** | **<LOD** | **<LOD** | **<LOD** |
|  |  | 25 | 33.71 | 21.05 | 13.96 | 12.44 | 16.50 |
|  |  | 50 | 83.30 | 73.57 | 68.38 | 67.72 | 71.60 |
|  |  | 75 | 132.89 | 126.09 | 122.79 | 123.00 | 126.71 |
|  |  | 95 | 204.23 | 201.65 | 201.08 | 202.53 | 205.99 |
|  | Asia - EPA | 10 | **ND** | 30.67 | 21.88 | 19.58 | 23.79 |
|  |  | 25 | **ND** | 31.16 | 24.06 | 22.55 | 26.60 |
|  |  | 50 | **ND** | 31.69 | 26.50 | 25.84 | 29.72 |
|  |  | 75 | **ND** | 32.23 | 28.93 | 29.14 | 32.84 |
|  |  | 95 | **ND** | 32.99 | 32.43 | 33.88 | 37.34 |
|  | Asia - MESA | 10 | 49.73 | 34.43 | 25.63 | 23.33 | 27.54 |
|  |  | 25 | 50.82 | 38.16 | 31.07 | 29.55 | 33.61 |
|  |  | 50 | 52.04 | 42.31 | 37.12 | 36.46 | 40.35 |
|  |  | 75 | 53.26 | 46.47 | 43.17 | 43.38 | 47.09 |
|  |  | 95 | 55.02 | 52.44 | 51.87 | 53.32 | 56.78 |
|  | Australia | 10 | 37.51 | 22.21 | 13.41 | 11.12 | 15.32 |
|  |  | 25 | 38.18 | 25.52 | 18.43 | 16.91 | 20.96 |
|  |  | 50 | 38.93 | 29.20 | 24.01 | 23.35 | 27.23 |
|  |  | 75 | 39.68 | 32.88 | 29.59 | 29.79 | 33.50 |
|  |  | 95 | 40.76 | 38.18 | 37.61 | 39.06 | 42.52 |
| MCMHP (ng/mL) | Global | 10 | 36.90 | 16.53 | 6.57 | 7.03 | 17.91 |
|  |  | 25 | 51.17 | 23.87 | 10.45 | 10.90 | 25.23 |
|  |  | 50 | 67.03 | 32.03 | 14.75 | 15.20 | 33.36 |
|  |  | 75 | 82.88 | 40.19 | 19.06 | 19.49 | 41.50 |
|  |  | 95 | 105.69 | 51.92 | 25.25 | 25.68 | 53.20 |
|  | Latin America | 10 | 38.19 | 18.39 | 8.56 | 8.71 | 18.84 |
|  |  | 25 | 57.85 | 31.87 | 19.00 | 19.26 | 32.63 |
|  |  | 50 | 79.71 | 46.85 | 30.60 | 30.97 | 47.94 |
|  |  | 75 | 101.56 | 61.83 | 42.21 | 42.68 | 63.26 |
|  |  | 95 | 132.99 | 83.38 | 58.90 | 59.54 | 85.30 |
|  | Africa | 10 | **ND** | **ND** | **ND** | **ND** | **ND** |
|  |  | 25 | **ND** | **ND** | **ND** | **ND** | **ND** |
|  |  | 50 | **ND** | **ND** | **ND** | **ND** | **ND** |
|  |  | 75 | **ND** | **ND** | **ND** | **ND** | **ND** |
|  |  | 95 | **ND** | **ND** | **ND** | **ND** | **ND** |
|  | Asia - EPA | 10 | 36.33 | 16.53 | 6.70 | 6.86 | 16.98 |
|  |  | 25 | 49.48 | 23.50 | 10.63 | 10.88 | 24.25 |
|  |  | 50 | 64.09 | 31.24 | 14.99 | 15.36 | 32.33 |
|  |  | 75 | 78.70 | 38.98 | 19.35 | 19.83 | 40.41 |
|  |  | 95 | 99.72 | 50.11 | 25.63 | 26.27 | 52.03 |
|  | Asia - MESA | 10 | 40.09 | 20.29 | 10.47 | 10.62 | 20.75 |
|  |  | 25 | 54.67 | 28.69 | 15.82 | 16.08 | 29.45 |
|  |  | 50 | 70.88 | 38.02 | 21.78 | 22.14 | 39.12 |
|  |  | 75 | 87.08 | 47.35 | 27.73 | 28.21 | 48.79 |
|  |  | 95 | 110.39 | 60.78 | 36.29 | 36.93 | 62.70 |
|  | Australia | 10 | **ND** | **ND** | **ND** | **ND** | **ND** |
|  |  | 25 | **ND** | **ND** | **ND** | **ND** | **ND** |
|  |  | 50 | **ND** | **ND** | **ND** | **ND** | **ND** |
|  |  | 75 | **ND** | **ND** | **ND** | **ND** | **ND** |
|  |  | 95 | **ND** | **ND** | **ND** | **ND** | **ND** |
| MCOP (ng/mL) | Global | 10 | 3.74 | 5.12 | 3.79 | **<LOD** | **ND** |
|  |  | 25 | 4.63 | 5.68 | 4.08 | **<LOD** | **ND** |
|  |  | 50 | 5.62 | 6.29 | 4.40 | **<LOD** | **ND** |
|  |  | 75 | 6.60 | 6.91 | 4.72 | **<LOD** | **ND** |
|  |  | 95 | 8.03 | 7.80 | 5.18 | 0.16 | **ND** |
|  | Latin America | 10 | 4.71 | 4.78 | 3.01 | **<LOD** | **<LOD** |
|  |  | 25 | 4.90 | 5.43 | 3.99 | 0.57 | **<LOD** |
|  |  | 50 | 5.11 | 6.15 | 5.07 | 1.85 | **<LOD** |
|  |  | 75 | 5.32 | 6.88 | 6.15 | 3.14 | **<LOD** |
|  |  | 95 | 5.62 | 7.91 | 7.71 | 5.00 | **<LOD** |
|  | Africa | 10 | **<LOD** | **<LOD** | **<LOD** | **<LOD** | **<LOD** |
|  |  | 25 | **<LOD** | **<LOD** | **<LOD** | **<LOD** | **<LOD** |
|  |  | 50 | 1.10 | 2.14 | 1.05 | **<LOD** | **<LOD** |
|  |  | 75 | 2.64 | 4.19 | 3.47 | 0.46 | **<LOD** |
|  |  | 95 | 4.86 | 7.15 | 6.94 | 4.24 | **<LOD** |
|  | Asia - EPA | 10 | **ND** | **ND** | **ND** | **<LOD** | **<LOD** |
|  |  | 25 | **ND** | **ND** | **ND** | **<LOD** | **<LOD** |
|  |  | 50 | **ND** | **ND** | **ND** | **<LOD** | **<LOD** |
|  |  | 75 | **ND** | **ND** | **ND** | **<LOD** | **<LOD** |
|  |  | 95 | **ND** | **ND** | **ND** | 0.13 | **<LOD** |
|  | Asia - MESA | 10 | 1.71 | 1.78 | **<LOD** | **<LOD** | **<LOD** |
|  |  | 25 | 2.61 | 3.14 | 1.69 | **<LOD** | **<LOD** |
|  |  | 50 | 3.61 | 4.65 | 3.57 | 0.36 | **<LOD** |
|  |  | 75 | 4.61 | 6.17 | 5.44 | 2.44 | **<LOD** |
|  |  | 95 | 6.06 | 8.35 | 8.14 | 5.43 | 0.23 |
|  | Australia | 10 | **ND** | **ND** | **ND** | **ND** | **ND** |
|  |  | 25 | **ND** | **ND** | **ND** | **ND** | **ND** |
|  |  | 50 | **ND** | **ND** | **ND** | **ND** | **ND** |
|  |  | 75 | **ND** | **ND** | **ND** | **ND** | **ND** |
|  |  | 95 | **ND** | **ND** | **ND** | **ND** | **ND** |
| MiNP (ng/mL) | Global | 10 | 2.489 | 0.593 | 0.052 | 0.866 | 3.035 |
|  |  | 25 | 2.721 | 0.725 | 0.182 | 1.090 | 3.452 |
|  |  | 50 | 2.978 | 0.871 | 0.325 | 1.339 | 3.914 |
|  |  | 75 | 3.236 | 1.018 | 0.469 | 1.588 | 4.377 |
|  |  | 95 | 3.606 | 1.228 | 0.675 | 1.947 | 5.042 |
|  | Latin America | 10 | 1.576 | **<LOD** | **<LOD** | 0.083 | 2.329 |
|  |  | 25 | 1.651 | **<LOD** | **<LOD** | 0.178 | 2.630 |
|  |  | 50 | 1.735 | **<LOD** | **<LOD** | 0.284 | 2.964 |
|  |  | 75 | 1.819 | **<LOD** | **<LOD** | 0.389 | 3.298 |
|  |  | 95 | 1.939 | **<LOD** | **<LOD** | 0.541 | 3.778 |
|  | Africa | 10 | **ND** | **ND** | **ND** | **ND** | **ND** |
|  |  | 25 | **ND** | **ND** | **ND** | **ND** | **ND** |
|  |  | 50 | **ND** | **ND** | **ND** | **ND** | **ND** |
|  |  | 75 | **ND** | **ND** | **ND** | **ND** | **ND** |
|  |  | 95 | **ND** | **ND** | **ND** | **ND** | **ND** |
|  | Asia - EPA | 10 | 2.354 | 0.484 | -0.014 | 0.860 | 3.106 |
|  |  | 25 | 2.560 | 0.598 | 0.106 | 1.087 | 3.538 |
|  |  | 50 | 2.790 | 0.724 | 0.240 | 1.338 | 4.018 |
|  |  | 75 | 3.019 | 0.850 | 0.374 | 1.590 | 4.498 |
|  |  | 95 | 3.349 | 1.032 | 0.566 | 1.951 | 5.188 |
|  | Asia - MESA | 10 | **<LOD** | **<LOD** | **<LOD** | **<LOD** | **<LOD** |
|  |  | 25 | **<LOD** | **<LOD** | **<LOD** | **<LOD** | **<LOD** |
|  |  | 50 | 7.417 | 5.351 | 4.867 | 5.965 | 8.645 |
|  |  | 75 | 23.840 | 21.671 | 21.195 | 22.411 | 25.319 |
|  |  | 95 | 47.468 | 45.151 | 44.685 | 46.071 | 49.308 |
|  | Australia | 10 | **ND** | **ND** | **ND** | **ND** | **ND** |
|  |  | 25 | **ND** | **ND** | **ND** | **ND** | **ND** |
|  |  | 50 | **ND** | **ND** | **ND** | **ND** | **ND** |
|  |  | 75 | **ND** | **ND** | **ND** | **ND** | **ND** |
|  |  | 95 | **ND** | **ND** | **ND** | **ND** | **ND** |
| MOiNP (ng/mL) | Global | 10 | 3.294 | 3.820 | 2.260 | **<LOD** | **<LOD** |
|  |  | 25 | 3.712 | 4.514 | 3.611 | 1.000 | **<LOD** |
|  |  | 50 | 4.175 | 5.286 | 5.112 | 3.654 | 0.912 |
|  |  | 75 | 4.639 | 6.057 | 6.613 | 6.307 | 5.140 |
|  |  | 95 | 5.306 | 7.167 | 8.773 | 10.124 | 11.222 |
|  | Latin America | 10 | **ND** | **ND** | **ND** | **ND** | **ND** |
|  |  | 25 | **ND** | **ND** | **ND** | **ND** | **ND** |
|  |  | 50 | **ND** | **ND** | **ND** | **ND** | **ND** |
|  |  | 75 | **ND** | **ND** | **ND** | **ND** | **ND** |
|  |  | 95 | **ND** | **ND** | **ND** | **ND** | **ND** |
|  | Africa | 10 | **ND** | **ND** | **ND** | **ND** | **ND** |
|  |  | 25 | **ND** | **ND** | **ND** | **ND** | **ND** |
|  |  | 50 | **ND** | **ND** | **ND** | **ND** | **ND** |
|  |  | 75 | **ND** | **ND** | **ND** | **ND** | **ND** |
|  |  | 95 | **ND** | **ND** | **ND** | **ND** | **ND** |
|  | Asia - EPA | 10 | 2.823 | 4.135 | 2.821 | **<LOD** | **<LOD** |
|  |  | 25 | 3.947 | 4.432 | 3.334 | 0.753 | **<LOD** |
|  |  | 50 | 5.195 | 4.761 | 3.905 | 2.611 | 0.879 |
|  |  | 75 | 6.443 | 5.091 | 4.475 | 4.468 | 5.070 |
|  |  | 95 | 8.239 | 5.566 | 5.296 | 7.141 | 11.099 |
|  | Asia - MESA | 10 | **<LOD** | 0.798 | **<LOD** | **<LOD** | **<LOD** |
|  |  | 25 | 4.587 | 5.072 | 3.975 | 1.394 | **<LOD** |
|  |  | 50 | 10.255 | 9.821 | 8.965 | 7.671 | 5.939 |
|  |  | 75 | 15.923 | 14.571 | 13.955 | 13.948 | 14.549 |
|  |  | 95 | 24.077 | 21.403 | 21.134 | 22.978 | 26.937 |
|  | Australia | 10 | **ND** | **ND** | **ND** | **ND** | **ND** |
|  |  | 25 | **ND** | **ND** | **ND** | **ND** | **ND** |
|  |  | 50 | **ND** | **ND** | **ND** | **ND** | **ND** |
|  |  | 75 | **ND** | **ND** | **ND** | **ND** | **ND** |
|  |  | 95 | **ND** | **ND** | **ND** | **ND** | **ND** |
| OH-MiNP (ng/mL) | Global | 10 | 4.788 | 6.418 | 4.676 | **<LOD** | **<LOD** |
|  |  | 25 | 5.674 | 6.707 | 5.535 | 2.155 | **<LOD** |
|  |  | 50 | 6.658 | 7.029 | 6.489 | 5.038 | 2.676 |
|  |  | 75 | 7.642 | 7.350 | 7.443 | 7.921 | 8.783 |
|  |  | 95 | 9.058 | 7.813 | 8.816 | 12.068 | 17.568 |
|  | Latin America | 10 | **ND** | **ND** | **ND** | **ND** | **ND** |
|  |  | 25 | **ND** | **ND** | **ND** | **ND** | **ND** |
|  |  | 50 | **ND** | **ND** | **ND** | **ND** | **ND** |
|  |  | 75 | **ND** | **ND** | **ND** | **ND** | **ND** |
|  |  | 95 | **ND** | **ND** | **ND** | **ND** | **ND** |
|  | Africa | 10 | **ND** | **ND** | **ND** | **ND** | **ND** |
|  |  | 25 | **ND** | **ND** | **ND** | **ND** | **ND** |
|  |  | 50 | **ND** | **ND** | **ND** | **ND** | **ND** |
|  |  | 75 | **ND** | **ND** | **ND** | **ND** | **ND** |
|  |  | 95 | **ND** | **ND** | **ND** | **ND** | **ND** |
|  | Asia - EPA | 10 | 4.698 | 6.372 | 4.219 | **<LOD** | **<LOD** |
|  |  | 25 | 5.875 | 6.550 | 4.971 | 1.395 | **<LOD** |
|  |  | 50 | 7.182 | 6.748 | 5.806 | 4.338 | 2.343 |
|  |  | 75 | 8.489 | 6.947 | 6.642 | 7.281 | 8.864 |
|  |  | 95 | 10.369 | 7.231 | 7.843 | 11.515 | 18.247 |
|  | Asia - MESA | 10 | **<LOD** | 1.582 | **<LOD** | **<LOD** | **<LOD** |
|  |  | 25 | 6.194 | 6.870 | 5.290 | 1.714 | **<LOD** |
|  |  | 50 | 13.178 | 12.745 | 11.803 | 10.334 | 8.339 |
|  |  | 75 | 20.163 | 18.620 | 18.315 | 18.955 | 20.538 |
|  |  | 95 | 30.211 | 27.073 | 27.685 | 31.357 | 38.088 |
|  | Australia | 10 | **ND** | **ND** | **ND** | **ND** | **ND** |
|  |  | 25 | **ND** | **ND** | **ND** | **ND** | **ND** |
|  |  | 50 | **ND** | **ND** | **ND** | **ND** | **ND** |
|  |  | 75 | **ND** | **ND** | **ND** | **ND** | **ND** |
|  |  | 95 | **ND** | **ND** | **ND** | **ND** | **ND** |
| MCiNP (ng/mL) | Global | 10 | **ND** | **ND** | **<LOD** | 0.819 | **ND** |
|  |  | 25 | **ND** | **ND** | **<LOD** | 0.833 | **ND** |
|  |  | 50 | **ND** | **ND** | 0.102 | 0.847 | **ND** |
|  |  | 75 | **ND** | **ND** | 0.181 | 0.862 | **ND** |
|  |  | 95 | **ND** | **ND** | 0.294 | 0.883 | **ND** |
|  | Latin America | 10 | **ND** | **ND** | **ND** | **<LOD** | **<LOD** |
|  |  | 25 | **ND** | **ND** | **ND** | **<LOD** | **<LOD** |
|  |  | 50 | **ND** | **ND** | **ND** | **<LOD** | **<LOD** |
|  |  | 75 | **ND** | **ND** | **ND** | **<LOD** | **<LOD** |
|  |  | 95 | **ND** | **ND** | **ND** | 0.500 | 0.836 |
|  | Africa | 10 | **ND** | **<LOD** | **<LOD** | **ND** | **ND** |
|  |  | 25 | 0.258 | **<LOD** | **<LOD** | **ND** | **ND** |
|  |  | 50 | 1.536 | 1.131 | 0.374 | **ND** | **ND** |
|  |  | 75 | 2.814 | 2.432 | 1.936 | **ND** | **ND** |
|  |  | 95 | 4.652 | 4.304 | 4.184 | **ND** | **ND** |
|  | Asia - EPA | 10 | **ND** | **ND** | **ND** | **<LOD** | **<LOD** |
|  |  | 25 | **ND** | **ND** | **ND** | **<LOD** | **<LOD** |
|  |  | 50 | **ND** | **ND** | **ND** | **<LOD** | **<LOD** |
|  |  | 75 | **ND** | **ND** | **ND** | **<LOD** | **<LOD** |
|  |  | 95 | **ND** | **ND** | **ND** | 0.256 | 0.592 |
|  | Asia - MESA | 10 | 10.037 | 9.587 | 8.334 | 6.279 | 3.421 |
|  |  | 25 | 12.724 | 12.296 | 11.278 | 9.671 | 7.476 |
|  |  | 50 | 15.711 | 15.305 | 14.549 | 13.441 | 11.981 |
|  |  | 75 | 18.697 | 18.315 | 17.819 | 17.210 | 16.487 |
|  |  | 95 | 22.993 | 22.645 | 22.525 | 22.633 | 22.969 |
|  | Australia | 10 | **ND** | **ND** | **ND** | **ND** | **ND** |
|  |  | 25 | **ND** | **ND** | **ND** | **ND** | **ND** |
|  |  | 50 | **ND** | **ND** | **ND** | **ND** | **ND** |
|  |  | 75 | **ND** | **ND** | **ND** | **ND** | **ND** |
|  |  | 95 | **ND** | **ND** | **ND** | **ND** | **ND** |
| MOP (ng/mL) | Global | 10 | **ND** | 0.979 | 0.701 | 0.662 | **ND** |
|  |  | 25 | **ND** | 1.058 | 1.077 | 0.879 | **ND** |
|  |  | 50 | **ND** | 1.146 | 1.496 | 1.120 | **ND** |
|  |  | 75 | **ND** | 1.233 | 1.914 | 1.361 | **ND** |
|  |  | 95 | **ND** | 1.359 | 2.516 | 1.708 | **ND** |
|  | Latin America | 10 | **ND** | **ND** | 0.308 | 0.406 | **ND** |
|  |  | 25 | **ND** | **ND** | 0.512 | 0.488 | **ND** |
|  |  | 50 | **ND** | **ND** | 0.739 | 0.580 | **ND** |
|  |  | 75 | **ND** | **ND** | 0.966 | 0.671 | **ND** |
|  |  | 95 | **ND** | **ND** | 1.292 | 0.803 | **ND** |
|  | Africa | 10 | **ND** | **ND** | **ND** | **ND** | **ND** |
|  |  | 25 | **ND** | **ND** | **ND** | **ND** | **ND** |
|  |  | 50 | **ND** | **ND** | **ND** | **ND** | **ND** |
|  |  | 75 | **ND** | **ND** | **ND** | **ND** | **ND** |
|  |  | 95 | **ND** | **ND** | **ND** | **ND** | **ND** |
|  | Asia - EPA | 10 | **ND** | **ND** | **<LOD** | **<LOD** | **ND** |
|  |  | 25 | **ND** | **ND** | **<LOD** | **<LOD** | **ND** |
|  |  | 50 | **ND** | **ND** | 0.341 | 0.182 | **ND** |
|  |  | 75 | **ND** | **ND** | 0.628 | 0.334 | **ND** |
|  |  | 95 | **ND** | **ND** | 1.042 | 0.552 | **ND** |
|  | Asia - MESA | 10 | 0.380 | **<LOD** | **<LOD** | **<LOD** | **<LOD** |
|  |  | 25 | 2.208 | 1.990 | 1.869 | 1.845 | 1.919 |
|  |  | 50 | 4.240 | 4.769 | 4.954 | 4.795 | 4.291 |
|  |  | 75 | 6.271 | 7.548 | 8.040 | 7.745 | 6.664 |
|  |  | 95 | 9.193 | 11.547 | 12.479 | 11.989 | 10.078 |
|  | Australia | 10 | **ND** | **ND** | **ND** | **ND** | **ND** |
|  |  | 25 | **ND** | **ND** | **ND** | **ND** | **ND** |
|  |  | 50 | **ND** | **ND** | **ND** | **ND** | **ND** |
|  |  | 75 | **ND** | **ND** | **ND** | **ND** | **ND** |
|  |  | 95 | **ND** | **ND** | **ND** | **ND** | **ND** |
| MCPP (ng/mL) | Global | 10 | **ND** | 1.501 | 1.291 | 0.573 | **ND** |
|  |  | 25 | **ND** | 1.640 | 1.481 | 0.704 | **ND** |
|  |  | 50 | **ND** | 1.793 | 1.691 | 0.850 | **ND** |
|  |  | 75 | **ND** | 1.947 | 1.902 | 0.996 | **ND** |
|  |  | 95 | **ND** | 2.168 | 2.204 | 1.205 | **ND** |
|  | Latin America | 10 | **ND** | 1.688 | 1.041 | **<LOD** | **<LOD** |
|  |  | 25 | **ND** | 1.714 | 1.146 | **<LOD** | **<LOD** |
|  |  | 50 | **ND** | 1.743 | 1.261 | **<LOD** | **<LOD** |
|  |  | 75 | **ND** | 1.771 | 1.377 | **<LOD** | **<LOD** |
|  |  | 95 | **ND** | 1.813 | 1.543 | 0.293 | **<LOD** |
|  | Africa | 10 | **ND** | **ND** | **ND** | **ND** | **ND** |
|  |  | 25 | **ND** | **ND** | **ND** | **ND** | **ND** |
|  |  | 50 | **ND** | **ND** | **ND** | **ND** | **ND** |
|  |  | 75 | **ND** | **ND** | **ND** | **ND** | **ND** |
|  |  | 95 | **ND** | **ND** | **ND** | **ND** | **ND** |
|  | Asia - EPA | 10 | **ND** | **ND** | **ND** | 0.943 | **<LOD** |
|  |  | 25 | **ND** | **ND** | **ND** | 0.961 | **<LOD** |
|  |  | 50 | **ND** | **ND** | **ND** | 0.982 | **<LOD** |
|  |  | 75 | **ND** | **ND** | **ND** | 1.003 | **<LOD** |
|  |  | 95 | **ND** | **ND** | **ND** | 1.033 | **<LOD** |
|  | Asia - MESA | 10 | **ND** | **ND** | **ND** | **ND** | **<LOD** |
|  |  | 25 | **ND** | **ND** | **ND** | **ND** | **<LOD** |
|  |  | 50 | **ND** | **ND** | **ND** | **ND** | **<LOD** |
|  |  | 75 | **ND** | **ND** | **ND** | **ND** | **<LOD** |
|  |  | 95 | **ND** | **ND** | **ND** | **ND** | **<LOD** |
|  | Australia | 10 | **<LOD** | **<LOD** | **<LOD** | **<LOD** | **<LOD** |
|  |  | 25 | **<LOD** | **<LOD** | **<LOD** | **<LOD** | **<LOD** |
|  |  | 50 | 0.809 | 1.267 | 0.786 | **<LOD** | **<LOD** |
|  |  | 75 | 2.206 | 2.769 | 2.374 | 1.023 | **<LOD** |
|  |  | 95 | 4.217 | 4.929 | 4.660 | 3.409 | 1.177 |
| MiDP (ng/mL) | Global | 10 | #REF! | **ND** | **ND** | **<LOD** | **<LOD** |
|  |  | 25 | #REF! | **ND** | **ND** | #REF! | #REF! |
|  |  | 50 | #REF! | **ND** | **ND** | #REF! | #REF! |
|  |  | 75 | #REF! | **ND** | **ND** | #REF! | #REF! |
|  |  | 95 | #REF! | **ND** | **ND** | #REF! | #REF! |
|  | Latin America | 10 | **ND** | **ND** | **ND** | **ND** | **ND** |
|  |  | 25 | **ND** | **ND** | **ND** | **ND** | **ND** |
|  |  | 50 | **ND** | **ND** | **ND** | **ND** | **ND** |
|  |  | 75 | **ND** | **ND** | **ND** | **ND** | **ND** |
|  |  | 95 | **ND** | **ND** | **ND** | **ND** | **ND** |
|  | Africa | 10 | **ND** | **ND** | **ND** | **ND** | **ND** |
|  |  | 25 | **ND** | **ND** | **ND** | **ND** | **ND** |
|  |  | 50 | **ND** | **ND** | **ND** | **ND** | **ND** |
|  |  | 75 | **ND** | **ND** | **ND** | **ND** | **ND** |
|  |  | 95 | **ND** | **ND** | **ND** | **ND** | **ND** |
|  | Asia - EPA | 10 | **<LOD** | **<LOD** | **ND** | **<LOD** | **<LOD** |
|  |  | 25 | **<LOD** | **<LOD** | **ND** | 1.243 | 2.636 |
|  |  | 50 | 14.372 | 2.998 | **ND** | 4.109 | 16.594 |
|  |  | 75 | 28.800 | 6.099 | **ND** | 6.974 | 30.551 |
|  |  | 95 | 49.558 | 10.560 | **ND** | 11.097 | 50.632 |
|  | Asia - MESA | 10 | **<LOD** | **ND** | **ND** | **<LOD** | **<LOD** |
|  |  | 25 | **<LOD** | **ND** | **ND** | 1.210 | 2.603 |
|  |  | 50 | 14.314 | **ND** | **ND** | 4.051 | 16.536 |
|  |  | 75 | 28.718 | **ND** | **ND** | 6.892 | 30.469 |
|  |  | 95 | 49.439 | **ND** | **ND** | 10.979 | 50.514 |
|  | Australia | 10 | **ND** | **ND** | **ND** | **ND** | **ND** |
|  |  | 25 | **ND** | **ND** | **ND** | **ND** | **ND** |
|  |  | 50 | **ND** | **ND** | **ND** | **ND** | **ND** |
|  |  | 75 | **ND** | **ND** | **ND** | **ND** | **ND** |
|  |  | 95 | **ND** | **ND** | **ND** | **ND** | **ND** |
| **Low Molecular Weight** | | | | | | | |
| MEP (ng/mL) | Global | 10 | 56.691 | 34.212 | 21.113 | 17.394 | 23.055 |
|  |  | 25 | 60.980 | 38.962 | 26.335 | 23.098 | 29.252 |
|  |  | 50 | 65.745 | 44.240 | 32.136 | 29.435 | 36.137 |
|  |  | 75 | 70.510 | 49.517 | 37.938 | 35.773 | 43.022 |
|  |  | 95 | 77.366 | 57.110 | 46.285 | 44.891 | 52.927 |
|  | Latin America | 10 | 80.985 | 64.549 | 55.173 | 52.857 | 57.601 |
|  |  | 25 | 84.773 | 68.283 | 59.202 | 57.531 | 63.270 |
|  |  | 50 | 88.982 | 72.431 | 63.678 | 62.725 | 69.569 |
|  |  | 75 | 93.191 | 76.580 | 68.155 | 67.918 | 75.868 |
|  |  | 95 | 99.247 | 82.548 | 74.595 | 75.390 | 84.931 |
|  | Africa | 10 | **<LOD** | **<LOD** | **<LOD** | **<LOD** | **<LOD** |
|  |  | 25 | **<LOD** | **<LOD** | **<LOD** | **<LOD** | **<LOD** |
|  |  | 50 | 384.674 | 368.122 | 359.370 | 358.416 | 365.261 |
|  |  | 75 | 1017.584 | 1000.972 | 992.547 | 992.310 | 1000.261 |
|  |  | 95 | 1928.128 | 1911.429 | 1903.477 | 1904.271 | 1913.812 |
|  | Asia - EPA | 10 | 34.599 | 18.163 | 8.786 | 6.470 | 11.215 |
|  |  | 25 | 39.978 | 23.487 | 14.406 | 12.735 | 18.474 |
|  |  | 50 | 45.954 | 29.402 | 20.650 | 19.696 | 26.541 |
|  |  | 75 | 51.930 | 35.318 | 26.894 | 26.657 | 34.607 |
|  |  | 95 | 60.528 | 43.829 | 35.876 | 36.671 | 46.212 |
|  | Asia - MESA | 10 | 80.034 | 63.598 | 54.222 | 51.906 | 56.650 |
|  |  | 25 | 85.153 | 68.662 | 59.582 | 57.911 | 63.650 |
|  |  | 50 | 90.840 | 74.289 | 65.536 | 64.582 | 71.427 |
|  |  | 75 | 96.528 | 79.916 | 71.491 | 71.254 | 79.205 |
|  |  | 95 | 104.710 | 88.011 | 80.058 | 80.853 | 90.394 |
|  | Australia | 10 | 105.922 | 89.486 | 80.110 | 77.793 | 82.538 |
|  |  | 25 | 115.057 | 98.566 | 89.485 | 87.814 | 93.553 |
|  |  | 50 | 125.206 | 108.655 | 99.902 | 98.948 | 105.793 |
|  |  | 75 | 135.355 | 118.743 | 110.319 | 110.082 | 118.032 |
|  |  | 95 | 149.957 | 133.258 | 125.305 | 126.100 | 135.641 |
| MiBP (ng/mL) | Global | 10 | 22.749 | 21.661 | 20.709 | 19.896 | 19.219 |
|  |  | 25 | 26.320 | 27.386 | 28.319 | 29.120 | 29.788 |
|  |  | 50 | 30.287 | 33.747 | 36.774 | 39.369 | 41.530 |
|  |  | 75 | 34.254 | 40.108 | 45.229 | 49.617 | 53.272 |
|  |  | 95 | 39.961 | 49.260 | 57.393 | 64.362 | 70.166 |
|  | Latin America | 10 | 17.439 | 12.431 | 9.415 | 8.392 | 9.362 |
|  |  | 25 | 20.830 | 18.079 | 16.941 | 17.419 | 19.510 |
|  |  | 50 | 24.598 | 24.354 | 25.304 | 27.448 | 30.785 |
|  |  | 75 | 28.366 | 30.629 | 33.666 | 37.476 | 42.061 |
|  |  | 95 | 33.787 | 39.658 | 45.697 | 51.905 | 58.282 |
|  | Africa | 10 | 17.102 | 12.094 | 9.078 | 8.055 | 9.025 |
|  |  | 25 | 25.218 | 22.466 | 21.329 | 21.806 | 23.898 |
|  |  | 50 | 34.235 | 33.991 | 34.940 | 37.084 | 40.422 |
|  |  | 75 | 43.252 | 45.515 | 48.552 | 52.362 | 56.946 |
|  |  | 95 | 56.225 | 62.095 | 68.134 | 74.342 | 80.719 |
|  | Asia - EPA | 10 | 27.789 | 22.781 | 19.766 | 18.743 | 19.713 |
|  |  | 25 | 32.314 | 29.563 | 28.425 | 28.903 | 30.994 |
|  |  | 50 | 37.342 | 37.097 | 38.047 | 40.191 | 43.529 |
|  |  | 75 | 42.369 | 44.632 | 47.669 | 51.479 | 56.063 |
|  |  | 95 | 49.602 | 55.472 | 61.511 | 67.719 | 74.096 |
|  | Asia - MESA | 10 | 36.612 | 31.604 | 28.588 | 27.566 | 28.536 |
|  |  | 25 | 43.491 | 40.740 | 39.603 | 40.080 | 42.172 |
|  |  | 50 | 51.135 | 50.890 | 51.840 | 53.984 | 57.322 |
|  |  | 75 | 58.778 | 61.041 | 64.078 | 67.888 | 72.472 |
|  |  | 95 | 69.774 | 75.644 | 81.684 | 87.892 | 94.268 |
|  | Australia | 10 | 26.198 | 21.190 | 18.175 | 17.152 | 18.122 |
|  |  | 25 | 27.004 | 24.252 | 23.115 | 23.593 | 25.684 |
|  |  | 50 | 27.899 | 27.655 | 28.605 | 30.749 | 34.087 |
|  |  | 75 | 28.795 | 31.058 | 34.094 | 37.905 | 42.489 |
|  |  | 95 | 30.083 | 35.953 | 41.992 | 48.200 | 54.577 |
| MnBP (ng/mL) | Global | 10 | 46.07 | 24.94 | 30.72 | 63.40 | 123.00 |
|  |  | 25 | 50.63 | 30.09 | 36.77 | 70.64 | 131.73 |
|  |  | 50 | 55.69 | 35.82 | 43.49 | 78.69 | 141.43 |
|  |  | 75 | 60.76 | 41.55 | 50.21 | 86.74 | 151.13 |
|  |  | 95 | 68.05 | 49.79 | 59.88 | 98.31 | 165.08 |
|  | Latin America | 10 | 35.31 | 12.57 | 17.51 | 50.14 | 110.46 |
|  |  | 25 | 37.52 | 14.75 | 20.47 | 54.68 | 117.36 |
|  |  | 50 | 39.98 | 17.19 | 23.76 | 59.71 | 125.03 |
|  |  | 75 | 42.44 | 19.62 | 27.05 | 64.74 | 132.69 |
|  |  | 95 | 45.98 | 23.12 | 31.79 | 71.99 | 143.72 |
|  | Africa | 10 | **<LOD** | **<LOD** | **<LOD** | **<LOD** | 42.48 |
|  |  | 25 | 41.72 | 18.95 | 24.67 | 58.87 | 121.56 |
|  |  | 50 | 124.37 | 101.58 | 108.15 | 144.10 | 209.42 |
|  |  | 75 | 207.02 | 184.20 | 191.64 | 229.33 | 297.27 |
|  |  | 95 | 325.93 | 303.07 | 311.74 | 351.94 | 423.67 |
|  | Asia - EPA | 10 | 46.13 | 23.39 | 28.34 | 60.97 | 121.29 |
|  |  | 25 | 52.25 | 29.48 | 35.20 | 69.41 | 132.09 |
|  |  | 50 | 59.05 | 36.26 | 42.83 | 78.78 | 144.10 |
|  |  | 75 | 65.85 | 43.03 | 50.46 | 88.15 | 156.10 |
|  |  | 95 | 75.63 | 52.77 | 61.44 | 101.64 | 173.37 |
|  | Asia - MESA | 10 | 95.42 | 72.68 | 77.63 | 110.26 | 170.58 |
|  |  | 25 | 109.78 | 87.01 | 92.73 | 126.93 | 189.62 |
|  |  | 50 | 125.73 | 102.93 | 109.51 | 145.46 | 210.77 |
|  |  | 75 | 141.68 | 118.86 | 126.29 | 163.98 | 231.93 |
|  |  | 95 | 164.62 | 141.76 | 150.43 | 190.63 | 262.36 |
|  | Australia | 10 | 32.43 | 9.69 | 14.63 | 47.26 | 107.58 |
|  |  | 25 | 38.36 | 15.59 | 21.31 | 55.51 | 118.20 |
|  |  | 50 | 44.95 | 22.15 | 28.73 | 64.68 | 129.99 |
|  |  | 75 | 51.53 | 28.71 | 36.15 | 73.84 | 141.79 |
|  |  | 95 | 61.01 | 38.15 | 46.82 | 87.03 | 158.76 |
| MBzP (ng/mL) | Global | 10 | 5.297 | 3.258 | 2.035 | 1.629 | 2.040 |
|  |  | 25 | 5.837 | 3.786 | 2.480 | 1.921 | 2.108 |
|  |  | 50 | 6.437 | 4.372 | 2.975 | 2.245 | 2.184 |
|  |  | 75 | 7.036 | 4.958 | 3.470 | 2.570 | 2.259 |
|  |  | 95 | 7.899 | 5.802 | 4.181 | 3.036 | 2.368 |
|  | Latin America | 10 | 4.791 | 2.613 | 1.290 | 0.821 | **ND** |
|  |  | 25 | 5.287 | 3.054 | 1.628 | 1.008 | **ND** |
|  |  | 50 | 5.838 | 3.545 | 2.004 | 1.216 | **ND** |
|  |  | 75 | 6.388 | 4.035 | 2.380 | 1.423 | **ND** |
|  |  | 95 | 7.181 | 4.740 | 2.920 | 1.722 | **ND** |
|  | Africa | 10 | 0.280 | **<LOD** | **<LOD** | **<LOD** | **<LOD** |
|  |  | 25 | 2.753 | 0.521 | **<LOD** | **<LOD** | **<LOD** |
|  |  | 50 | 5.501 | 3.208 | 1.667 | 0.879 | 0.843 |
|  |  | 75 | 8.249 | 5.895 | 4.240 | 3.284 | 3.026 |
|  |  | 95 | 12.202 | 9.762 | 7.942 | 6.743 | 6.165 |
|  | Asia - EPA | 10 | 5.670 | 3.492 | 2.168 | 1.700 | 2.086 |
|  |  | 25 | 6.274 | 4.042 | 2.615 | 1.995 | 2.182 |
|  |  | 50 | 6.946 | 4.653 | 3.112 | 2.324 | 2.288 |
|  |  | 75 | 7.617 | 5.264 | 3.609 | 2.652 | 2.394 |
|  |  | 95 | 8.583 | 6.143 | 4.323 | 3.124 | 2.547 |
|  | Asia - MESA | 10 | 5.376 | 3.198 | 1.875 | 1.406 | 1.793 |
|  |  | 25 | 7.035 | 4.802 | 3.376 | 2.756 | 2.942 |
|  |  | 50 | 8.878 | 6.585 | 5.044 | 4.256 | 4.220 |
|  |  | 75 | 10.720 | 8.367 | 6.712 | 5.755 | 5.497 |
|  |  | 95 | 13.372 | 10.931 | 9.112 | 7.913 | 7.335 |
|  | Australia | 10 | 6.310 | 4.131 | 2.808 | 2.340 | 2.726 |
|  |  | 25 | 7.155 | 4.922 | 3.496 | 2.876 | 3.062 |
|  |  | 50 | 8.094 | 5.801 | 4.261 | 3.472 | 3.436 |
|  |  | 75 | 9.034 | 6.680 | 5.025 | 4.068 | 3.810 |
|  |  | 95 | 10.385 | 7.944 | 6.125 | 4.926 | 4.348 |

a. Predictive equations used to obtain the predicted values did not adjust for international regions.

b. Predictive equations used to obtain the predicted values were adjusted for international regions, pregnancy status, and age groups.

c. ND: Not detectable at the given time; <LOD: Value was below the limit of detection (pre-defined limit: 0.1 ng/mL).

Supplement Table 7: Meta-regressions between standard deviation values and year of sampling.

| **Phthalate Polymers** | **Predictor Variable** | **Meta-Regression Beta-Coefficient (95% CI) *** | **P-value** | **Meta-Regression with Quadric Term Beta-Coefficient (95% CI) **** | **P-value** |
| --- | --- | --- | --- | --- | --- |
| **High Molecular Weight** | | | | | |
| MEHP | Year | 0.012 (-0.12, 0.16) | 0.80 | 0.02 (-0.13, 0.18) | 0.76 |
|  | Year^2 | NA | NA | 0.002 (-0.02, 0.02) | 0.88 |
| MEHHP | Year | 0.09 (-0.15, 0.33) | 0.47 | 0.11 (-0.18, 0.40) | 0.45 |
|  | Year^2 | NA | NA | 0.01 (-0.03, 0.04) | 0.78 |
| MEOHP | Year | 0.06 (-0.14, 0.26) | 0.57 | 0.08 (-0.14, 0.29) | 0.48 |
|  | Year^2 | NA | NA | 0.01 (-0.02, 0.04) | 0.56 |
| MECPP | Year | 0.47 (-0.05, 0.99) | 0.08 | 0.35 (-0.25, 0.95) | 0.25 |
|  | Year^2 | NA | NA | -0.03 (-0.11, 0.05) | 0.44 |
| MCMHP | Year | -0.11 (-0.69, 0.47) | 0.70 | -0.19 (-0.89, 0.52) | 0.60 |
|  | Year^2 | NA | NA | 0.10 (-0.07, 0.28) | 0.24 |
| MCOP | Year | 0.08 (-0.10, 0.25) | 0.37 | 0.08 (-0.10, 0.26) | 0.36 |
|  | Year^2 | NA | NA | -0.005 (-0.02, 0.01) | 0.62 |
| MiNP | Year | 0.003 (-0.04, 0.04) | 0.89 | 0.02 (-0.04, 0.08) | 0.45 |
|  | Year^2 | NA | NA | 0.003 (-0.004, 0.01) | 0.38 |
| MOiNP | Year | 0.03 (-0.24, 0.31) | 0.74 | 0.25 (-0.46, 0.96) | 0.35 |
|  | Year^2 | NA | NA | 0.03 (-0.07, 0.12) | 0.37 |
| OH-MiNP | Year | 0.06 (-0.17, 0.29) | 0.55 | 0.44 (-0.27, 1.15) | 0.17 |
|  | Year^2 | NA | NA | 0.04 (-0.03, 0.12) | 0.21 |
| MCiNP | Year | 0.06 (0.03, 0.09) | 0.00 | 0.10 (0.04, 0.15) | 0.00 |
|  | Year^2 | NA | NA | 0.01 (-0.003, 0.02) | 0.12 |
| MOP | Year | 0.01 (-0.06, 0.07) | 0.65 | -0.04 (-0.13, 0.04) | 0.29 |
|  | Year^2 | NA | NA | -0.01 (-0.03, 0.002) | 0.09 |
|  |  | a. |  |  |  |
| MCPP | Year | 0.03 (-0.07, 0.12) | 0.58 | 0.02 (-0.09, 0.13) | 0.67 |
|  | Year^2 | NA | NA | -0.001 (-0.01, 0.01) | 0.94 |
| MiDP | Year | 0.64 (-1.00, 2.28) | 0.24 | 1.06 (-2.58, 4.70) | 0.17 |
|  | Year^2 | NA | NA | 0.22 (-0.41, 0.86) | 0.14 |
|  |  | a. |  | a. |  |
| **Low Molecular Weight** | | | | | |
| MEP | Year | 0.14 (-0.26, 0.53) | 0.50 | 0.18 (-0.28, 0.64) | 0.45 |
|  | Year^2 | NA | NA | 0.01 (-0.06, 0.08) | 0.74 |
| MiBP | Year | 0.50 (-0.02, 1.02) | 0.06 | 0.53 (-0.15, 1.21) | 0.13 |
|  | Year^2 | NA | NA | -0.01 (-0.11, 0.08) | 0.80 |
| MnBP | Year | 0.28 (-0.27, 0.84) | 0.32 | 0.41 (-0.29, 1.10) | 0.25 |
|  | Year^2 | NA*** | NA | 0.03 (-0.08, 0.13) | 0.62 |
| MBzP | Year | -0.04 (-0.09, 0.01) | 0.09 | -0.04 (-0.09, 0.01) | 0.08 |
|  | Year^2 | NA | NA | -0.001 (-0.01, 0.01) | 0.68 |

*Weighted for each study's standard error; adjusted for geographical location, age group, and pregnancy status

**Weighted for each study's standard error; adjusted for geographical location, age group, pregnancy status, and quadratic term for time

*** NA = Not Available

a. Pregnancy Status was omitted during the regression due to collinearity

b. Analysis was unfeasible due to limited or unavailable observations in the region

Supplement Table 8: Sensitivity Meta-Analysis between phthalates and year of sampling internationally and stratified for geographical regions without Geometric Mean Articles

| **Phthalate Polymers** | **Regions** | **Predictor Variable** | **Meta-Regression β-Coefficient (95% CI) *** | **P-value** | **Meta-Regression with Quadric Term β-Coefficient (95% CI) **** | **P-value** |
| --- | --- | --- | --- | --- | --- | --- |
| **High Molecular Weight** | | | | | | |
| MEHP | International | Year | -0.26 (-0.82, 0.29) | 0.3 | -0.18 (-0.78, 0.42) | 0.56 |
|  |  | Year^2 | NA | NA | 0.02 (-0.07, 0.11) | 0.62 |
|  | Latin America | Year | -2.87 (-58.45, 52.72) | 0.63 | -0.46 (-46.66, 45.73) | 0.919 |
|  |  | Year^2 | NA | NA | -0.33 (-14.71,14.04) | 0.818 |
|  |  |  | b. |  | d. |  |
|  | Africa | Year | 1.02 (-3.91, 5.95) | 0.23 | 1.02 (-3.91, 5.95) | 0.23 |
|  |  | Year^2 | NA | NA | NA | NA |
|  |  |  | d. |  | d. |  |
|  | Asia - EPA | Year | -0.22 (-0.64, 0.21) | 0.31 | -0.10 (-0.56, 0.36) | 0.68 |
|  |  | Year^2 | NA | NA | 0.03 (-0.03, 0.10) | 0.31 |
|  | Asia - MESA | Year | -8.63 (-461.55, 444.28) | 0.85 | 6.97 (-673.69, 687.63) | 0.92 |
|  |  | Year^2 | NA | NA | -12.9 (-187.23, 161.29) | 0.52 |
|  |  |  | a. |  | b. |  |
|  | Australia | Year | NA | NA | NA | NA |
|  |  | Year^2 | NA | NA | NA | NA |
|  |  |  | c. |  | c. |  |
| MEHHP | International | Year | -0.38 (-1.80, 1.04) | 0.6 | -0.14 (-1.72, 1.45) | 0.86 |
|  |  | Year^2 | NA | NA | 0.09 (-0.15, 0.33) | 0.46 |
|  | Latin America | Year | -1.69 (-128.43, 125.06) | 0.89 | NA | NA |
|  |  | Year^2 | NA | NA | NA | NA |
|  |  |  | d. |  | c. |  |
|  | Africa | Year | NA | NA | NA | NA |
|  |  | Year^2 | NA | NA | NA | NA |
|  |  |  | c. |  | c. |  |
|  | Asia - EPA | Year | -0.39 (-1.12, 0.33) | 0.28 | -0.15 (-0.94, 0.64) | 0.71 |
|  |  | Year^2 | NA | NA | 0.10 (-0.02, 0.21) | 0.11 |
|  | Asia - MESA | Year | -83.54 (-4301.64, 4134.57) | 0.84 | 346.01 (-776.79, 1468.80) | 0.16 |
|  |  | Year^2 | NA | NA | -117.95 (-412.12, 176.23) | 0.12 |
|  |  |  | a. |  | b. |  |
|  | Australia | Year | NA | NA | NA | NA |
|  |  | Year^2 | NA | NA | NA | NA |
|  |  |  | c. |  | c. |  |
| MEOHP | International | Year | -0.91 (-1.73, -0.10) | 0.03 | -0.76 (-1.59, 0.07) | 0.07 |
|  |  | Year^2 | NA | NA | 0.05 (-0.08, 0.18) | 0.45 |
|  | Latin America | Year | -2.10 (-82.52, 78.31) | 0.8 | NA | NA |
|  |  | Year^2 | NA | NA | NA | NA |
|  |  |  | d. |  | c. |  |
|  | Africa | Year | NA | NA | NA | NA |
|  |  | Year^2 | NA | NA | NA | NA |
|  |  |  | c. |  | c. |  |
|  | Asia - EPA | Year | -0.74 (-1.37, -0.10) | 0.02 | -0.57 (-1.23, 0.09) | 0.09 |
|  |  | Year^2 | NA | NA | 0.06 (-0.04, 0.16) | 0.22 |
|  | Asia - MESA | Year | 17.42 (-979.47, 1014.31) | 0.86 | 94.93 (-1514.69, 1704.55) | 0.59 |
|  |  | Year^2 | NA | NA | -21.16 (-316.63, 274.31) | 0.53 |
|  |  |  | b. |  | d. |  |
|  | Australia | Year | NA | NA | NA | NA |
|  |  | Year^2 | NA | NA | NA | NA |
|  |  |  | c. |  | c. |  |
| MECPP | International | Year | -0.37 (-2.60, 1.86) | 0.74 | -0.47 (-2.73, 1.12) | 0.68 |
|  |  | Year^2 | NA | NA | 0.25 (-0.32, 0.82) | 0.39 |
|  | Latin America | Year | -3.89 (-247.25, 239.47) | 0.872 | NA | NA |
|  |  | Year^2 | NA | NA | NA | NA |
|  |  |  | d. |  | c. |  |
|  | Africa | Year | NA | NA | NA | NA |
|  |  | Year^2 | NA | NA | NA | NA |
|  |  |  | c. |  | c. |  |
|  | Asia - EPA | Year | 0.06 (-2.15, 2.27) | 0.96 | -0.08 (-2.34, 2.18) | 0.94 |
|  |  | Year^2 | NA | NA | 0.20 (-0.38, 0.78) | 0.49 |
|  | Asia - MESA | Year | -6.87 (-197.18, 183.44) | 0.73 | NA | NA |
|  |  | Year^2 | NA | NA | NA | NA |
|  |  |  | d. |  | c. |  |
|  | Australia | Year | NA | NA | NA | NA |
|  |  | Year^2 | NA | NA | NA | NA |
|  |  |  | c. |  | c. |  |
| MCMHP | International | Year | 0.89 (-0.36, 2.14) | 0.15 | 0.56 (-0.76, 1.88) | 0.38 |
|  |  | Year^2 | NA | NA | 0.16 (-0.08, 0.41) | 0.18 |
|  | Latin America | Year | NA | NA | NA | NA |
|  |  | Year^2 | NA | NA | NA | NA |
|  |  |  | c. |  | c. |  |
|  | Africa | Year | NA | NA | NA | NA |
|  |  | Year^2 | NA | NA | NA | NA |
|  |  |  | c. |  | c. |  |
|  | Asia - EPA | Year | 0.94 (-0.29, 2.17) | 0.12 | 0.63 (-0.66, 1.93) | 0.32 |
|  |  | Year^2 | NA | NA | 0.16 (-0.09, 0.40) | 0.19 |
|  | Asia - MESA | Year | NA | NA | NA | NA |
|  |  | Year^2 | NA | NA | NA | NA |
|  |  |  | c. |  | c. |  |
|  | Australia | Year | NA | NA | NA | NA |
|  |  | Year^2 | NA | NA | NA | NA |
|  |  |  | c. |  | c. |  |
| MCOP | International | Year | 0.36 (-0.25, 0.97) | 0.18 | 1.00 (-0.76, 2.75) | 0.17 |
|  |  | Year^2 | NA | NA | 0.05 (-0.08, 0.19) | 0.3 |
|  | Latin America | Year | 0.26 (-10.84, 11.37) | 0.81 | -0.99 (-12.82, 10.84) | 0.48 |
|  |  | Year^2 | NA | NA | -0.12 (-1.3, 1.08) | 0.41 |
|  |  |  | b. |  | d. |  |
|  | Africa | Year | NA | NA | NA | NA |
|  |  | Year^2 | NA | NA | NA | NA |
|  |  |  | c. |  | c. |  |
|  | Asia - EPA | Year | -0.02 (-5.02, 5.00) | 0.98 | -1.48 (-15.11, 12. 14) | 0.40 |
|  |  | Year^2 | NA | NA | 0.15 (-1.29, 1.60) | 0.40 |
|  |  |  | a. |  | c. |  |
|  | Asia - MESA | Year | NA | NA | NA | NA |
|  |  | Year^2 | NA | NA | NA | NA |
|  |  |  | c. |  | c. |  |
|  | Australia | Year | NA | NA | NA | NA |
|  |  | Year^2 | NA | NA | NA | NA |
|  |  |  | c. |  | c. |  |
| MiNP | International | Year | 0.11 (-0.26, 0.47) | 0.54 | 0.23 (-0.32, 0.78) | 0.38 |
|  |  | Year^2 | NA | NA | 0.02 (-0.06, 0.10) | 0.51 |
|  | Latin America | Year | NA | NA | NA | NA |
|  |  | Year^2 | NA | NA | NA | NA |
|  |  |  | c. |  | c. |  |
|  | Africa | Year | NA | NA | NA | NA |
|  |  | Year^2 | NA | NA | NA | NA |
|  |  |  | c. |  | c. |  |
|  | Asia - EPA | Year | 0.11 (-0.26, 0.47) | 0.54 | 0.23 (-0.32, 0.78) | 0.38 |
|  |  | Year^2 | NA | NA | 0.02 (-0.06, 0.10) | 0.51 |
|  | Asia - MESA | Year | NA | NA | NA | NA |
|  |  | Year^2 | NA | NA | NA | NA |
|  |  |  | c. |  | c. |  |
|  | Australia | Year | NA | NA | NA | NA |
|  |  | Year^2 | NA | NA | NA | NA |
|  |  |  | c. |  | c. |  |
| MOiNP | International | Year | -1.77 (-2.25, -1.28) | 0.01 | -1.77 (-2.25, -1.28) | 0.01 |
|  |  | Year^2 | NA | NA | NA | NA |
|  |  |  | d. |  | f. |  |
|  | Latin America | Year | NA | NA | NA | NA |
|  |  | Year^2 | NA | NA | NA | NA |
|  |  |  | c. |  | c. |  |
|  | Africa | Year | NA | NA | NA | NA |
|  |  | Year^2 | NA | NA | NA | NA |
|  |  |  | c. |  | c. |  |
|  | Asia - EPA | Year | -1.77 (-2.25, -1.28) | 0.01 | -1.77 (-2.25, -1.28) | 0.01 |
|  |  | Year^2 | NA | NA | NA | NA |
|  |  |  | d. |  | f. |  |
|  | Asia - MESA | Year | NA | NA | NA | NA |
|  |  | Year^2 | NA | NA | NA | NA |
|  |  |  | c. |  | c. |  |
|  | Australia | Year | NA | NA | NA | NA |
|  |  | Year^2 | NA | NA | NA | NA |
|  |  |  | c. |  | c. |  |
| OH-MiNP | International | Year | -1.10 (-1.89, -0.30) | 0.04 | -1.10 (-1.89, -0.30) | 0.04 |
|  |  | Year^2 | NA | NA | NA | NA |
|  |  |  | d. |  | f. |  |
|  | Latin America | Year | NA | NA | NA | NA |
|  |  | Year^2 | NA | NA | NA | NA |
|  |  |  | c. |  | c. |  |
|  | Africa | Year | NA | NA | NA | NA |
|  |  | Year^2 | NA | NA | NA | NA |
|  |  |  | c. |  | c. |  |
|  | Asia - EPA | Year | -1.10 (-1.89, -0.30) | 0.04 | -1.10 (-1.89, -0.30) | 0.04 |
|  |  | Year^2 | NA | NA | NA | NA |
|  |  |  | d. |  | f. |  |
|  | Asia - MESA | Year | NA | NA | NA | NA |
|  |  | Year^2 | NA | NA | NA | NA |
|  |  |  | c. |  | c. |  |
|  | Australia | Year | NA | NA | NA | NA |
|  |  | Year^2 | NA | NA | NA | NA |
|  |  |  | c. |  | c. |  |
| MCiNP | International | Year | 0.01 (-0.31, 0.33) | 0.82 | -0.10 (-0.32, 0.13) | 0.12 |
|  |  | Year^2 | NA | NA | 0.29 (-1.87, 2.46) | 0.18 |
|  |  |  | a. |  | d. |  |
|  | Latin America | Year | -0.05 (-0.59, 0.48) | 0.435 | NA | NA |
|  |  | Year^2 | NA | NA | NA | NA |
|  |  |  | d. |  | c. |  |
|  | Africa | Year | NA | NA | NA | NA |
|  |  | Year^2 | NA | NA | NA | NA |
|  |  |  | c. |  | c. |  |
|  | Asia - EPA | Year | NA | NA | NA | NA |
|  |  | Year^2 | NA | NA | NA | NA |
|  |  |  | c. |  | c. |  |
|  | Asia - MESA | Year | NA | NA | NA | NA |
|  |  | Year^2 | NA | NA | NA | NA |
|  |  |  | c. |  | c. |  |
|  | Australia | Year | NA | NA | NA | NA |
|  |  | Year^2 | NA | NA | NA | NA |
|  |  |  | c. |  | c. |  |
| MOP | International | Year | 0.01 (-0.20, 0.22) | 0.92 | -0.002 (-0.89, 0.89) | 1 |
|  |  | Year^2 | NA | NA | -0.002 (-0.17, 0.16) | 0.98 |
|  | Latin America | Year | NA | NA | NA | NA |
|  |  | Year^2 | NA | NA | NA | NA |
|  |  |  | c. |  | c. |  |
|  | Africa | Year | NA | NA | NA | NA |
|  |  | Year^2 | NA | NA | NA | NA |
|  |  |  | c. |  | c. |  |
|  | Asia - EPA | Year | 0.01 (-0.04, 0.06) | 0.58 | 0.06 (-0.14, 0.26) | 0.47 |
|  |  | Year^2 | NA | NA | 0.01 (-0.03, 0.05) | 0.54 |
|  | Asia - MESA | Year | NA | NA | NA | NA |
|  |  | Year^2 | NA | NA | NA | NA |
|  |  |  | c. |  | c. |  |
|  | Australia | Year | NA | NA | NA | NA |
|  |  | Year^2 | NA | NA | NA | NA |
|  |  |  | c. |  | c. |  |
| MCPP | International | Year | -0.16 (-0.37, 0.04) | 0.1 | -0.18 (-0.40, 0.04) | 0.1 |
|  |  | Year^2 | NA | NA | -0.03 (-0.11, 0.05) | 0.39 |
|  | Latin America | Year | -0.20 (-5.77, 5.37) | 0.73 | NA | NA |
|  |  | Year^2 | NA | NA | NA | NA |
|  |  |  | d. |  | c. |  |
|  | Africa | Year | NA | NA | NA | NA |
|  |  | Year^2 | NA | NA | NA | NA |
|  |  |  | c. |  | c. |  |
|  | Asia - EPA | Year | -0.18 (-0.45, 0.09) | 0.15 | 0.41 (-0.38, 1.19) | 0.24 |
|  |  | Year^2 | NA | NA | -0.64 (-1.45, 0.18) | 0.1 |
|  | Asia - MESA | Year | NA | NA | NA | NA |
|  |  | Year^2 | NA | NA | NA | NA |
|  |  |  | c. |  | c. |  |
|  | Australia | Year | NA | NA | NA | NA |
|  |  | Year^2 | NA | NA | NA | NA |
|  |  |  | c. |  | c. |  |
| MiDP | International | Year | 1.41 (1.00, 1.81) | 0.01 | NA | NA |
|  |  | Year^2 | NA | NA | NA | NA |
|  |  |  | f. |  | c. |  |
|  | Latin America | Year | NA | NA | NA | NA |
|  |  | Year^2 | NA | NA | NA | NA |
|  |  |  | c. |  | c. |  |
|  | Africa | Year | NA | NA | NA | NA |
|  |  | Year^2 | NA | NA | NA | NA |
|  |  |  | c. |  | c. |  |
|  | Asia - EPA | Year | NA | NA | NA | NA |
|  |  | Year^2 | NA | NA | NA | NA |
|  |  |  | c. |  | c. |  |
|  | Asia - MESA | Year | NA | NA | NA | NA |
|  |  | Year^2 | NA | NA | NA | NA |
|  |  |  | c. |  | c. |  |
|  | Australia | Year | NA | NA | NA | NA |
|  |  | Year^2 | NA | NA | NA | NA |
|  |  |  | c. |  | c. |  |
| **Low Molecular Weight** | | | | | | |
| MEP | International | Year | -0.65 (-2.77, 1.47) | 0.54 | -0.27 (-2.63, 2.10) | 0.82 |
|  |  | Year^2 | NA | NA | 0.14 (-0.20, 0.49) | 0.43 |
|  | Latin America | Year | -17.58 (-682.69, 647.53) | 0.79 | NA | NA |
|  |  | Year^2 | NA | NA | NA | NA |
|  |  |  | d. |  | c. |  |
|  | Africa | Year | -8.98 (-9.70, -8.26) | <0.01 | -8.98 (-9.70, -8.26) | <0.01 |
|  |  | Year^2 | NA | NA | NA | NA |
|  |  |  | d. |  | f. |  |
|  | Asia - EPA | Year | -0.71 (-2.5548, 1.13) | 0.44 | -0.26 (-2.38, 1.69) | 0.74 |
|  |  | Year^2 | NA | NA | 0.12 (-0.18, 0.41) | 0.43 |
|  | Asia - MESA | Year | 95.53 (51.29, 139.76) | 0.02 | NA | NA |
|  |  | Year^2 | NA | NA | NA | NA |
|  |  |  | d. |  | c. |  |
|  | Australia | Year | NA | NA | NA | NA |
|  |  | Year^2 | NA | NA | NA | NA |
|  |  |  | c. |  | c. |  |
| MiBP | International | Year | 0.56 (-1.01, 2.13) | 0.48 | 0.61 (-1.05, 2.26) | 0.47 |
|  |  | Year^2 | NA | NA | 0.03 (-0.35, 0.40) | 0.89 |
|  | Latin America | Year | -0.11 (-13.87, 13.65) | 0.94 | -16.35 (-40.43, 7.73) | 0.07 |
|  |  | Year^2 | NA | NA | 4.63 (-5.21, 14.47) | 0.11 |
|  |  |  | a. |  | b. |  |
|  | Africa | Year | 0.86 (-23.36, 25.08) | 0.73 | NA | NA |
|  |  | Year^2 | NA | NA | -0.73 (-21.29, 19.83) | 0.73 |
|  |  |  | d. |  | e. |  |
|  | Asia - EPA | Year | 0.42 (-1.19, 2.04) | 0.6 | 0.38 (-1.34, 2.10) | 0.66 |
|  |  | Year^2 | NA | NA | -0.04 (-0.41, 0.33) | 0.84 |
|  | Asia - MESA | Year | 8.69 (-589.28, 606.65) | 0.88 | NA | NA |
|  |  | Year^2 | NA | NA | NA | NA |
|  |  |  | d. |  | c. |  |
|  | Australia | Year | NA | NA | NA | NA |
|  |  | Year^2 | NA | NA | NA | NA |
|  |  |  | c. |  | c. |  |
| MnBP | International | Year | 6.45 (3.35, 9.56) | <0.01 | 9.19 (5.81, 12.58) | <0.01 |
|  |  | Year^2 | NA*** | NA | 0.93 (0.39, 1.47) | <0.01 |
|  | Latin America | Year | -2.35 (-49.64. 44.95) | 0.64 | -24.49 (-368.27, 319.29) | 0.53 |
|  |  | Year^2 | NA | NA | 5.95 (-85.08, 96.98) | 0.56 |
|  |  |  | a. |  | b. |  |
|  | Africa | Year | 13.75 (-0.61, 28.10) | 0.05 | 13.75 (-0.61, 28.10) | 0.05 |
|  |  | Year^2 | NA | NA | NA | NA |
|  |  |  | d. |  | f. |  |
|  | Asia - EPA | Year | 6.34 (3.12, 9.57) | <0.01 | 9.50 (5.91, 13.09) | <0.01 |
|  |  | Year^2 | NA | NA | 0.96 (0.41, 1.52) | <0.01 |
|  | Asia - MESA | Year | 51.55 (-1053.64, 1156.73) | 0.66 | 168.12 (147.79, 188.45) | 0.01 |
|  |  | Year^2 | NA | NA | -31.16 (-36.57, -25.76) | 0.01 |
|  |  |  | a. |  | b. |  |
|  | Australia | Year | NA | NA | NA | NA |
|  |  | Year^2 | NA | NA | NA | NA |
|  |  |  | c. |  | c. |  |
| MBzP | International | Year | -0.36 (-0.58, -0.15) | <0.01 | -0.25 (-0.48, -0.03) | 0.03 |
|  |  | Year^2 | NA | NA | 0.04 ( 0.01, 0.08) | 0.02 |
|  | Latin America | Year | -0.28 (-4.30, 3.74) | 0.54 | -1.30 (-7.74, 5.13) | 0.24 |
|  |  | Year^2 | NA | NA | 0.32 (-1.37, 2.01) | 0.25 |
|  |  |  | a. |  | b. |  |
|  | Africa | Year | NA | NA | NA | NA |
|  |  | Year^2 | NA | NA | NA | NA |
|  |  |  | c. |  | c. |  |
|  | Asia - EPA | Year | -0.37 (-0.58, -0.16) | <0.01 | -0.25 (-0.47, -0.03) | 0.03 |
|  |  | Year^2 | NA | NA | 0.04 (0.01, 0.08) | 0.02 |
|  | Asia - MESA | Year | 49.86 (-2025.92, 2125.77) | 0.81 | 236.96 (-1385.25, 1859.18) | 0.32 |
|  |  | Year^2 | NA | NA | -59.13 (-4112.64, 294.37) | 0.28 |
|  |  |  | a. |  | b. |  |
|  | Australia | Year | NA | NA | NA | NA |
|  |  | Year^2 | NA | NA | NA | NA |
|  |  |  | c. |  | c. |  |

*Weighted for each study's standard error; adjusted for geographical location, age group, and pregnancy status

**Weighted for each study's standard error; adjusted for geographical location, age group, pregnancy status, and quadratic term for time

*** NA = Not Available

a. Pregnancy status was omitted during the regression due to collinearity

b. Age group was omitted during the regression due to collinearity

c. Analysis was unfeasible due to limited or unavailable observations in the region

d. Age group and pregnancy was omitted during the regression due to collinearity

e. All variables besides the quadratic variable for time is omitted due to collinearity

f. All variables besides the time year of sampling is omitted due to collinearity

Supplement Table 9: Sensitivity Meta-Analysis between phthalates and year of sampling internationally and stratified for geographical regions without Arithmetic Mean Articles

| **Phthalate Polymers** | **Regions** | **Predictor Variable** | **Meta-Regression β-Coefficient (95% CI) *** | **P-value** | **Meta-Regression with Quadric Term β-Coefficient (95% CI) **** | **P-value** |
| --- | --- | --- | --- | --- | --- | --- |
| **High Molecular Weight** | | | | | | |
| MEHP | International | Year | -0.46 (-0.70, -0.22) | <0.01 | -0.41 (-0.68, -0.13) | <0.01 |
|  |  | Year^2 | NA | NA | 0.02 (-0.02, 0.06) | 0.41 |
|  | Latin America | Year | -0.04 (-0.45, 0.38) | 0.85 | -0.12 (-0.58, 0.35) | 0.57 |
|  |  | Year^2 | NA | NA | -0.01 (-0.08, 0.06) | 0.78 |
|  | Africa | Year | NA | NA | NA | NA |
|  |  | Year^2 | NA | NA | NA | NA |
|  |  |  | c. |  | c. |  |
|  | Asia - EPA | Year | -0.60 (-0.87, -0.32) | <0.01 | -0.54 (-0.84, -0.23) | <0.01 |
|  |  | Year^2 | NA | NA | 0.02 (-0.02, 0.07) | 0.32 |
|  | Asia - MESA | Year | -0.06 (-6.97, 6.85) | 0.93 | NA | NA |
|  |  | Year^2 | NA | NA | NA | NA |
|  |  |  | d. |  | c. |  |
|  | Australia | Year | NA | NA | NA | NA |
|  |  | Year^2 | NA | NA | NA | NA |
|  |  |  | c. |  | c. |  |
| MEHHP | International | Year | -0.93 (-1.44, -0.43) | <0.01 | -0.59 (-1.17, -0.001) | 0.05 |
|  |  | Year^2 | NA | NA | 0.09 (0.01, 0.17) | 0.02 |
|  | Latin America | Year | -0.82 (-2.70, 1.06) | 0.31 | -1.06 (-4.59, 2.48) | 0.45 |
|  |  | Year^2 | NA | NA | -0.04 (-0.51, 0.43) | 0.82 |
|  | Africa | Year | NA | NA | NA | NA |
|  |  | Year^2 | NA | NA | NA | NA |
|  |  |  | c. |  | c. |  |
|  | Asia - EPA | Year | -0.97 (-1.53, -0.41) | <0.01 | -0.59 (-1.21, 0.03) | 0.06 |
|  |  | Year^2 | NA | NA | 0.12 (0.03, 0.21) | 0.01 |
|  | Asia - MESA | Year | -0.73 (-6.67, 5.21) | 0.36 | NA | NA |
|  |  | Year^2 | NA | NA | NA | NA |
|  |  |  | d. |  | c. |  |
|  | Australia | Year | NA | NA | NA | NA |
|  |  | Year^2 | NA | NA | NA | NA |
|  |  |  | c. |  | c. |  |
| MEOHP | International | Year | -1.01 (-1.40, -0.62) | <0.01 | -1.02 (-1.43, -0.60) | <0.01 |
|  |  | Year^2 | NA | NA | -0.003 (-0.07, 0.06) | 0.92 |
|  | Latin America | Year | -0.51 (-1.82, 0.81) | 0.37 | -0.97 (-3.19, 1.25) | 0.29 |
|  |  | Year^2 | NA | NA | -0.09 (-0.39, 0.22) | 0.48 |
|  | Africa | Year | NA | NA | NA | NA |
|  |  | Year^2 | NA | NA | NA | NA |
|  |  |  | c. |  | c. |  |
|  | Asia - EPA | Year | -1.11 (-1.53, -0.69) | <0.01 | -1.08 (-1.53, -0.64) | <0.01 |
|  |  | Year^2 | NA | NA | 0.02 (-0.06, 0.09) | 0.68 |
|  | Asia - MESA | Year | -0.77 (-7.29, 5.75) | 0.38 | NA | NA |
|  |  | Year^2 | NA | NA | NA | NA |
|  |  |  | d. |  | c. |  |
|  | Australia | Year | NA | NA | NA | NA |
|  |  | Year^2 | NA | NA | NA | NA |
|  |  |  | c. |  | c. |  |
| MECPP | International | Year | -1.17 (-2.19, -0.15) | 0.03 | -0.90 (-2.13, 0.33) | 0.15 |
|  |  | Year^2 | NA | NA | 0.07 (-0.11, 0.24) | 0.45 |
|  | Latin America | Year | -1.02 (-5.76, 3.72) | 0.54 | -1.94 (-15.32, 11.43) | 0.6 |
|  |  | Year^2 | NA | NA | -0.13 (-1.67, 1.41) | 0.75 |
|  | Africa | Year | NA | NA | NA | NA |
|  |  | Year^2 | NA | NA | NA | NA |
|  |  |  | c. |  | c. |  |
|  | Asia - EPA | Year | -1.37 (-2.57, -0.16) | 0.03 | -0.90 (-2.20, 0.39) | 0.17 |
|  |  | Year^2 | NA | NA | 0.21 (-0.02, 0.43) | 0.07 |
|  | Asia - MESA | Year | -1.71 (-14.16, 10.74) | 0.33 | NA | NA |
|  |  | Year^2 | NA | NA | NA | NA |
|  |  |  |  |  | c. |  |
|  | Australia | Year | NA | NA | NA | NA |
|  |  | Year^2 | NA | NA | NA | NA |
|  |  |  | c. |  | c. |  |
| MCMHP | International | Year | -0.70 (-1.39, -0.01) | 0.05 | -0.67 (-1.43, 0.10) | 0.08 |
|  |  | Year^2 | NA | NA | 0.03 (-0.22, 0.27) | 0.81 |
|  | Latin America | Year | NA | NA | NA | NA |
|  |  | Year^2 | NA | NA | NA | NA |
|  |  |  | c. |  | c. |  |
|  | Africa | Year | NA | NA | NA | NA |
|  |  | Year^2 | NA | NA | NA | NA |
|  |  |  | c. |  | c. |  |
|  | Asia - EPA | Year | -0.70 (-1.41, 0.01) | 0.05 | -0.67 (-1.45, 0.11) | 0.09 |
|  |  | Year^2 | NA | NA | 0.03 (-0.22, 0.28) | 0.82 |
|  | Asia - MESA | Year | NA | NA | NA | NA |
|  |  | Year^2 | NA | NA | NA | NA |
|  |  |  | c. |  | c. |  |
|  | Australia | Year | NA | NA | NA | NA |
|  |  | Year^2 | NA | NA | NA | NA |
|  |  |  | c. |  | c. |  |
| MCOP | International | Year | -0.19 (-0.91, 0.53) | 0.57 | -0.87 (-1.94, 0.19) | 0.1 |
|  |  | Year^2 | NA | NA | -0.08 (-0.17, 0.02) | 0.1 |
|  | Latin America | Year | -0.21 (-2.49, 2.08) | 0.74 | -1.31 (-5.58, 2.96) | 0.16 |
|  |  | Year^2 | NA | NA | -0.11 (-0.47, 0.25) | 0.16 |
|  |  |  | a. |  |  |  |
|  | Africa | Year | NA | NA | NA | NA |
|  |  | Year^2 | NA | NA | NA | NA |
|  |  |  | c. |  | c. |  |
|  | Asia - EPA | Year | -0.34 (-2.73, 2.05) | 0.74 | -14.89 (-21.58, -8.19) | <0.01 |
|  |  | Year^2 | NA | NA | 2.13 (1.16, 3.10) | <0.01 |
|  | Asia - MESA | Year | NA | NA | NA | NA |
|  |  | Year^2 | NA | NA | NA | NA |
|  |  |  | c. |  | c. |  |
|  | Australia | Year | NA | NA | NA | NA |
|  |  | Year^2 | NA | NA | NA | NA |
|  |  |  | c. |  | c. |  |
| MiNP | International | Year | -0.05 (-0.24, 0.13) | 0.55 | 0.09 (-0.10, 0.28) | 0.32 |
|  |  | Year^2 | NA | NA | 0.03 (0.01, 0.06) | 0.02 |
|  | Latin America | Year | -0.04 (-0.13, 0.05) | 0.11 | -0.03 (-0.09, 0.03) | 0.1 |
|  |  | Year^2 | NA | NA | 0.004 (-0.01, 0.02) | 0.24 |
|  |  |  | a. |  | d. |  |
|  | Africa | Year | NA | NA | NA | NA |
|  |  | Year^2 | NA | NA | NA | NA |
|  |  |  | c. |  | c. |  |
|  | Asia - EPA | Year | -0.08 (-0.29, 0.14) | 0.45 | 0.15 (-0.13, 0.44) | 0.27 |
|  |  | Year^2 | NA | NA | 0.04 (0.002, 0.08) | 0.04 |
|  | Asia - MESA | Year | NA | NA | NA | NA |
|  |  | Year^2 | NA | NA | NA | NA |
|  |  |  | c. |  | c. |  |
|  | Australia | Year | NA | NA | NA | NA |
|  |  | Year^2 | NA | NA | NA | NA |
|  |  |  | c. |  | c. |  |
| MOiNP | International | Year | -0.15 (-0.46, 0.16) | 0.22 | -0.36 (-1.32, 0.61) | 0.25 |
|  |  | Year^2 | NA | NA | -0.03 (-0.17, 0.10) | 0.41 |
|  | Latin America | Year | NA | NA | NA | NA |
|  |  | Year^2 | NA | NA | NA | NA |
|  |  |  | c. |  | c. |  |
|  | Africa | Year | NA | NA | NA | NA |
|  |  | Year^2 | NA | NA | NA | NA |
|  |  |  | c. |  | c. |  |
|  | Asia - EPA | Year | -0.14 (-0.60, 0.33) | 0.33 | -0.39 (-3.20, 2.42) | 0.33 |
|  |  | Year^2 | NA | NA | -0.04 (-0.44, 0.36) | 0.43 |
|  | Asia - MESA | Year | NA | NA | NA | NA |
|  |  | Year^2 | NA | NA | NA | NA |
|  |  |  | c. |  | c. |  |
|  | Australia | Year | NA | NA | NA | NA |
|  |  | Year^2 | NA | NA | NA | NA |
|  |  |  | c. |  | c. |  |
| OH-MiNP | International | Year | -0.13 (-0.39, 0.13) | 0.27 | -0.67 (-1.24, -0.09) | 0.03 |
|  |  | Year^2 | NA | NA | -0.06 (-0.13, 0.001) | 0.05 |
|  | Latin America | Year | NA | NA | NA | NA |
|  |  | Year^2 | NA | NA | NA | NA |
|  |  |  | c. |  | c. |  |
|  | Africa | Year | NA | NA | NA | NA |
|  |  | Year^2 | NA | NA | NA | NA |
|  |  |  | c. |  | c. |  |
|  | Asia - EPA | Year | -0.13 (-0.44, 0.18) | 0.32 | -0.68 (-1.39, 0.04) | 0.06 |
|  |  | Year^2 | NA | NA | -0.07 (-0.15, 0.01) | 0.08 |
|  | Asia - MESA | Year | NA | NA | NA | NA |
|  |  | Year^2 | NA | NA | NA | NA |
|  |  |  | c. |  | c. |  |
|  | Australia | Year | NA | NA | NA | NA |
|  |  | Year^2 | NA | NA | NA | NA |
|  |  |  | c. |  | c. |  |
| MCiNP | International | Year | 0.01 (-0.54, 0.56) | 0.97 | -0.08 (-0.81, 0.65) | 0.79 |
|  |  | Year^2 | NA | NA | -0.02 (-0.13, 0.08) | 0.61 |
|  | Latin America | Year | -0.02 (-0.14, 0.09) | 0.48 | -0.05 (-0.50, 0.41) | 0.42 |
|  |  | Year^2 | NA | NA | -0.004 (-0.06, 0.05) | 0.5 |
|  |  |  | a. |  | a. |  |
|  | Africa | Year | NA | NA | NA | NA |
|  |  | Year^2 | NA | NA | NA | NA |
|  |  |  | c. |  | c. |  |
|  | Asia - EPA | Year | -0.58 (-1.15, -0.01) | 0.05 | -0.57 (-1.13, -0.01) | 0.05 |
|  |  | Year^2 | NA | NA | -0.10 (-0.19, -0.0002) | 0.05 |
|  |  |  |  |  | a. |  |
|  | Asia - MESA | Year | NA | NA | NA | NA |
|  |  | Year^2 | NA | NA | NA | NA |
|  |  |  | c. |  | c. |  |
|  | Australia | Year | NA | NA | NA | NA |
|  |  | Year^2 | NA | NA | NA | NA |
|  |  |  | c. |  | c. |  |
| MOP | International | Year | 0.01 (-0.09, 0.11) | 0.81 | -0.01 (-0.16, 0.14) | 0.86 |
|  |  | Year^2 | NA | NA | -0.01 (-0.03, 0.02) | 0.63 |
|  | Latin America | Year |  |  | NA | NA |
|  |  | Year^2 | NA | NA | NA | NA |
|  |  |  |  |  | c. |  |
|  | Africa | Year | NA | NA | NA | NA |
|  |  | Year^2 | NA | NA | NA | NA |
|  |  |  | c. |  | c. |  |
|  | Asia - EPA | Year | 0.01 (-0.09, 0.11) | 0.83 | 0.01 (-0.16, 0.17) | 0.91 |
|  |  | Year^2 | NA | NA | -0.0003 (-0.03, 0.03) | 0.98 |
|  | Asia - MESA | Year | NA | NA | NA | NA |
|  |  | Year^2 | NA | NA | NA | NA |
|  |  |  | c. |  | c. |  |
|  | Australia | Year | NA | NA | NA | NA |
|  |  | Year^2 | NA | NA | NA | NA |
|  |  |  | c. |  | c. |  |
| MCPP | International | Year | -0.10 (-0.26, 0.06) | 0.21 | -0.25 (-0.46, -0.03) | 0.03 |
|  |  | Year^2 | NA | NA | -0.02 (-0.04, 0.0001) | 0.05 |
|  | Latin America | Year | -0.01 (-0.31, 0.29) | 0.95 | -0.12 (-0.66, 0.43) | 0.6 |
|  |  | Year^2 | NA | NA | -0.01 (-0.07, 0.04) | 0.56 |
|  | Africa | Year | NA | NA | NA | NA |
|  |  | Year^2 | NA | NA | NA | NA |
|  |  |  | c. |  | c. |  |
|  | Asia - EPA | Year | -0.66 (-1.11, -0.22) | 0.01 | -0.45 (-1.27, 0.36) | 0.25 |
|  |  | Year^2 | NA | NA | -0.05 (-0.21, 0.11) | 0.49 |
|  | Asia - MESA | Year | NA | NA | NA | NA |
|  |  | Year^2 | NA | NA | NA | NA |
|  |  |  | c. |  | c. |  |
|  | Australia | Year | NA | NA | NA | NA |
|  |  | Year^2 | NA | NA | NA | NA |
|  |  |  | c. |  | c. |  |
| MBzP | International | Year | -0.25 (-0.44, -0.07) | 0.01 | -0.23 (-0.44, -0.01) | 0.04 |
|  |  | Year^2 | NA | NA | 0.01 (-0.02, 0.04) | 0.65 |
|  | Latin America | Year | -0.05 (-0.33, 0.22) | 0.67 | -0.21 (-0.63, 0.21) | 0.28 |
|  |  | Year^2 | NA | NA | -0.03 (-0.08, 0.03) | 0.28 |
|  | Africa | Year | NA | NA | NA | NA |
|  |  | Year^2 | NA | NA | NA | NA |
|  |  |  | c. |  | c. |  |
|  | Asia - EPA | Year | -0.30 (-0.51, -0.09) | 0.01 | -0.24 (-0.49, 0.01) | 0.06 |
|  |  | Year^2 | NA | NA | 0.02 (-0.02, 0.05) | 0.38 |
|  | Asia - MESA | Year | NA | NA | NA | NA |
|  |  | Year^2 | NA | NA | NA | NA |
|  |  |  | c. |  | c. |  |
|  | Australia | Year | NA | NA | NA | NA |
|  |  | Year^2 | NA | NA | NA | NA |
|  |  |  | c. |  | c. |  |
| MiDP | International | Year | -0.11 (-0.33, 0.10) | 0.1 | NA | NA |
|  |  | Year^2 | NA | NA | NA | NA |
|  |  |  | d. |  | c. |  |
|  | Latin America | Year | NA | NA | NA | NA |
|  |  | Year^2 | NA | NA | NA | NA |
|  |  |  | c. |  | c. |  |
|  | Africa | Year | NA | NA | NA | NA |
|  |  | Year^2 | NA | NA | NA | NA |
|  |  |  | c. |  | c. |  |
|  | Asia - EPA | Year | NA | NA | NA | NA |
|  |  | Year^2 | NA | NA | NA | NA |
|  |  |  | c. |  | c. |  |
|  | Asia - MESA | Year | NA | NA | NA | NA |
|  |  | Year^2 | NA | NA | NA | NA |
|  |  |  | c. |  | c. |  |
|  | Australia | Year | NA | NA | NA | NA |
|  |  | Year^2 | NA | NA | NA | NA |
|  |  |  | c. |  | c. |  |
| **Low Molecular Weight** | | | | | | |
| MEP | International | Year | -1.79 (-2.72, -0.85) | <0.01 | -1.35 (-2.39, -0.30) | 0.01 |
|  |  | Year^2 | NA | NA | 0.14 (-0.01, 0.28) | 0.07 |
|  | Latin America | Year | -6.42 (-10.47, -2.37) | 0.01 | -8.39 (-13.62, -3.16) | 0.01 |
|  |  | Year^2 | NA | NA | -0.44 (-1.16, 0.27) | 0.19 |
|  | Africa | Year | NA | NA | NA | NA |
|  |  | Year^2 | NA | NA | NA | NA |
|  |  |  | c. |  | c. |  |
|  | Asia - EPA | Year | -0.51 (-1.34, 0.32) | 0.22 | -0.06 (-0.94, 0.81) | 0.89 |
|  |  | Year^2 | NA | NA | 0.15 (0.03, 0.28) | 0.02 |
|  | Asia - MESA | Year | NA | NA | NA | NA |
|  |  | Year^2 | NA | NA | NA | NA |
|  |  |  | c. |  | c. |  |
|  | Australia | Year | NA | NA | NA | NA |
|  |  | Year^2 | NA | NA | NA | NA |
|  |  |  | c. |  | c. |  |
| MiBP | International | Year | -0.29 (-1.19, 0.61) | 0.53 | -0.26 (-1.30, 0.79) | 0.63 |
|  |  | Year^2 | NA | NA | 0.02 (-0.14, 0.18) | 0.82 |
|  | Latin America | Year | -0.20 (-1.60, 1.19) | 0.73 | -1.15 (-3.26, 0.95) | 0.22 |
|  |  | Year^2 | NA | NA | -0.11 (-0.30, 0.08) | 0.2 |
|  | Africa | Year | NA | NA | NA | NA |
|  |  | Year^2 | NA | NA | NA | NA |
|  |  |  | c. |  | c. |  |
|  | Asia - EPA | Year | -0.29 (-1.42, 0.83) | 0.61 | -0.20 (-1.41, 1.02) | 0.75 |
|  |  | Year^2 | NA | NA | 0.07 (-0.15, 0.29) | 0.52 |
|  | Asia - MESA | Year | NA | NA | NA | NA |
|  |  | Year^2 | NA | NA | NA | NA |
|  |  |  | c. |  | c. |  |
|  | Australia | Year | NA | NA | NA | NA |
|  |  | Year^2 | NA | NA | NA | NA |
|  |  |  | c. |  | c. |  |
| MnBP | International | Year | 0.93 (-0.39, 2.26) | 0.17 | 2.47 (0.99, 3.96) | <0.01 |
|  |  | Year^2 | NA*** | NA | 0.46 (0.23, 0.69) | <0.01 |
|  | Latin America | Year | -2.89 (-5.72, -0.06) | 0.05 | -3.75 (-7.23. -0.26) | 0.04 |
|  |  | Year^2 | NA | NA | -0.26 (-0.78, 0.27) | 0.28 |
|  | Africa | Year | NA | NA | NA | NA |
|  |  | Year^2 | NA | NA | NA | NA |
|  |  |  | c. |  | c. |  |
|  | Asia - EPA | Year | 1.43 (-0.02, 2.89) | 0.05 | 3.28 (1.70, 4.87) | <0.01 |
|  |  | Year^2 | NA | NA | 0.56 (0.32, 0.81) | <0.01 |
|  | Asia - MESA | Year | NA | NA | NA | NA |
|  |  | Year^2 | NA | NA | NA | NA |
|  |  |  | c. |  | c. |  |
|  | Australia | Year | NA | NA | NA | NA |
|  |  | Year^2 | NA | NA | NA | NA |
|  |  |  | c. |  | c. |  |
| MBzP | International | Year | -0.25 (-0.44, -0.07) | 0.01 | -0.23 (-0.44, -0.01) | 0.04 |
|  |  | Year^2 | NA | NA | 0.01 (-0.02, 0.04) | 0.65 |
|  | Latin America | Year | -0.05 (-0.33, 0.22) | 0.67 | -0.21 (-0.63, 0.21) | 0.28 |
|  |  | Year^2 | NA | NA | -0.03 (-0.08, 0.03) | 0.28 |
|  | Africa | Year | NA | NA | NA | NA |
|  |  | Year^2 | NA | NA | NA | NA |
|  |  |  | c. |  | c. |  |
|  | Asia - EPA | Year | -0.30 (-0.51, -0.09) | 0.01 | -0.24 (-0.49, 0.01) | 0.06 |
|  |  | Year^2 | NA | NA | 0.02 (-0.02, 0.05) | 0.38 |
|  | Asia - MESA | Year | NA | NA | NA | NA |
|  |  | Year^2 | NA | NA | NA | NA |
|  |  |  | c. |  | c. |  |
|  | Australia | Year | NA | NA | NA | NA |
|  |  | Year^2 | NA | NA | NA | NA |
|  |  |  | c. |  | c. |  |

*Weighted for each study's standard error; adjusted for geographical location, age group, and pregnancy status

**Weighted for each study's standard error; adjusted for geographical location, age group, pregnancy status, and quadratic term for time

*** NA = Not Available/Applicable

a. pregnancy status was omitted during the regression due to collinearity

b. Age group was omitted during the regression due to collinearity

c. Analysis was unfeasible due to limited or unavailable observations in the region

d. Age group and pregnancy was omitted during the regression due to collinearity

e. All variables besides the quadratic variable for time is omitted due to collinearity

f. All variables besides the time year of sampling is omitted due to collinearity

Supplement Figure 1: Predicted phthalate monoester concentrations from 2003 to 2023 in Latin America

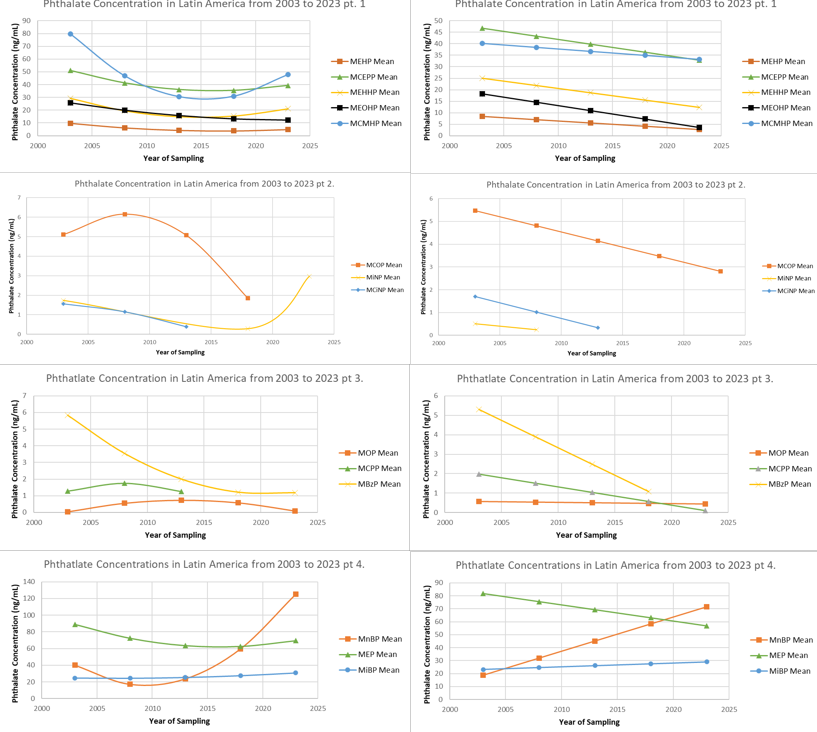

Note: If a monoester is not present, it is due to insufficient information.

Supplement Figure 2: Predicted phthalate monoester concentrations from 2003 to 2023 in Africa.

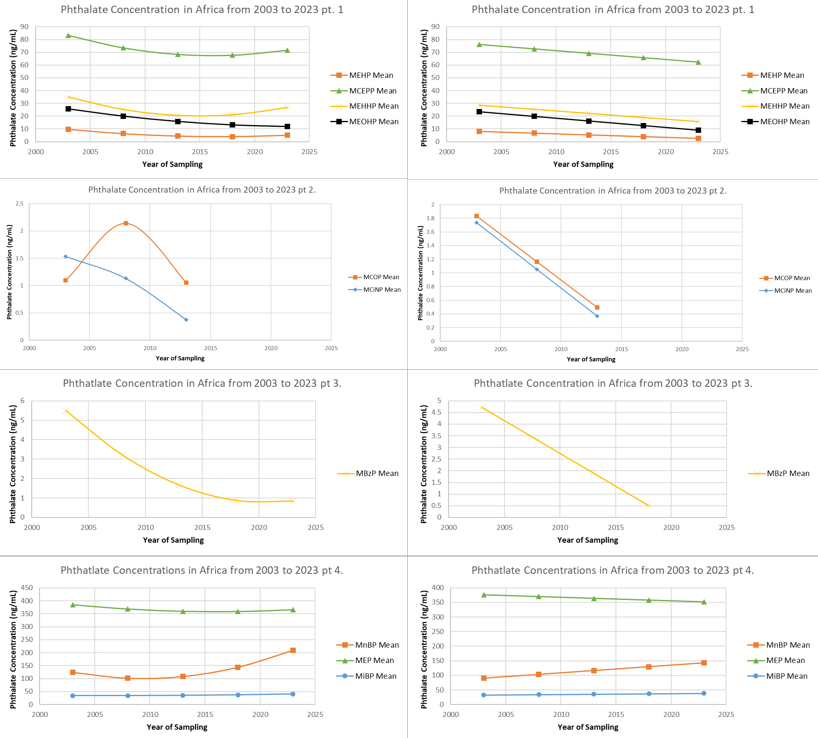

Note: If a monoester is not present, it is due to insufficient information.

Supplement Figure 3: Predicted phthalate monoester concentrations from 2003 to 2023 in Easter and Pacific Asia

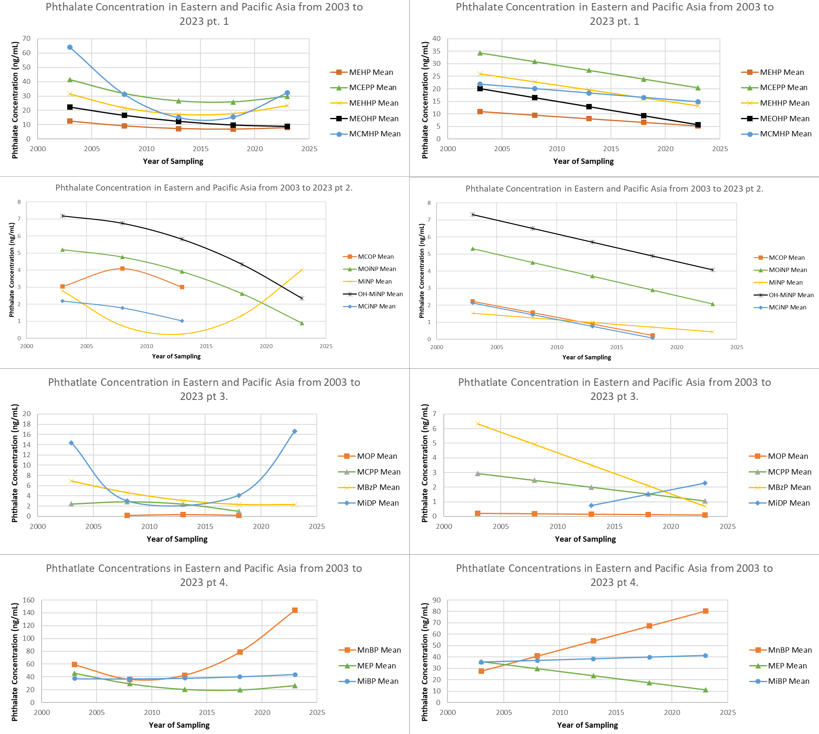

Note: If a monoester is not present, it is due to insufficient information.

Supplement Figure 4: Predicted phthalate monoester concentrations from 2003 to 2023 in Middle East and South Asia

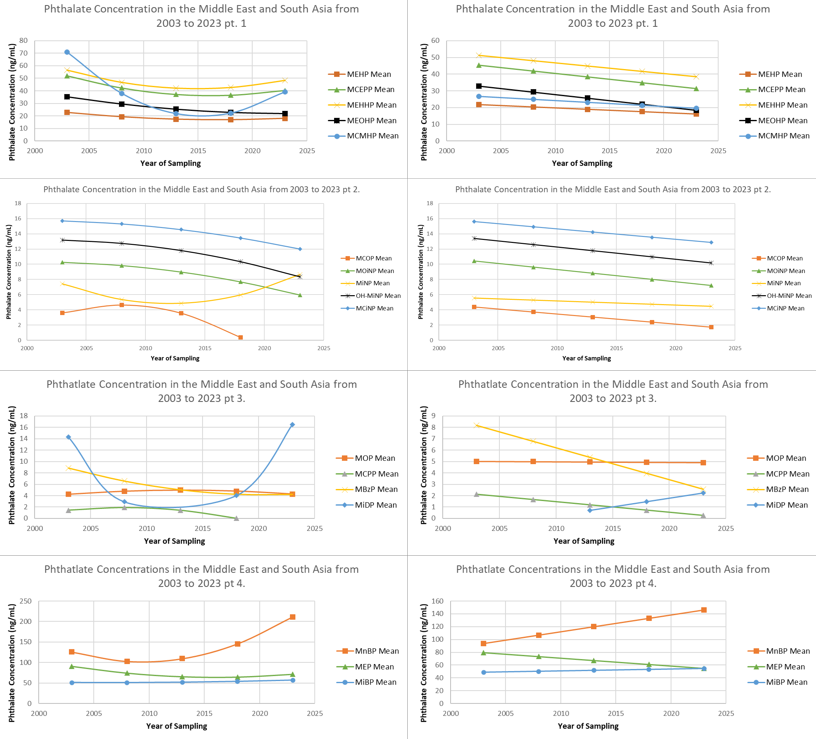

Note: If a monoester is not present, it is due to insufficient information.

Supplement Figure 5: Predicted phthalate monoester concentrations from 2003 to 2023 in Australia

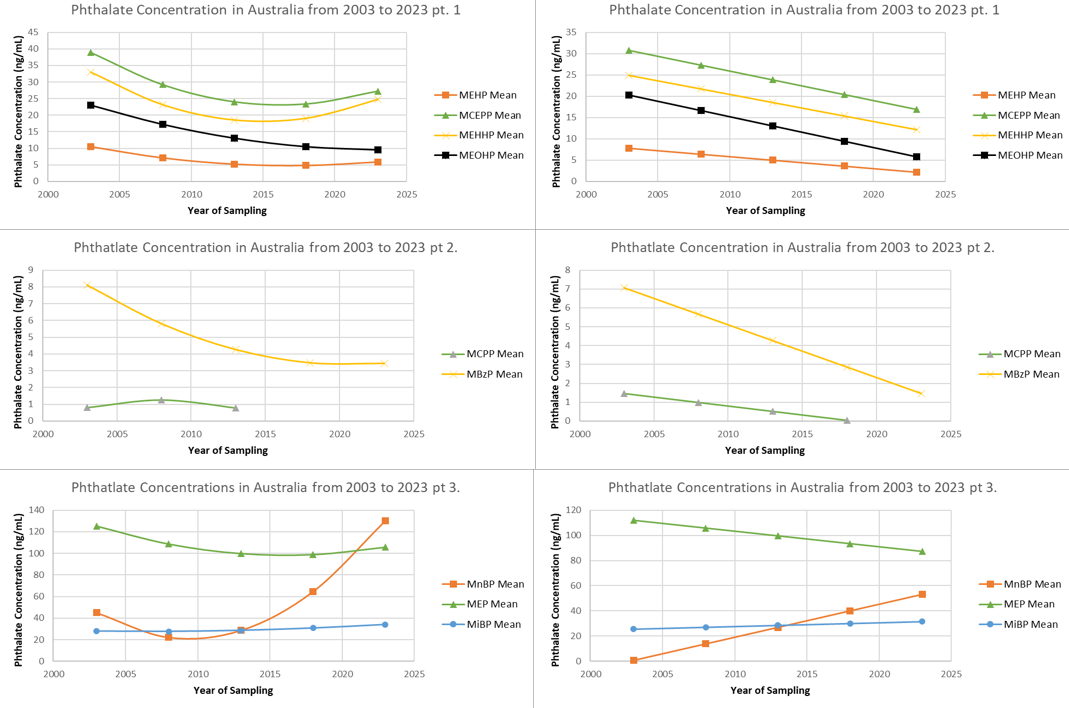

Note: If a monoester is not present, it is due to insufficient information.
